# Enterovirus D68 in United States wastewater is associated with climatic and demographic factors: a comparison with clinical diagnoses

**DOI:** 10.1101/2025.11.03.25339422

**Authors:** Sheena Conforti, Alessandro Zulli, Alexandria Boehm

## Abstract

Enterovirus D68 (EV-D68) is a respiratory virus that can cause severe illness and is associated with acute flaccid myelitis (AFM), yet testing is uncommon and no vaccine exists. Here, we analyzed 43,876 wastewater samples collected from 147 U.S. treatment plants across 40 states between July 2023 and July 2025. We quantified EV-D68 RNA in wastewater solids to identify the center of season and the season duration of EV-D68, assessed climatic and sociodemographic factors of variation, and compared wastewater trends to clinical diagnoses for AFM, wheezing, and enterovirus-specific encounters from Epic Cosmos. At national level, the EV-D68 center of season occurred in late summer 2024, showing a biennial pattern and wide geographic variation: detection occurred earlier in southern states and duration was longer in denser, more urban communities with higher population density, more hospitals, nursing homes, crowded households, and childcare facilities. Temperature and dew point were the strongest correlates of the center of season. Nationally, EV-D68 RNA did not correlate with diagnoses of AFM or wheezing, likely due to underdiagnosis and reporting limitations, but weak to moderate correlations with enterovirus-coded encounters were observed. Our findings demonstrate that wastewater monitoring can detect EV-D68 at scale, even when clinical data are sparse or delayed, and reveal that the center of season is associated with environmental variables, while duration is associated with sociodemographic factors. These results underscore the utility of wastewater surveillance as a timely, actionable tool for public health response in the absence of routine testing or vaccination.

**Significance Statement:** It is important to detect and respond to Enterovirus D68 (EV-D68) outbreaks because the virus can cause severe respiratory illness and neurological complications in children. However, EV-D68 is rarely tested in clinics, making it hard to track. This study shows that wastewater can be used to monitor EV-D68 across the United States in real time. We found that warm, humid climates lead to earlier outbreaks, and that the virus lasts longer in urban areas with higher population density, crowded households, and more nursing homes, hospitals, and childcare centers. These findings can help health practitioners target testing and prevention efforts. Wastewater surveillance offers a scalable, timely tool for tracking underdiagnosed respiratory viruses as climate and transmission patterns shift.

## Introduction

Enterovirus D68 (EV-D68) is a non-enveloped picornavirus transmitted primarily via the respiratory route and for which no vaccine is available (1). Symptoms range from mild upper respiratory illness to severe lower respiratory disease with dyspnea, wheezing, hypoxia and, especially in children, can involve central nervous system complications (CNS) such as acute flaccid myelitis (AFM), cranial nerve dysfunction, encephalitis, and aseptic meningitis (1). AFM cases have already been shown to increase after EV-D68 outbreaks (2), and since 2014, the U.S. Centers for Disease Control and Prevention (CDC) has conducted active AFM surveillance, documenting 779 confirmed cases as of Aug 5, 2025 (3), and biennial peaks consistent with observed EV-D68 infections (4).

Diagnosing and tracking EV-D68 remains difficult because clinical testing is not a standard practice in most laboratories, and the virus is typically identifiable in nasopharyngeal specimens only during the first week after CNS symptom onset (1). Population-level wastewater surveillance could facilitate the detection of EV-D68 circulation at scale, since viral RNA is occasionally detected in stool (5,6). Proof-of-concept studies have shown that wastewater EV-D68 RNA tracks trends in laboratory-confirmed cases in California and internationally (7–11). However, it remains unclear whether wastewater signals serve as a proxy for broader clinical diagnoses such as AFM, wheezing, and enterovirus-specific encounters.

Furthermore, the drivers of EV-D68 seasonality and its biennial pattern remain uncertain. Enterovirus circulation in the U.S. is linked to climatic factors, including temperature, dew point, humidity, rainfall, wind speed, sunshine, and air pressure (12–14). In temperate regions, enteroviruses tend to peak in summer and early autumn (15) with flatter, earlier seasons in the south and sharper, later peaks in the north (13). A latitudinal gradient has been reported specifically for EV-D68 (2). Demographic factors such as school-term timing (12) and birthrates (13) have also been associated with outbreak timing and duration. Yet, most analyses are at the state level, where microclimate and community heterogeneity may be masked. Given diagnostic challenges, higher-resolution understanding of climatic and demographic drivers would help clinicians anticipate when the virus is circulating and focus testing accordingly to reduce the risk of severe outcomes such as AFM.

Here, we analyzed wastewater from 147 U.S. wastewater treatment plants (WWTPs) collected between July 2023 and July 2025 to quantify EV-D68 concentrations and characterize spatiotemporal patterns by estimating the center of season and season duration in each location. We then used climatic and demographic indicators to examine spatial differences in center of season and season duration. Finally, we retrieved diagnostic data from Epic Cosmos, a dataset created in collaboration with a community of Epic health systems representing more than 300 million patient records from more than 1,800 hospitals and 41,000 clinics from all 50 U.S. States, D.C. Lebanon, and Saudi Arabia as of October 2025, to compare wastewater trends with clinical diagnoses for AFM, wheezing, and enterovirus-specific encounters. Overall, wastewater-derived timing metrics, alongside climatic and demographic drivers, may serve as early, actionable signals to prioritize EV-D68 testing and clinical readiness where and when activity is likely to intensify.

## Results

### Spatio-temporal analysis of wastewater EV-D68

Between July 2023 and July 2025, 43,876 wastewater solids samples were collected from 147 WWTPs across the United States. Across all wastewater treatment plants (WWTPs), between 178 and 694 samples were analyzed per site (Table S1). EV-D68 RNA was detected in 24% of all samples analyzed, with concentrations ranging from 580 to 4.8 × 10^6^ gene copies per gram dry weight (gc/g) (median = 8.1 × 10^3^ gc/g). Detection frequency varied substantially by location: from 0.7% of samples in Clinton, Iowa, to 82% in San Jose, California (Table S2). EV-D68 RNA was infrequently detected between July 2023 and early February 2024 (5% of samples), after which detections became more common (29% of samples) (Figure 1).

**Figure 1.**
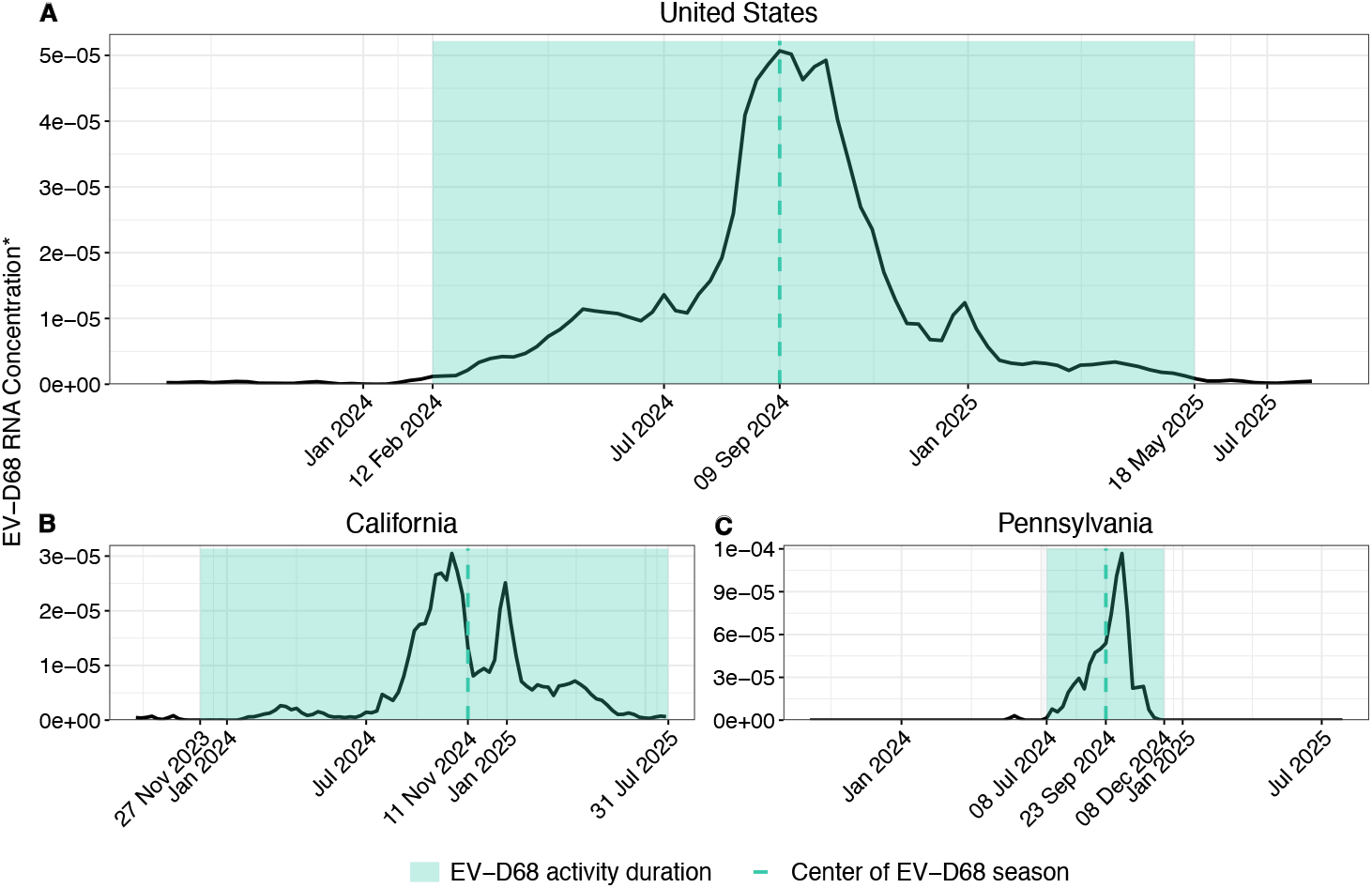
EV-D68 RNA in wastewater at national and state levels, 2023–2025. Smoothed, PMMoV-normalized EV-D68 RNA concentration for (A) United States, (B) California, and (C) Pennsylvania. The shaded band denotes the EV-D68 activity duration; the vertical line marks the center of the EV-D68 season. For the national series (A), duration was defined as the period when concentrations exceeded a threshold equal to a baseline plus three standard deviations, where the baseline and standard deviation were estimated from the quiet tail (lowest tertile) of the data distribution. For state series (B–C), duration was defined as the continuous run containing the center of season during which values were detectable, with a minimum of at least two consecutive weeks with EV-D68 RNA concentration above 0. Dates with day-of-month annotate the start and end of the duration and the season center in each state. *EV-D68 RNA concentration refers to the smoothed, PMMoV-normalized concentration, calculated using a 5-day centered trimmed moving average. For each 5-day window, the highest and lowest daily values were excluded, and the remaining three were averaged.

The national EV-D68 center of season, defined as the week during which most of the seasonal EV-D68 detection was concentrated, occurred in late summer 2024 on September 9, 2024. The season duration was estimated as the continuous period of elevated national EV-D68 RNA concentrations surrounding the center of season, and at national level it lasted 66 epidemiological weeks: from February 12, 2024, to May 18, 2025.

At the state level, center of season ranged from early June to November 2024, occurring earlier in southern and southeastern states (June–August), and progressively later in northern and western regions (September–November) (Figure 2A, Figure S1). Most states exhibited relatively short season duration (≤5 months), while few states, notably California and Florida, showed prolonged duration (>13 months) (Figure 2B). Season duration in California spanned from November 27, 2023, to July 31, 2025, with a center of season on November 11, 2024 (Figure 1B), whereas for example Pennsylvania experienced a shorter season duration from July 8 to December 8, 2024, centered on September 23, 2024 (Figure 1C), aligning with the national center of season.

**Figure 2.**
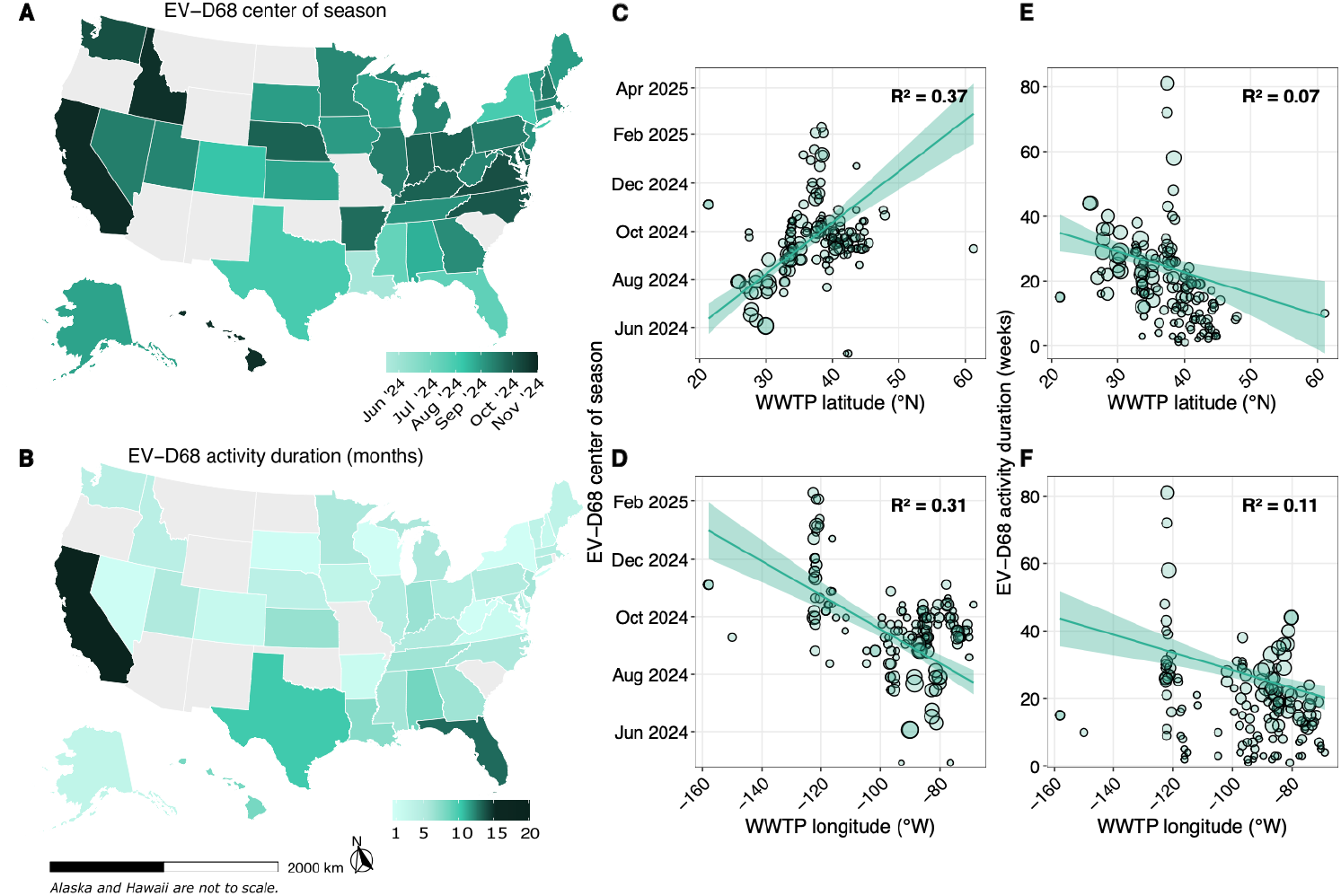
State-level timing and duration of EV-D68 activity in U.S. wastewater, with WWTP-level geographic gradients. (A) Map shows, for each state, the calendar date of the EV-D68 seasonal center estimated from the state-aggregated weekly series (PMMoV-normalized, 5-day–trimmed), after first aggregating across WWTPs within state and week. Colors run from earlier (Jun ‘24) to later (Nov ‘24). (B) Map shows the length of the detectable activity window (displayed in months) around the seasonal center of the state, computed on the same state-aggregated weekly series by identifying consecutive weeks with detectable signal (runs of at least two weeks above the non-detect threshold) and converting weeks to months. Gray states do not have wastewater data available. Alaska and Hawaii are shown out of scale for layout. Panels C–F are weighted bivariate regressions displaying gradients between longitude or latitude and EV-D68 center of season (C and D) or activity duration (E and F). Each point is a WWTP within the United States (n=146) Clinton, Iowa had W=0 (no detections after normalisation and trimming) and therefore was excluded. Point size denotes the plant weight WW, defined as the sum across weeks of PMMoV-normalised, 5-day–trimmed EV-D68 concentrations, with non-detects set to 0. Larger circles indicate greater cumulative signal and therefore greater influence on the weighted least-squares fits. Lines show weighted least-squares fits with 95% CIs.

Weighted linear regression analyses using data from 146 WWTPs indicated that the EV-D68 center of season occurred significantly later in the north than in the south (6.5 days per degree latitude; 95% CI 5.1–7.9; p<0.001; *R*_*w*_^*2*^= 0.37) and toward the west (slope −1.8 days per degree longitude; 95% CI −2.3 to −1.4; p<0.001; *R*_*w*_^*2*^= 0.31) (Figure 2C and D). EV-D68 season duration also shortened with increasing latitude (−4.6 days per degree; 95% CI −7.4 to −1.7; p<0.001; *R*_*w*_^*2*^= and decreased towards the west (−1.8 days per degree; 95% CI −2.7 to −1.0; p<0.001; *R*_*w*_^*2*^= 0.11) (Figure 2E–F), though absolute effect sizes were small.

### Environmental variables and EV-D68 season

Temperature and dew point were dominant correlates of EV-D68 timing, with higher temperature and higher atmospheric moisture (dew point) associated with earlier seasonal centers (Figure 3). After adjustment for site latitude and longitude, a 5 °C higher weekly temperature was associated with an earlier seasonal center by 28.1 days (95% CI −33.7 to −22.6; p<0.001), and a 5 °C higher weekly dew point by 31.3 days (95% CI −37.0 to −25.6; p<0.001) (Table 1). Even after accounting for latitude and longitude, model fit improved: coordinates alone explained 59% of the weighted variance, which rose to 76% when temperature was added and to 78% with dew point. Surface pressure also showed an independent association, with later centers at higher pressure (+15.6 days per 5 hPa; 95% CI 9.3 to 21.8; p<0.001; *R*_*w*_^*2*^=0.06). By contrast, relative humidity, precipitation, and wind speed had no association with EV-D68 timing, either in bivariate analyses or after adjustment for latitude and longitude. As expected, temperature and dew point were highly correlated (Figure S2), and their adjusted estimates reflect the same underlying warm, moist conditions and should not be interpreted as additive. Analyses showed no associations between EV-D68 season duration and environmental variables (effects near zero; weighted R^2^ ≤ 0.04) (Figure S3; Table S3).

**Table 1.**
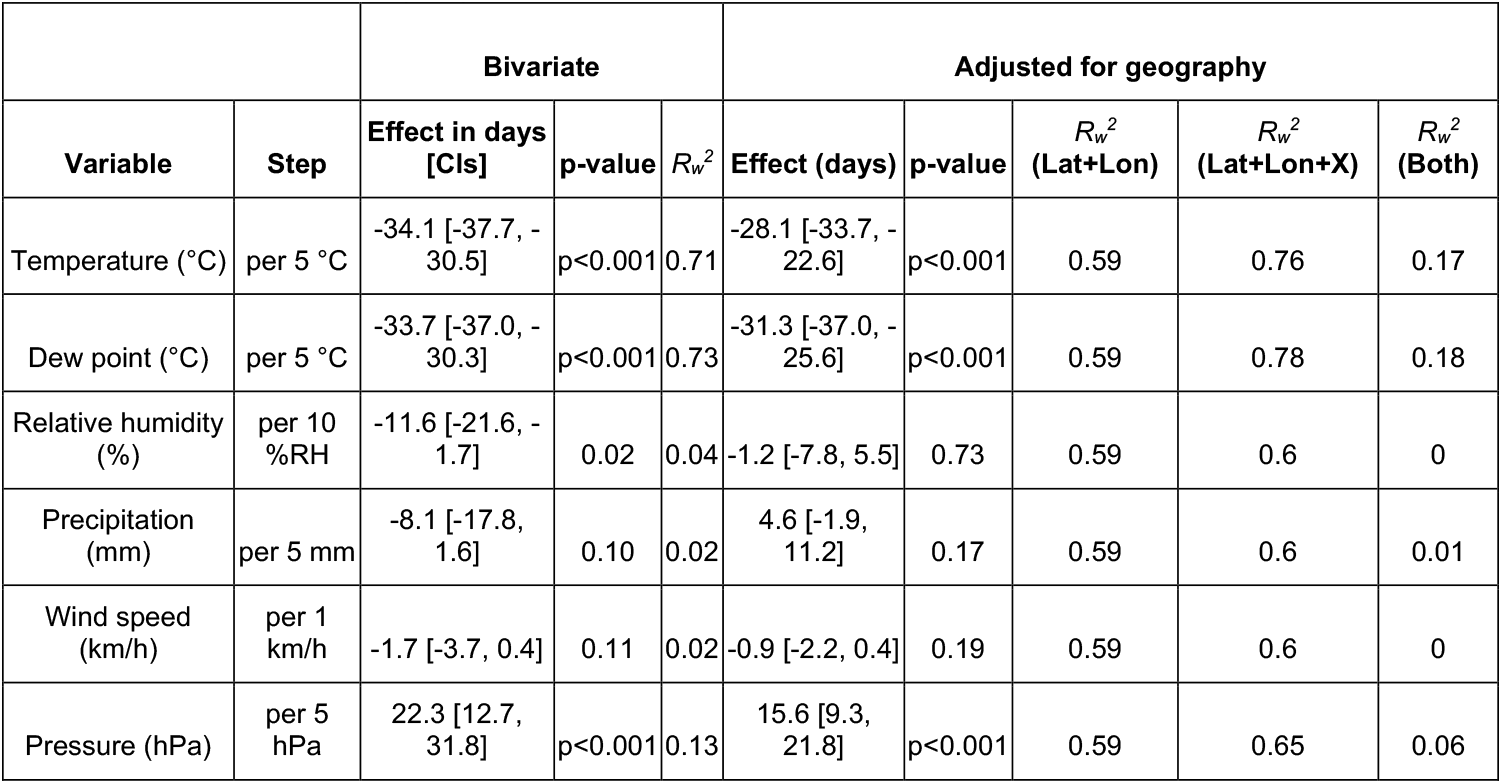
Effects of weekly environmental variables on the calendar date of the local EV-D68 seasonal center in wastewater. Effects are expressed in days, with negative values denoting earlier timing and positive values denoting later timing. Effects were estimated using weighted least-squares models fitted first without geographic covariates (bivariate) and then with geographic adjustment for latitude and longitude (Lat+Lon+X). For interpretability, effects correspond to fixed increments (temperature and dew point 5 °C; relative humidity 10%; precipitation 5 mm; wind speed 1 km/h; pressure 5 hPa). Weighted *R*_*w*_^*2*^is shown for the bivariate model and for the geography-only baseline (Lat+Lon) and the geography-adjusted model (Lat+Lon+X); “*R*_*w*_^*2*^(Both)” gives the gain (*R*_*w*_^*2*^) from adding the climate variable to geography. Models used plant-level weights proportional to cumulative EV-D68 signal after trimming (non-detects set to zero). Brackets denote 95% confidence intervals (CIs).

**Figure 3.**
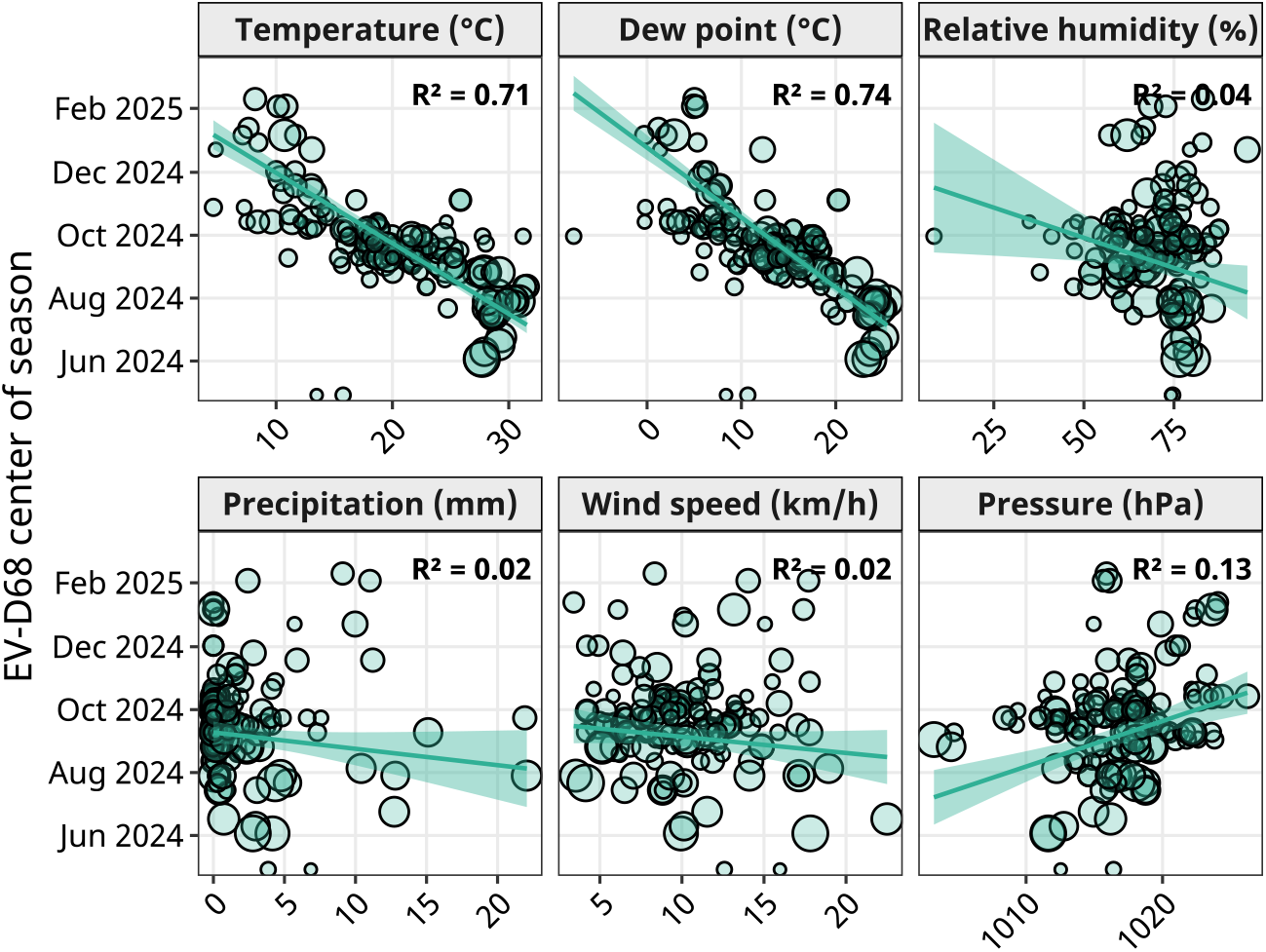
EV-D68 center of season as a function of mean environmental conditions across WWTPs (n = 146). For each plant, the x-axis is the mean of the indicated variable (daily values aggregated to weekly means over the EV-D68 activity window); the y-axis is the calendar date of the season center. Lines are univariate weighted least-squares fits with 95% CIs; weights are proportional to each plant’s cumulative EV-D68 signal, with non-detects set to 0. Point sizes are proportional to the weights and the weighted R^2^ is shown on each panel.

### Sociodemographic and structural variables of EV-D68 season

Higher childcare density, household crowding, number of hospitals, number of nursing homes, population density, and urbanicity were each associated with a longer EV-D68 season duration (Figure 4; Table S4). EV-D68 season duration was notably longer in catchments with greater urbanicity (median [IQR] 20 [13-28] weeks for >50% urban and 12 [4-18] weeks for ≤50%; p<0.001) and higher population density (median 24 [15-29] weeks in the highest tertile and 13 weeks [5-21] in the lowest; p<0.001). Childcare establishment density showed a strong gradient, with the longest duration in the highest tertile (median 25 [19-31] weeks) compared to the middle (15 [7-22] weeks) and lowest (14 [7.8-23] weeks) tertiles (p<0.001 for both comparisons). Household crowding, namely the proportion of households with more than one person per room, was similarly associated with duration (median 25 [16-31] weeks in the highest tertile vs 15 [6.8-21] weeks in the lowest; p<0.001). Catchments above the dataset median for hospitals (>2 per catchment) and nursing homes (>8 per catchment) also experienced longer seasons (median 22 [16-29] and 15 [8.5-23] weeks for hospitals, p=0.002; 23.5 [15-31] and 15 [8-20.8] weeks for nursing homes, p<0.001). A modest association was observed with the proportion of adults aged ≥65 years, with the highest median duration in the middle tertile (21 [16-29] weeks) compared to the lowest (16 [10-23] weeks, p=0.03). No associations were observed with the proportion of children aged ≤5 years (p=0.61), birth rate (p=0.16), or the presence of an airport in the catchment (p=0.11). Variable distributions and pairwise correlations are presented in Figure S4 and Figure S5. As expected, sociodemographic and structural variables showed no meaningful association with EV-D68 center of season. The only exception was a moderate association with childcare density: lower childcare density was linked to a later center of season (Figure S6, Table S5).

**Figure 4.**
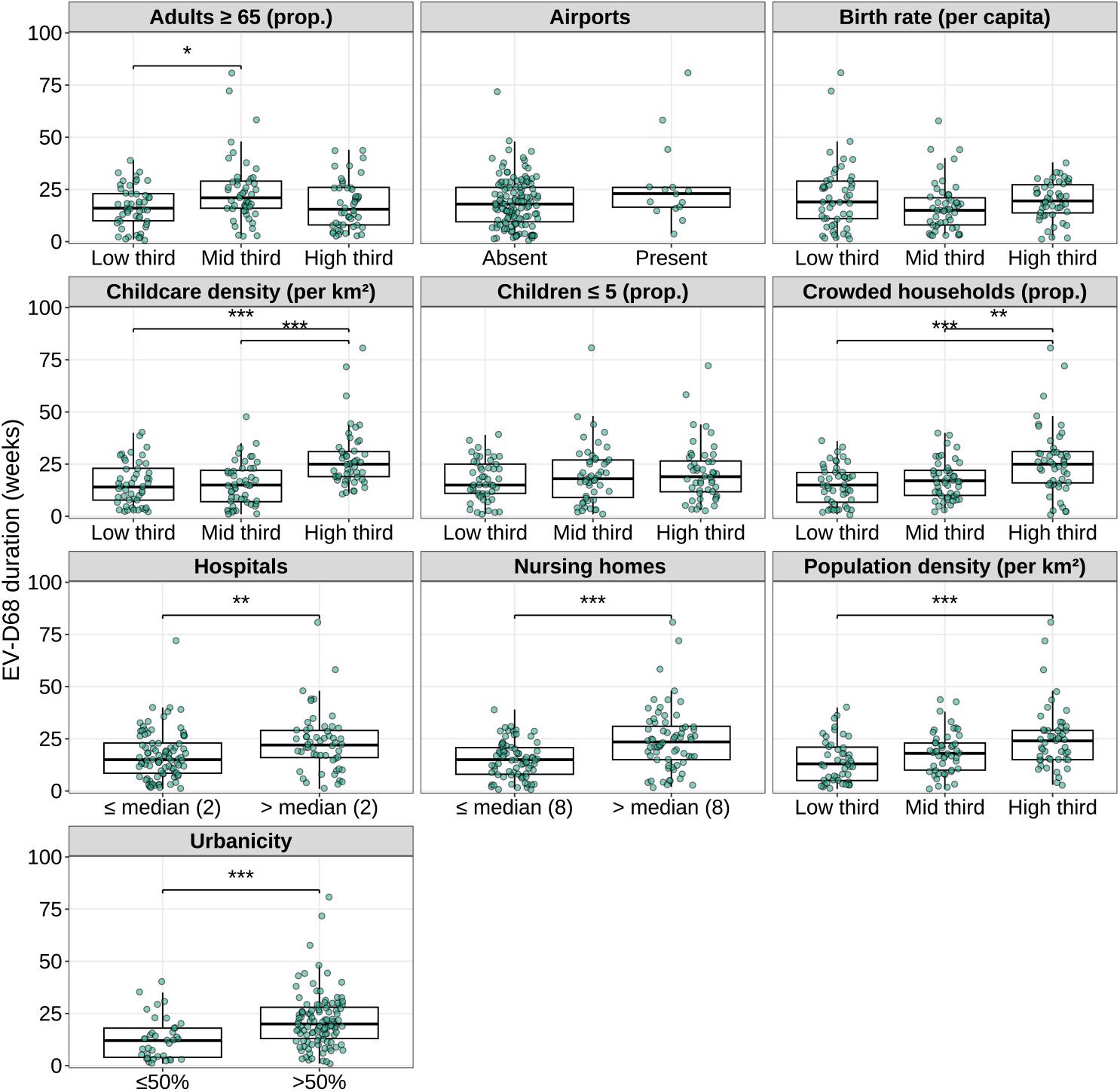
Duration of EV-D68 circulation by wastewater treatment plant (WWTP)-level characteristics. Each panel shows the distribution of EV-D68 circulation duration (in weeks) across categories of a single WWTP–level determinant. Points represent individual WWTPs and boxplots show the median, interquartile range (IQR), and whiskers at 1.5×IQR. Airport presence is dichotomised as absent and present. Hospital and nursing home coverage are split using the dataset medians: 2 hospitals and 8 nursing homes within each WWTP catchment area, respectively. Urbanicity is defined by the proportion of the WWTP catchment area classified as urban (≤50% vs >50%). The proportions of children aged ≤5 years, adults aged ≥65 years, crowded households, birth rate (per capita), childcare density (per km^2^), and population density (per km^2^) are grouped into within-study tertiles (“low”, “middle”, “high”). Kruskal–Wallis test was used to assess overall differences, and pairwise comparisons used Wilcoxon rank-sum tests with Bonferroni adjustment. Horizontal connector bars indicate statistically significant differences; asterisks denote adjusted p-values (*p<0.05, **p<0.01, ***p<0.001).

### Clinical diagnoses comparison with wastewater EV-D68

Nationally, wastewater EV-D68 RNA concentrations did not track clinical diagnostic proportions over the study period (Figure 5A–D). Wastewater showed a pronounced seasonal increase in autumn 2024, whereas clinical diagnoses were comparatively constant or diffuse. Enterovirus-specific diagnoses displayed a weak positive correlation with wastewater (Spearman ρ = 0.34, p = 0.005), while wheezing in adults aged ≥ 65 years correlated negatively (ρ = −0.49, p < 0.001) (Figure S7). No association was observed for wheezing in children ≤ 5 years (ρ = −0.03 0, p = 0.82) or for AFM (ρ = −0.13, p = 0.31). Correlation patterns varied across states (Figure S8; Table S6). AFM analyses were limited by masked monthly counts when diagnoses were ≤10, yielding few paired months per state and no Bonferroni-significant associations (Figure 5E). Nevertheless, positive correlations were observed in Ohio (ρ = 0.73, number of pairs months (n) = 8, p = 0.310), Tennessee (ρ = 0.36, n = 10, p = 1), and Georgia (ρ = 0.21, n = 10, p = 1), whereas negative but non-significant correlations appeared where paired months were few, such as in New York (ρ = −0.32, n = 4, p = 1) and Wisconsin (ρ = −0.54, n = 4, p = 1). Among the diagnoses investigated, enterovirus showed the broadest evidence of positive association across states, with at least weak positive correlations in 18 states (45%). Even so, strong positive and significant correlations were detected only in Kentucky (ρ = 0.72, n = 27, p < 0.01), North Carolina (ρ = 0.65, n = 28, p < 0.01), and Georgia (ρ = 0.59, n = 31, p = 0.02) (Figure 5F). Correlations between wastewater EV-D68 RNA concentrations and wheezing in children ≤ 5 years were heterogeneous and not significant overall, aligning with the national result (Figure 5G). In contrast, wheezing in adults ≥ 65 years tended to correlate negatively with wastewater, with strong negative and significant correlations in Georgia (ρ = −0.62, n = 31, p = 0.01), Florida (ρ = −0.82, n = 61, p < 0.01), and Texas (ρ = −0.84, n = 48, p < 0.01) (Figure 5H).

**Figure 5.**
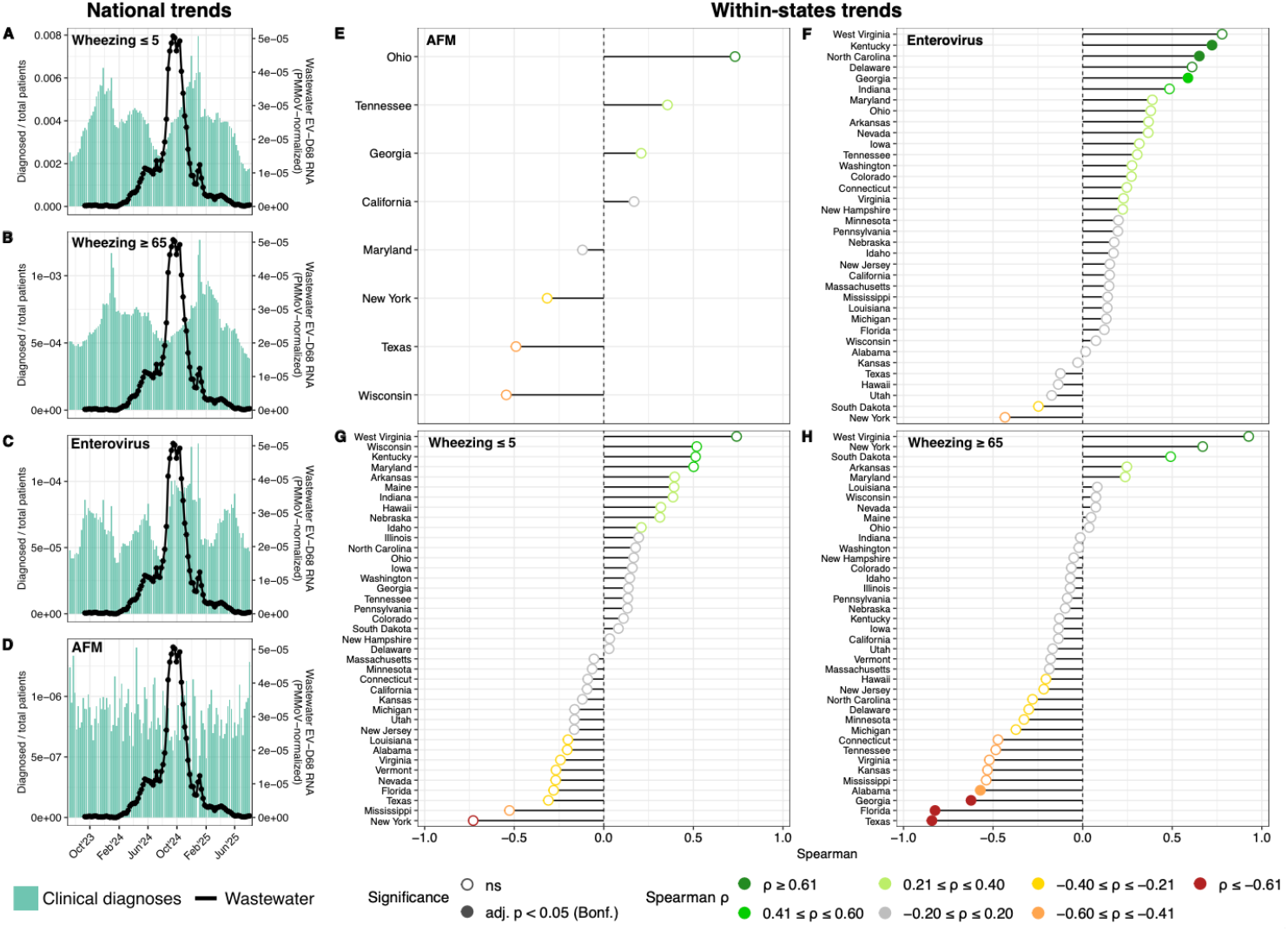
National and within-state correlations between wastewater EV-D68 RNA concentrations and clinical diagnoses. (A–D) National time-series comparing wastewater EV-D68 RNA concentrations (black lines, right axis) with proportions of clinical diagnoses (bars, left axis) for wheezing in children ≤ 5 years (A), wheezing in adults ≥ 65 years (B), enterovirus-specific encounters (C), and acute flaccid myelitis (AFM) (D). (E–H) State-level Spearman correlations between wastewater EV-D68 and AFM (E), enterovirus-specific encounters (F), wheezing ≤ 5 years (G), and wheezing ≥ 65 years (H). Points represent correlation coefficients, with colors denoting effect size and significance (adjusted p < 0.05, Bonferroni). Correlations at the state level were performed using data filtered to the EV-D68 seasonal duration defined by wastewater and extended ±2 weeks for clinical diagnoses, and ±2 months for AFM.

## Discussion

Enterovirus D68 (EV-D68) is a respiratory virus with no available vaccine that typically causes mild cold-like symptoms, but can also lead to severe respiratory illness and acute flaccid myelitis (AFM). Clinical surveillance is challenging because routine testing is uncommon and laboratory methods are not standardized. Here, we analyzed EV-D68 RNA concentrations in wastewater solids from 147 wastewater treatment plants across 40 states in the United States, using over 43,000 samples collected between July 2023 and July 2025 to characterize seasonal timing and duration of EV-D68, and their environmental and sociodemographic drivers. We observed a biennial pattern, with limited activity in 2023 and 2025, and widespread detections in summer-fall 2024, but with substantial variation in when and how long EV-D68 circulated across states. Overall, environmental factors were associated with when EV-D68 was detected in wastewater solids, whereas sociodemographic conditions were associated with how long it persisted.

EV-D68 center of season at national level occurred in early September 2024, consistent with the typical summer-to-autumn circulation observed for other enteroviruses (15). Clinical surveillance data from previous years have described EV-D68 as one of the latest-entering enteroviruses, with a mean timing of confirmed cases in September (13), and a tendency to recur every two years (9,16). An unlikely alternative for the near-absence of EV-D68 signal in 2025 wastewater is molecular assay mismatch from viral sequence changes reducing primer/probe specificity, but this is no evidence to support this. Further, there are no 2025 EV-D68 sequences publicly available for *in-silico* evaluation (as of August 20, 2025). Given the well-established biennial pattern of EV-D68 clinical activity (2), a routine inter-epidemic low-activity year is the plausible explanation. The timing and duration of EV-D68 activity varied across states, with centers of season ranging from early summer to late autumn 2024, and duration spanning from a few weeks to over a year, aligning with broad spatial heterogeneity in clinical EV-D68 seasonality in the United States, where timing varied from July to November (2).

EV-D68 center of season and season duration followed a latitudinal and longitudinal gradient. The center of season occurred earlier in the south and in the east, explaining 37% and 31% of the variance across sites, respectively. These findings align closely with previous analyses of clinical EV-D68 cases, which reported a latitudinal increase and longitudinal decrease in timing (2), but also with previous findings on non-polio enteroviruses in general, in which the latitude was associated with the timing of non-polio enterovirus peaks (R2=0.35 previously; R2=0.37 in our analysis) (13). Season duration was shorter in both northern and western locations, but the explanatory power of these associations was limited (R^2^ = 0.07 for latitude; R^2^ = 0.11 for longitude), suggesting that latitude and longitude are weakly associated with how long EV-D68 circulates in each location. California had the longest season observed in the study, extending from late 2023 to mid-2025. Prior studies predicted a large and lasting outbreak of EV-D68 in California in 2016, and only a small one in 2018 (2), suggesting that this very long 2024 season may precede a lower-intensity outbreak in 2026. Although the state lies in the west, its inclusion to more southerly latitudes may help explain the prolonged activity observed. Some enteroviruses have shown to have two outbreaks per year closer to the tropics (17), which could help explain the long and possibly multi-wave pattern observed in California, consistent with modeling studies suggesting that recent biennial EV-D68 cycles may be unstable (2).

Temperature and dew point were the strongest environmental correlates of EV-D68 center of season across the United States, explaining 71% and 74% of the variance, respectively. These findings align with earlier studies showing that enterovirus activity often follows periods of elevated temperature (14). Furthermore, dew point has been shown to significantly predict enterovirus transmission (12) and humidity metrics have been demonstrated to reflect the geographic structure of enterovirus transmission in the U.S. (13). However, it is difficult to disentangle the collinearity between temperature and dew point and to isolate their independent effects. Dew point reflects the amount of moisture in the air, which increases with both temperature and humidity, making it a reliable indicator of warm and humid atmospheric conditions (18). The association between higher dew point and earlier season timing suggests that EV-D68 tends to circulate earlier in environments that are both warm and moist. Indeed, viability experiments of respiratory viruses performed in aerosol chambers with controlled temperature and relative humidity indicated a striking correlation of the stability of summer viruses (such as EV-D68) at high relative humidity percentages (19). These associations may reflect that for EV-D68, warm and humid conditions create favorable microclimates for droplet-mediated transmission, maintaining viral stability and prolonging the survival of infectious particles in the air and on mucosal surfaces (20). Low ambient humidity can further impair mucociliary clearance and innate antiviral defenses, whereas warm, moist air preserves airway physiology that supports efficient viral replication (21). The strong explanatory power of these variables reinforces the importance of broad environmental conditions, rather than specific weather events, in influencing the seasonal dynamics of EV-D68. Interestingly, no environmental variable was linked to season duration, indicating that climate may help shape when EV-D68 activity begins in each area, but not how long it persists.

EV-D68 season duration appeared more closely linked to human and structural factors than to environmental variables. Longer seasons were observed in areas with higher population density, greater urbanicity, more crowded households, and higher densities of childcare facilities, hospitals, and nursing homes. These characteristics reflect opportunities for sustained person-to-person transmission and repeated introductions in dense or highly connected communities. Age structure was not associated with duration, even though children and older adults often present with more severe symptoms (1). This suggests that prolonged community circulation is maintained through overall contact intensity rather than age-specific susceptibility. Although previous studies have connected birthrates with enterovirus dynamics (13), we did not observe such effects here, possibly because birth rate data were estimated from 2024 and only partially overlapped the season duration estimated from wastewater solids. These patterns indicate that once EV-D68 becomes established, transmission can persist through frequent interpersonal contacts and continuous reintroduction in densely connected populations. Higher childcare density could plausibly advance EV-D68 circulation by promoting frequent close-contact interactions among young children, a key group for transmission and amplification (22). The center of season was not correlated with any sociodemographic variable except childcare density, which showed a moderate association whereby areas with fewer childcare facilities tended to experience later circulation. This association should, however, be interpreted cautiously, as it may reflect spatial variability or residual confounding with broader regional patterns. Together, these findings suggest that while environmental conditions trigger the onset of EV-D68 detection, namely when it occurs, demographic structure and social mixing determine how long it persists within a community.

EV-D68 infections in children can lead to AFM, and previous studies have shown that AFM cases typically increase about one month after a rise in EV-D68 infections, indicating a strong spatiotemporal association between the two (2). Furthermore, in 2016, 29 AFM cases in Europe and 149 in the United States were linked to EV-D68 (15). In our analysis, we did not detect a clear national-level association between EV-D68 RNA concentrations in wastewater solids and AFM diagnoses. However, in Ohio, Tennessee, and Georgia, EV-D68 RNA concentrations in wastewater solids increased concurrently with a rise in AFM encounters retrieved from Epic Cosmos. Because Epic Cosmos masks counts ≤10, many state-level data were missing or zero-inflated, likely obscuring true associations. This data limitation, combined with the rarity of AFM, probably explains why we did not detect significant correlations. These findings highlight the scarcity of clinical data for rare outcomes and underscore the potential of wastewater surveillance to provide early warning for pathogens that are underdetected in clinical settings. At the national level, EV-D68 RNA concentrations were moderately correlated with diagnoses of “enterovirus infection” in Epic Cosmos. Because EV-D68-specific testing is not routinely performed and laboratory methods remain non-standardized, enterovirus diagnoses likely capture a broad spectrum of infections. Most individuals infected with enteroviruses remain asymptomatic or experience only mild illness and are therefore unlikely to undergo clinical testing (7). The persistence of enterovirus diagnoses even in periods without EV-D68 detection suggests that these reflect other enterovirus species, whereas increases during EV-D68 activity indicate a substantial contribution of this virus to overall enterovirus circulation. We also observed a negative correlation between EV-D68 and wheezing in adults ≥65 years and no correlation in children ≤5 years. Wheezing is a nonspecific symptom caused by multiple respiratory pathogens with distinct seasonality, which likely masks any EV-D68-specific signal.

Limitations of the study include the aggregation of environmental variables as weekly averages corresponding to the EV-D68 center of season of each WWTP, which may have masked short-term variability and local microclimatic effects. Center of season and duration were derived from smoothed wastewater concentration series, and alternative smoothing or aggregation approaches could yield slightly different results. Demographic variables were often available only at the county level, meaning that the same values were assigned to multiple WWTPs, including some in large or heterogeneous counties. Clinical data from Epic Cosmos were masked for counts ≤10 and were incomplete for several states, reducing comparability with wastewater data.

Our findings show that environmental factors, particularly temperature and dew point, shape the timing of EV-D68 circulation in wastewater, whereas sociodemographic factors determine its duration. Exploring the mechanisms that mediate the association between dew point or temperature and EV-D68 virus would be beneficial to further analyze different intervention strategies. The extended season observed in California suggests that EV-D68 seasonality may be shifting and could become less predictably biennial under changing climatic conditions. Although no vaccine exists, targeted infection-control measures, especially in childcare settings where transmission amplifies, would help mitigate spread once community circulation is detected. The scarcity of clinical data, due to limited EV-D68 testing and the rarity of severe outcomes such as AFM, highlights the value of wastewater surveillance as an independent and timely indicator of EV-D68 circulation. Because wastewater data are specific and available within 24 hours of sampling, they can provide early warning for public health response, including alerts to clinicians for potential increases in severe respiratory illnesses caused by EV-D68 infections. Nevertheless, while wastewater reflects community-level infection dynamics, it cannot distinguish symptomatic from asymptomatic infections; dedicated EV-D68 shedding studies could help estimate infection prevalence by back-calculating from wastewater concentrations.

## Materials and Methods

### Ethical approval

This study was reviewed by the Stanford University Institutional Review Board and was determined not to involve human subjects as defined under 45 CFR 46.102(e), nor to constitute research or clinical investigation as defined under 45 CFR 46.102(l) or 21 CFR 50.3(c).

### Wastewater data collection

Enterovirus D68 (EV-D68) RNA was quantified in wastewater solids from 147 wastewater treatment plants (WWTPs) across 40 states in the United States between July 2023 and July 2025 (Figure 6). Samples were collected 1 to 7 times per week at each WWTP (median = 3) (Table S1) and analyzed by droplet digital RT-PCR (ddRT-PCR) as previously described (7,23). Full methods are provided in peer reviewed data descriptors (23).

**Figure 6.**
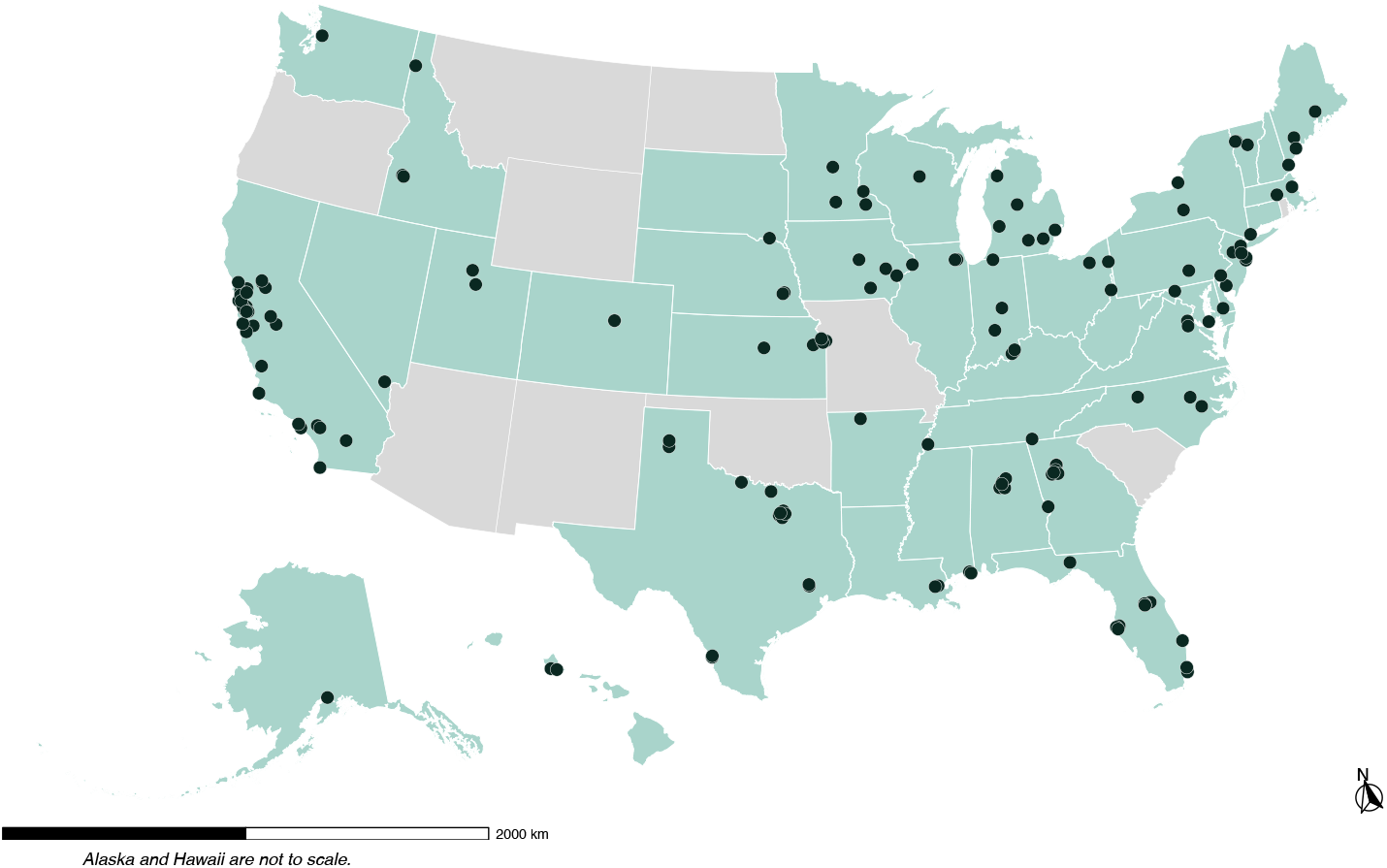
Locations of the 147 wastewater treatment plants (WWTPs) where samples were collected in this study (July 2023–July 2025). States with enrolled sites are shaded in light green; black dots mark individual plants. Alaska and Hawaii are not to scale.

### Wastewater variables

Two forms of EV-D68 RNA concentration measurements in wastewater solids were used for different analyses in this study. First, EV-D68 RNA concentrations were normalized to concentrations of pepper mild mottle virus (PMMoV) RNA to account for variations in fecal load and to correct for differences in viral RNA isolation efficiency (24,25). Second, a 5-day centered trimmed moving average was applied to daily PMMoV-normalized concentrations (referred to hereafter as the smoothed, PMMoV-normalized EV-D68 concentration). Within each 5-day window, the highest and lowest values were excluded, and the remaining three values averaged.

All wastewater data were aggregated at the weekly level, yielding a single concentration value per epidemiological week. Epidemiological weeks were defined according to the ISO 8601 standard as calendar weeks beginning on Monday and ending on Sunday. Weekly aggregation was first performed at the WWTP level by calculating the arithmetic mean of all samples collected within the same epidemiological week for each WWTP.

For state- and national-level aggregation, WWTP-level weekly concentrations were combined using a population-weighted mean based on WWTPs within each state, and population serving each WWTP, to obtain state-level weekly average concentrations. For national-level aggregation all 147 WWTPs were used to obtain weekly averaged concentrations for each epidemiological week over the study period. In some analyses, concentrations were further aggregated to the monthly level by calculating the arithmetic mean of concentrations from all epidemiological weeks falling within each calendar month, at either the state or national level.

### Wastewater-derived variables

Two variables were derived from the smoothed, PMMoV-normalized EV-D68 concentration data to characterize the temporal dynamics of EV-D68 detection in wastewater solids: the EV-D68 center of season and EV-D68 season duration. Smoothed, PMMoV-normalized EV-D68 RNA concentrations were used in preference to raw concentrations or PMMoV-normalized-only concentrations because they have less high-frequency noise (Figure S9).

The center of season was defined as the average week of EV-D68 detection, indicating the midpoint of the seasonal activity period based on the timing of detections. The specific data used for this calculation varied by spatial scale. At the state and national levels, the center of season was calculated using epidemiological week-level concentrations, previously aggregated across WWTPs as described above. A single reference base was set to the earliest Monday in the analysis window, and a global, monotonic week index was defined for each epidemiological week (e.g., the first week in the dataset was assigned index 1, the second week index 2, and so on) so that time increased continuously without wraparound at New Year (26,27). Each indexed week was then multiplied by its corresponding EV-D68 concentration (i.e., its weight). Weeks with no detectable concentration were assigned a weight of zero and did not contribute to the total. All products (index × concentration) were summed and divided by the total sum of concentrations across all weeks. The result was the weighted average of week indices, which defined the center of season. For interpretation, the center of season was mapped back to calendar time and rounded to the nearest week for display (Supplementary Information 1). To compute the center of season at the WWTP level, all individual daily sample measurements were used directly, without aggregation at epidemiological week. Each sample was assigned its corresponding index, and the calculation followed the same weighted average approach described above.

Season duration was defined as the number of epidemiological weeks during which EV-D68 RNA concentrations in wastewater solids remained elevated, providing a measure of the temporal extent of viral activity. Two different approaches were used to calculate season duration, depending on the spatial scale of analysis. At the WWTP and state levels, season duration was defined as the span of two or more consecutive “detectable weeks” that included the previously calculated center of season. A “detectable week” was defined as a week in which at least one sample had an EV-D68 RNA concentration above the assay detection limit (28). At the WWTP level, although the center of season was calculated using daily concentrations, season duration was computed at the weekly level. Daily samples were grouped by epidemiological week, and any week with at least one sample showing detectable EV-D68 RNA concentration (i.e., above the detection limit) was classified as “detectable”. At the national level, a “detection”-based threshold was unsuitable because the aggregation of data from 147 WWTPs produced a continuous low-level signal, even during periods of minimal viral concentration. To isolate periods of elevated viral concentrations, a baseline threshold was defined as the mean of the lowest tertile in the national smoothed PMMoV-normalized EV-D68 concentration aggregated by epidemiological week, plus three standard deviations. The national EV-D68 season duration was then defined as the number of weeks in which concentrations exceeded this threshold and included the center of season.

### Environmental variables

Environmental variables included daily measurements of average temperature, dew point, relative humidity, precipitation, wind speed, and surface pressure. These data were retrieved from NOAA via a custom Python script (accessed September 3, 2025). For each WWTP, environmental data were drawn from the nearest weather station; in total, 117 stations were used, each representing 1 - 5 WWTPs (Table S7). Daily environmental values were aggregated at the level of each weather station in two ways. First, daily measurements were averaged into weekly means based on epidemiological weeks to align with the temporal resolution of the weekly averaged EV-D68 concentration data described above. Second, we calculated the mean of daily environmental values across the EV-D68 season duration, as previously defined for each WWTP. This provided two environmental summaries per WWTP: one for the specific week of the center of season, and one for the entire season duration.

### Sociodemographic and structural variables

Sociodemographic and structural determinants included the proportion of children aged ≤5 years, the proportion of adults aged ≥65 years, household crowding (defined as the percentage of households with more than one person per room), birth rate, density of childcare services, number of hospitals, number of nursing homes, number of airports, population density, and urbanicity (Table S8).

Demographic and housing variables were derived from the 2023 5-year American Community Survey (ACS) at the census tract level (29). Because sewershed and census tract boundaries were not aligned, tract-level counts were aggregated to the sewershed level by calculating the proportion of each tract intersecting a given sewershed using ArcGIS Pro (version 3.1.1), as previously described (30). Counts were adjusted by these area proportions and summed to generate sewershed-level values. Birth rate (number of births per capita) was calculated at the sewershed level using 2024 US Census Bureau Population Estimates (31). County-level birth counts and population denominators were aggregated to sewersheds using the same weighted spatial procedure as for ACS data, but intersecting with counties instead of census tracts (30).

Childcare establishment density was calculated using 2023 County Business Patterns data (32), based on the number of childcare service businesses in each county (NAICS code 624410), divided by the land area of the county in square kilometers. Land area was calculated using 2022 US Census Bureau county boundaries (5-metre resolution) (33). Each sewershed was assigned the density value of its predominant county (i.e., the county containing the largest share of the sewershed area). Population density was similarly defined as the number of residents per square kilometer in the predominant county of each sewershed, using 2022 ACS population estimates (34) and county land area data from the US Census (33). Urbanicity was calculated as the proportion of each sewershed intersecting urban-designated areas, based on 2020 US Census Bureau Urban-Rural Classification data (35). Hospitals, nursing homes, and airports per sewershed were retrieved from a previous study (30).

### Clinical diagnoses variables

Clinical diagnostic records were obtained from Epic Cosmos (https://cosmos.epic.com/, accessed September 10, 2025) for the period July 2023–July 2025 and extracted at two geographic resolutions: national and U.S. state. Four diagnostic categories were analyzed: acute flaccid myelitis (AFM) (ICD-10-CM G04.82), wheezing in patients ≤ 5 years old (ICD-10-CM R06.2), wheezing in patients ≥ 65 years old, and enterovirus-specific diagnoses (ICD-10-CM B34.1, B97.10, B97.19). Counts of 10 or fewer are set to “zero” to minimize potential reidentification risk resulting in a “zero” value for the calculated rate for a given period. For both jurisdictions (state and national) and for each week, a diagnosis proportion was calculated as the number of encounters with the diagnosis divided by the total number of encounters in that jurisdiction-week. An exception was applied to AFM at the state level: AFM data were retrieved and analyzed monthly rather than weekly because weekly state counts were frequently small and masked. Monthly AFM proportions were calculated as the monthly AFM count divided by the total number of encounters in that state-month.

### Statistical analyses

All statistical analyses and visualizations were conducted in R version 4.5.1 using RStudio version 2025.09.0.

Latitude and longitude of each WWTP were used to evaluate spatial gradients in the timing and duration of EV-D68 detection in wastewater solids. Associations between geographic location (latitude and longitude) and two outcome variables, EV-D68 center of season and season duration, were assessed using four separate weighted linear regression models. In each model, latitude or longitude served as the independent variable, and either the center of season or season duration as the dependent variable. To account for variability in EV-D68 concentration and detection frequency across WWTPs, each WWTP g was assigned a regression weight *W*_*g*_, calculated as the cumulative smoothed, PMMoV-normalized EV-D68 RNA concentrations across all samples collected at that WWTP:

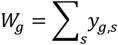

where *y*_*g,s*_ denotes the smoothed, PMMoV-normalized EV-D68 RNA concentration for sample *s* at WWTP *g*, with non-detects assigned a value of zero. This weighting approach gave greater influence to WWTPs with more frequent and/or higher concentrations of EV-D68 RNA over time. One WWTP (Clinton, Iowa) had no detectable smoothed, PMMoV-normalized EV-D68 RNA concentration (W=0) and was excluded from the analysis, resulting in a final sample of 146 WWTPs. All regression models were implemented using weighted least squares and evaluated independently for center of season and season duration.

Associations between environmental variables and EV-D68 center of season were evaluated at the WWTP level using weighted least-squares regression. The dependent variable was the EV-D68 center of season, defined as the epidemiological week number when detections were more concentrated at each WWTP. The predictor was the mean value of each environmental variable during the corresponding epidemiological week of the center of season for that WWTP. The same WWTP-level regression weights described above (*W*_*g*_), based on cumulative smoothed, PMMoV-normalized EV-D68 concentrations, were used to account for variation in detection intensity across sites. Because meteorological variables are often correlated with one another and with geographic location, particularly latitude and longitude (Figure S2), each environmental variable was analyzed in a separate regression model to avoid multicollinearity. Two model specifications were fitted for each variable. The first included the environmental variable alone. The second included the environmental variable along with latitude and longitude as predictors to adjust for geographic confounding. A third, geography-only model including only latitude and longitude was used as a baseline for comparison. Model performance was summarized using the weighted coefficient of determination (*R*_*w*_^*2*^), and differences in *R*_*w*_^*2*^relative to the geography-only baseline model were used to evaluate the added explanatory value of each environmental variable. A priori, meteorological variables were not included as predictors of EVD-68 season duration, on epidemiological grounds that duration is more plausibly driven by demographic structure and healthcare seeking behavior rather than short-term weather. However, as a robustness check, duration models were refitted with meteorological covariates: temperature, dew point, relative humidity, precipitation, wind speed, and surface pressure (Supplementary Information 2).

Sociodemographic and structural variables were evaluated for their association with EV-D68 season duration using non-parametric statistical methods. To assess whether season duration differed across categories of each variable, Kruskal–Wallis tests were applied. For variables with more than two categories, such as tertiles of household crowding, childcare density, and population density, pairwise Wilcoxon rank-sum tests were performed following a significant Kruskal–Wallis result, with Bonferroni correction for multiple comparisons. For binary variables, including urbanicity, presence of an airport, and presence of hospitals or nursing homes, Wilcoxon rank-sum tests were used without adjustment. As a robustness check, associations between EV-D68 center of season and sociodemographic and structural variables were also evaluated using the same non-parametric statistical approach applied to season duration.

Correlations between clinical diagnoses and unsmoothed, PMMoV-normalized EV-D68 RNA concentrations in wastewater solids were assessed at both the state and national levels. At the national level, Spearman rank correlation analyses were conducted using epidemiological week-level data for four diagnoses: wheezing in children aged ≤5 years, wheezing in adults aged ≥65 years, enterovirus, and AFM. At the state level, weekly correlations were performed for wheezing (both age groups) and enterovirus diagnoses, while AFM correlations were assessed at the monthly level due to its lower frequency of reporting. All time series were restricted to the EV-D68 season duration as defined from wastewater data, with an extension of ±2 epidemiological weeks for weekly series and ±2 calendar months for monthly series to account for potential reporting delays. Correlation results were summarized using Spearman rank coefficients, and statistical significance was assessed using Bonferroni-adjusted p-values to correct for multiple testing. Findings were visualized with lollipop plots by state and diagnosis.

## Data availability

All data used in this study, along with the Python scripts and R code for statistical analyses, are publicly available at the Stanford Digital Repository: https://purl.stanford.edu/xz608ws5349.

## Acknowledgments

We thank the participating wastewater treatment plants for their samples for the project, and we are grateful to Elana Chan for her help in retrieving sociodemographic variables. This work was supported by a gift to ABB from the Sergey Brin Family Foundation. Clinical diagnoses data used in this study were retrieved from Epic Cosmos (September 10, 2025), a dataset created in collaboration with a community of Epic health systems representing more than 284 million patient records from over 1,500 hospitals and 36,000 clinics from all 50 states, D.C., Lebanon, and Saudi Arabia.

## Author Contributions

S.C., A.Z., and A.B. conceptualized the study. S.C. and A.Z. curated the data. S.C. performed the formal analysis, developed the methodology, and generated the visualizations. A.B. acquired funding, administered the project, provided resources and supervision, and performed validation. S.C., A.Z., and A.B. conducted the investigation. S.C. wrote the original draft of the manuscript. S.C., A.B., and A.Z. reviewed and edited the manuscript.

