## Supplementary Information for "Enterovirus D68 in United States wastewater is associated with climatic and demographic factors: a comparison with clinical diagnoses"

### Supporting Information Text

#### 1. Calculation of EV-D68 center of season

Seasonal timing was summarized as the mean epidemic week, with each week weighted by EV-D68 signal, using an unwrapped week index across years so that time increases continuously without wraparound at New Year (1,2). This approach is appropriate for EV-D68, whose activity peaks in late summer–early autumn and does not straddle December-January.

A single reference Monday ("base") was set to the earliest Monday in the analysis window. For each epidemiological week *i* (defined by its Monday date $date_i$), a global, monotonic week index was defined as

$$t_i = 1 + \frac{date_i - base}{7},$$

so that indices increase monotonically across calendar years.

Weekly weights $w_i$ were taken as the normalized, 5-day–trimmed concentration for that week, with non-detects set to zero.

The mean epidemic week (center) and its spread (standard deviation), both on the global week scale, were computed as

$$\mu = \frac{\sum_i w_i t_i}{\sum_i w_i}, \qquad \sigma = \sqrt{\frac{\sum_i w_i (t_i - \mu)^2}{\sum_i w_i}}.$$

If $\sum_i w_i = 0$, the center was undefined, and the series was excluded from timing analyses. For interpretation, the center was mapped back to calendar time as

$$\text{mean date} = \text{base} + 7\mu$$

For discrete reporting by epidemiological week, μ was rounded to the nearest integer before mapping; for fractional centers, rounding was omitted.

#### 2. Environmental variables effect on EV-D68 season duration

To evaluate associations between environmental variables and EV-D68 season duration, we used the same environmental summaries defined in the main text. Specifically, the mean value of each variable across the full EV-D68 season duration at each WWTP. For each variable, univariate WWTP-level weighted least-squares regressions were fitted with season duration as the dependent variable. Weights (W) were defined as the inverse of the squared "within-season duration" standard deviation for each environmental variable (W=1/SD2), giving greater weight to WWTPs with more stable environmental conditions. As no association was observed between environmental variables and EV-D68 season duration, models were left unadjusted for geography. Results are shown in Figure S4 and Table S3.

#### 3. Specificity of ddPCR assay primers and probes in 2025

During January-July 2025, when wastewater detections of EV-D68 were minimal, NCBI Nucleotide (nuccore) was queried with a Python script on August 25, 2025, for EV-D68 VP1 sequences released in 2025 using the keywords "evd68 AND vp1." Records were retained only if annotated with a 2025 collection date. No sequences met this criterion; therefore, in-silico primer–probe matching was not performed.

### Figures

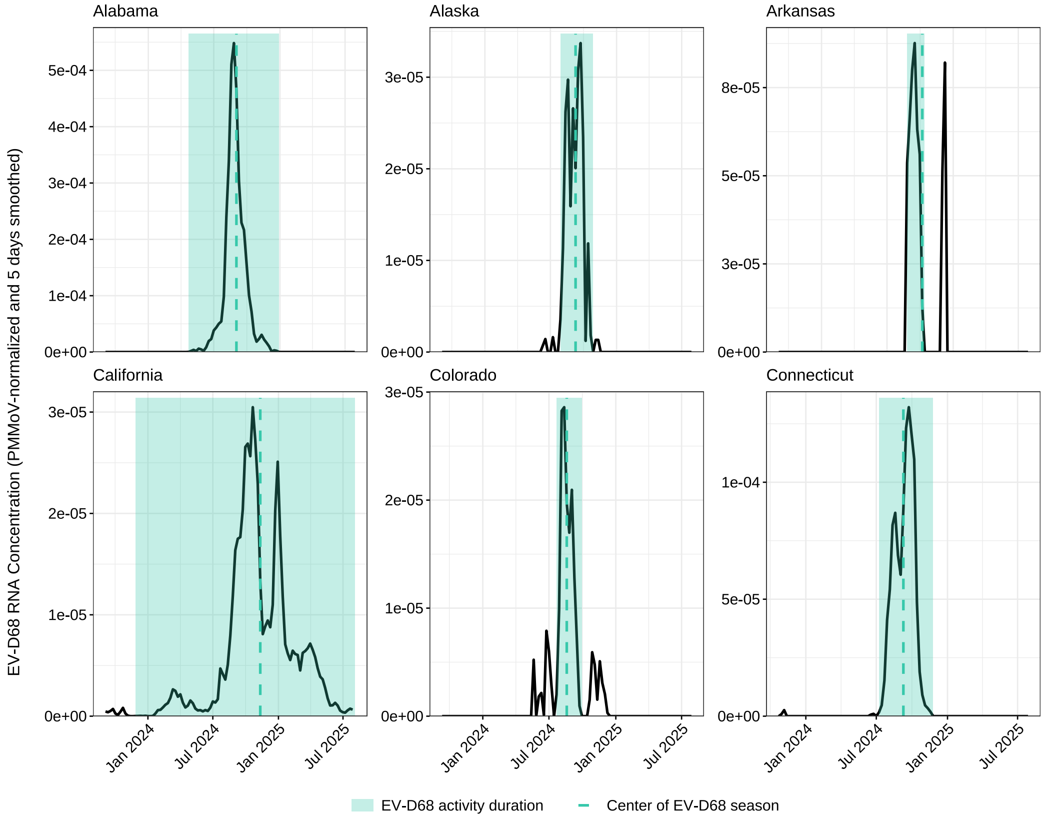
Alabama
Alaska
Arkansas
California
Colorado
Connecticut
EV-D68 RNA Concentration (PMMoV-normalized and 5 days smoothed)
Jan 2024
Jul 2024
Jan 2025
Jul 2025
EV-D68 activity duration
Center of EV-D68 season

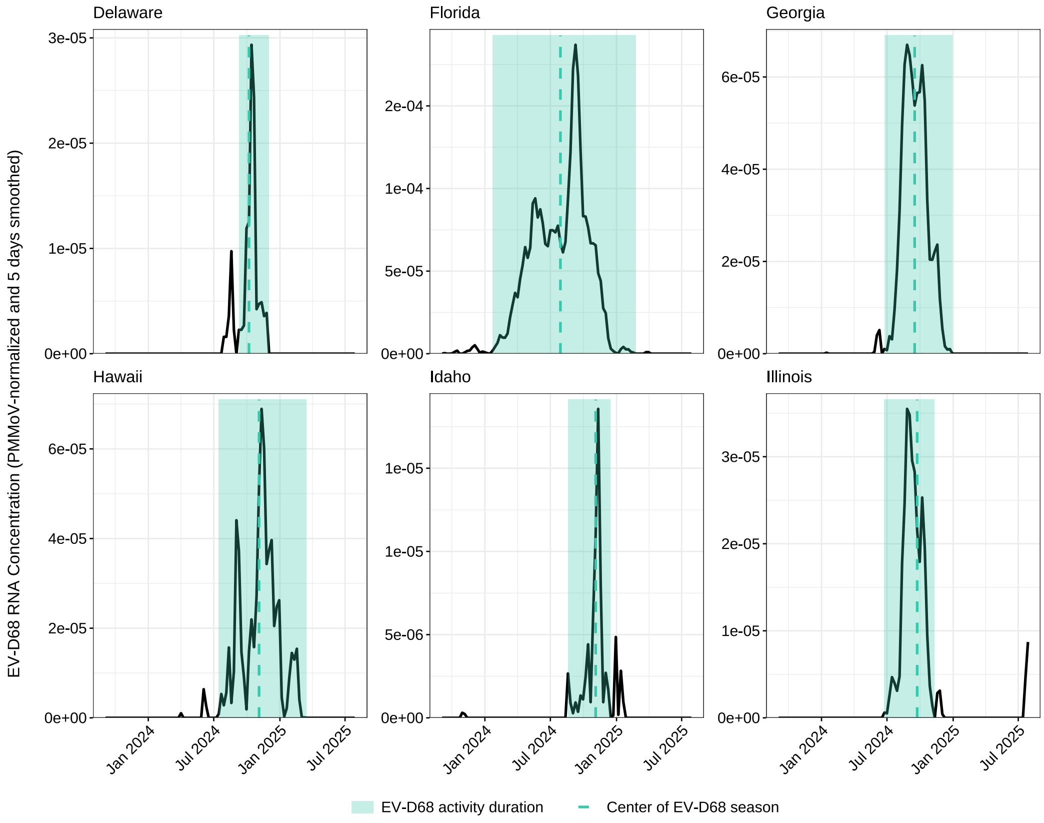
Delaware
Florida
Georgia
Hawaii
Idaho
Illinois
EV-D68 RNA Concentration (PMMoV-normalized and 5 days smoothed)
Jan 2024
Jul 2024
Jan 2025
Jul 2025
EV-D68 activity duration
Center of EV-D68 season

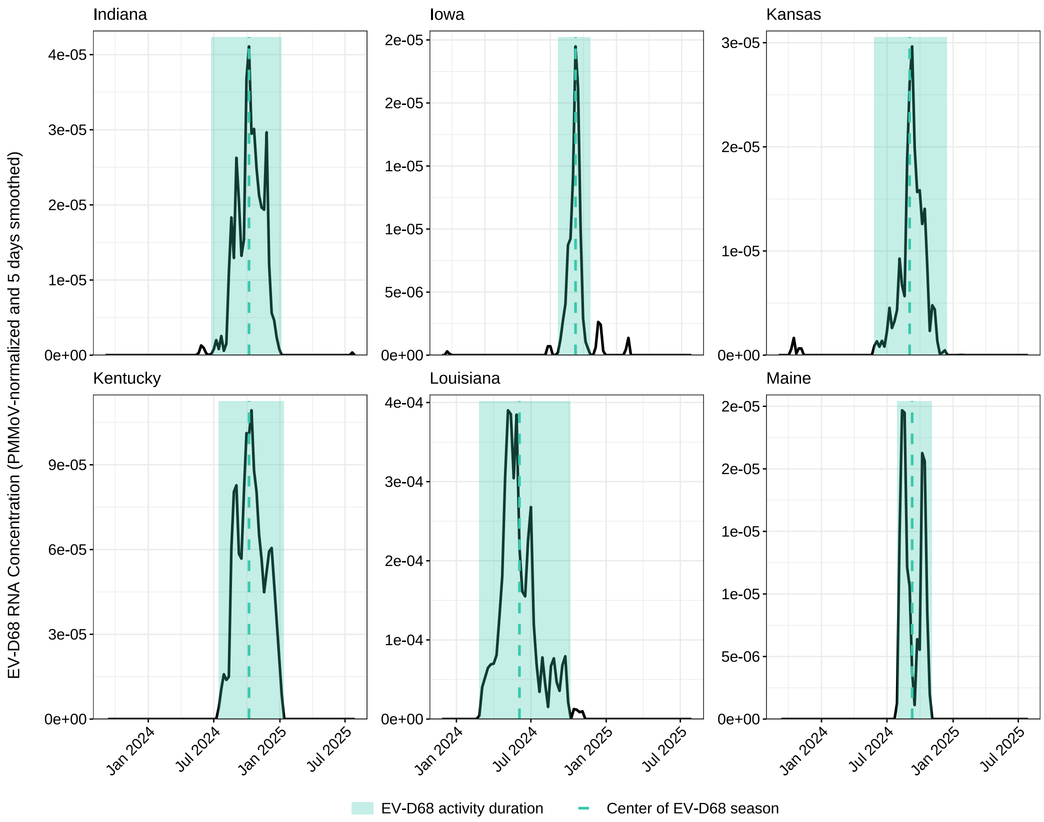
Indiana
Iowa
Kansas
Kentucky
Louisiana
Maine
EV-D68 RNA Concentration (PMMoV-normalized and 5 days smoothed)
Jan 2024
Jul 2024
Jan 2025
Jul 2025
EV-D68 activity duration
Center of EV-D68 season

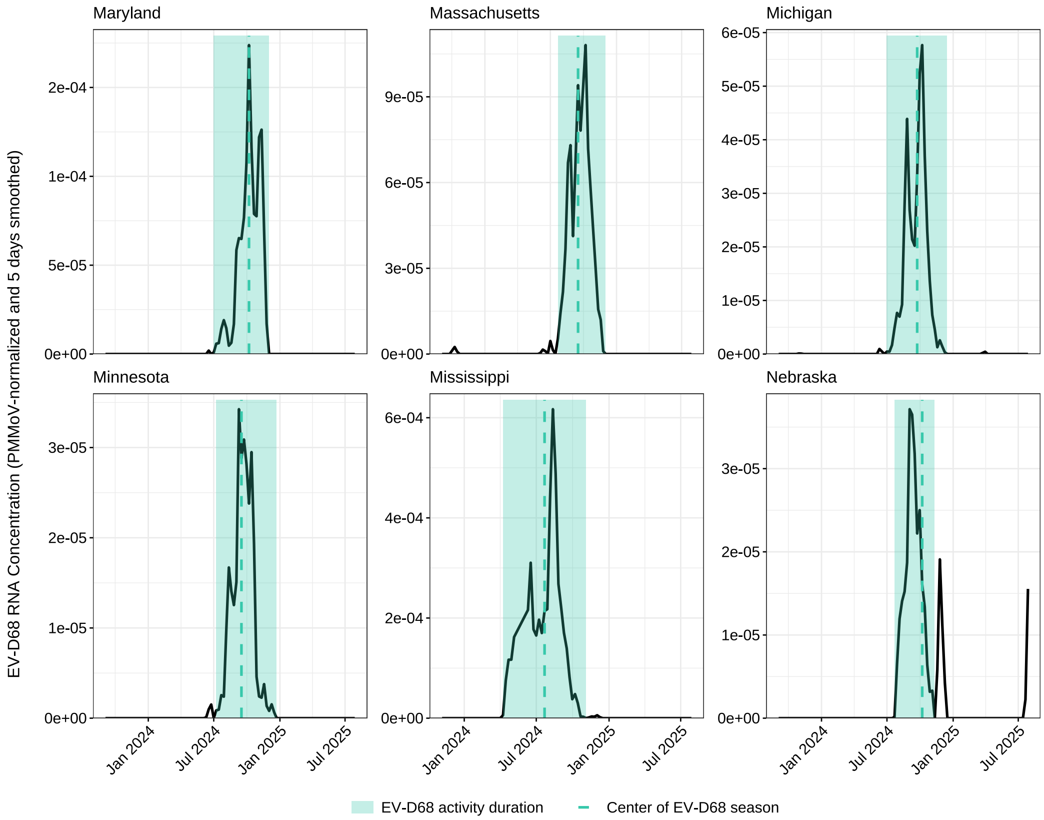
Maryland
Massachusetts
Michigan
Minnesota
Mississippi
Nebraska
EV-D68 RNA Concentration (PMMoV-normalized and 5 days smoothed)
Jan 2024
Jul 2024
Jan 2025
Jul 2025
EV-D68 activity duration
Center of EV-D68 season

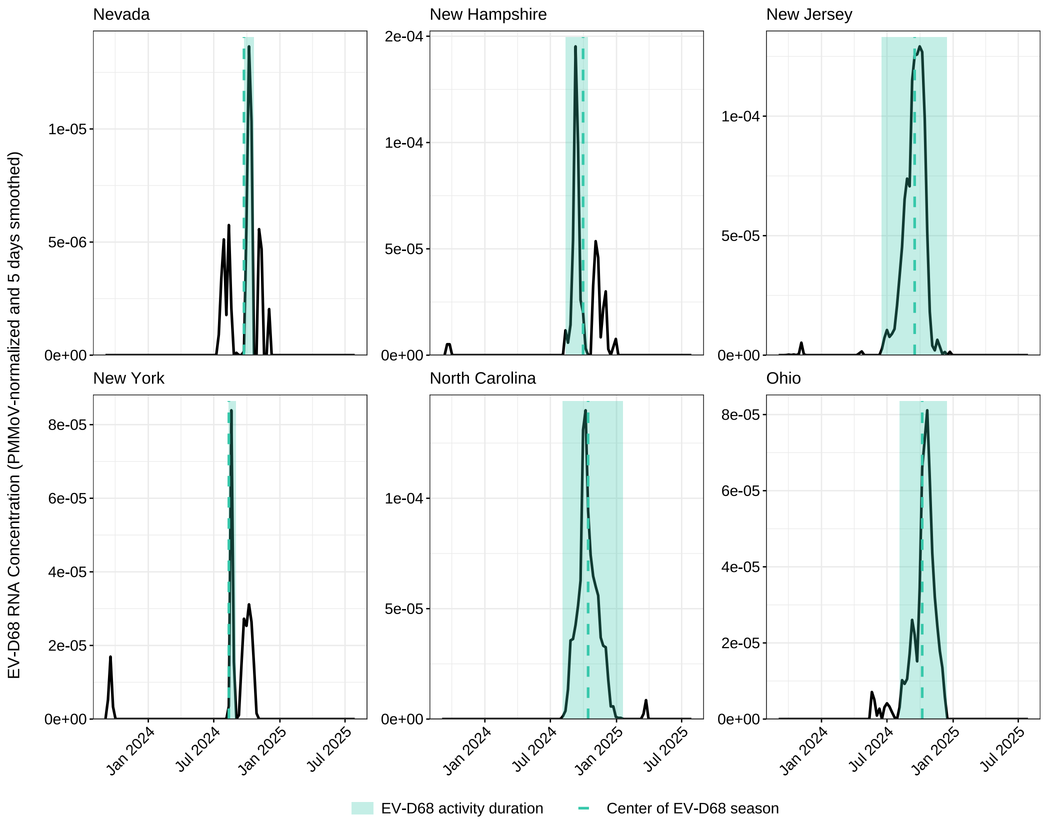
EV-D68 RNA Concentration (PMMoV-normalized and 5 days smoothed)
Nevada
New Hampshire
New Jersey
New York
North Carolina
Ohio
Jan 2024
Jul 2024
Jan 2025
Jul 2025
EV-D68 activity duration
Center of EV-D68 season

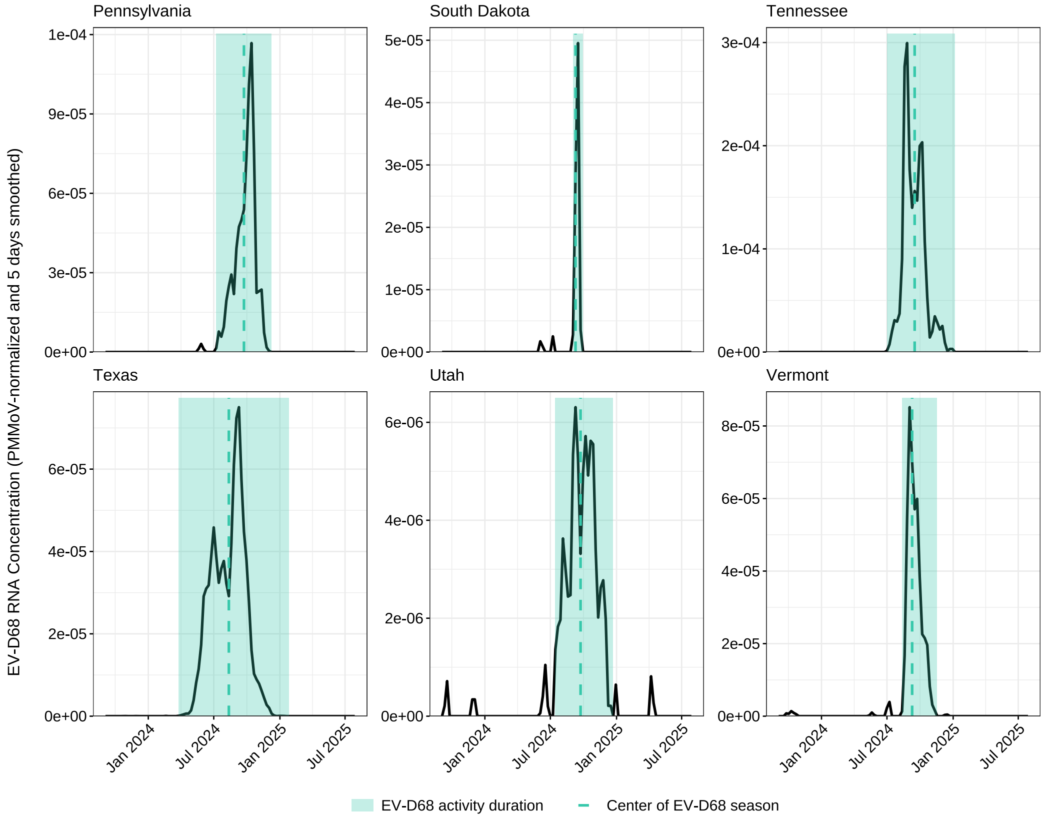
EV-D68 RNA Concentration (PMMoV-normalized and 5 days smoothed)
Pennsylvania
South Dakota
Tennessee
Texas
Utah
Vermont
Jan 2024
Jul 2024
Jan 2025
Jul 2025
EV-D68 activity duration
Center of EV-D68 season

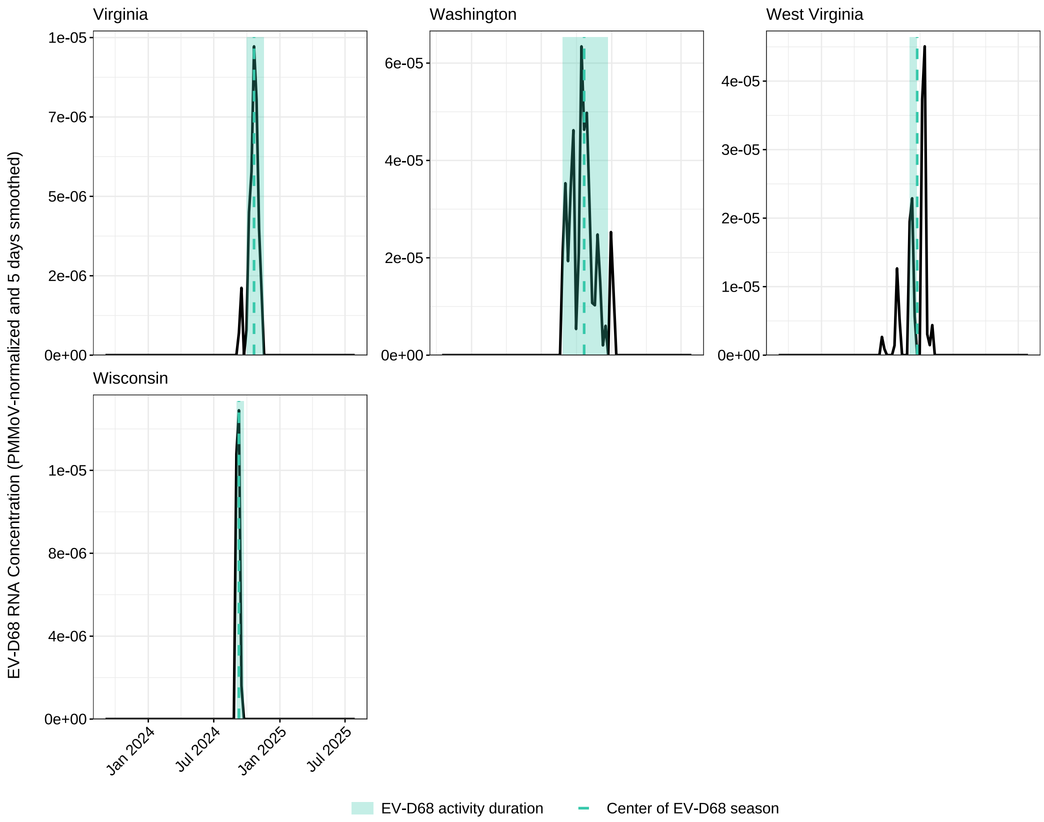


**Fig. S1.** State-level EV-D68 wastewater trends and seasonal timing across 40 U.S. states. Black curves depict weekly state series of smoothed, PMMoV-normalized EV-D68 RNA concentrations. For each wastewater treatment plant (WWTP) and week, smoothed, PMMoV-normalized EV-D68 RNA concentrations measurements were averaged arithmetically; state trends were then obtained as population-weighted means across WWTPs within each state (weights equal to population served). The teal band represents the estimated season duration centered on the center of season, and the dashed teal line denotes the center date.

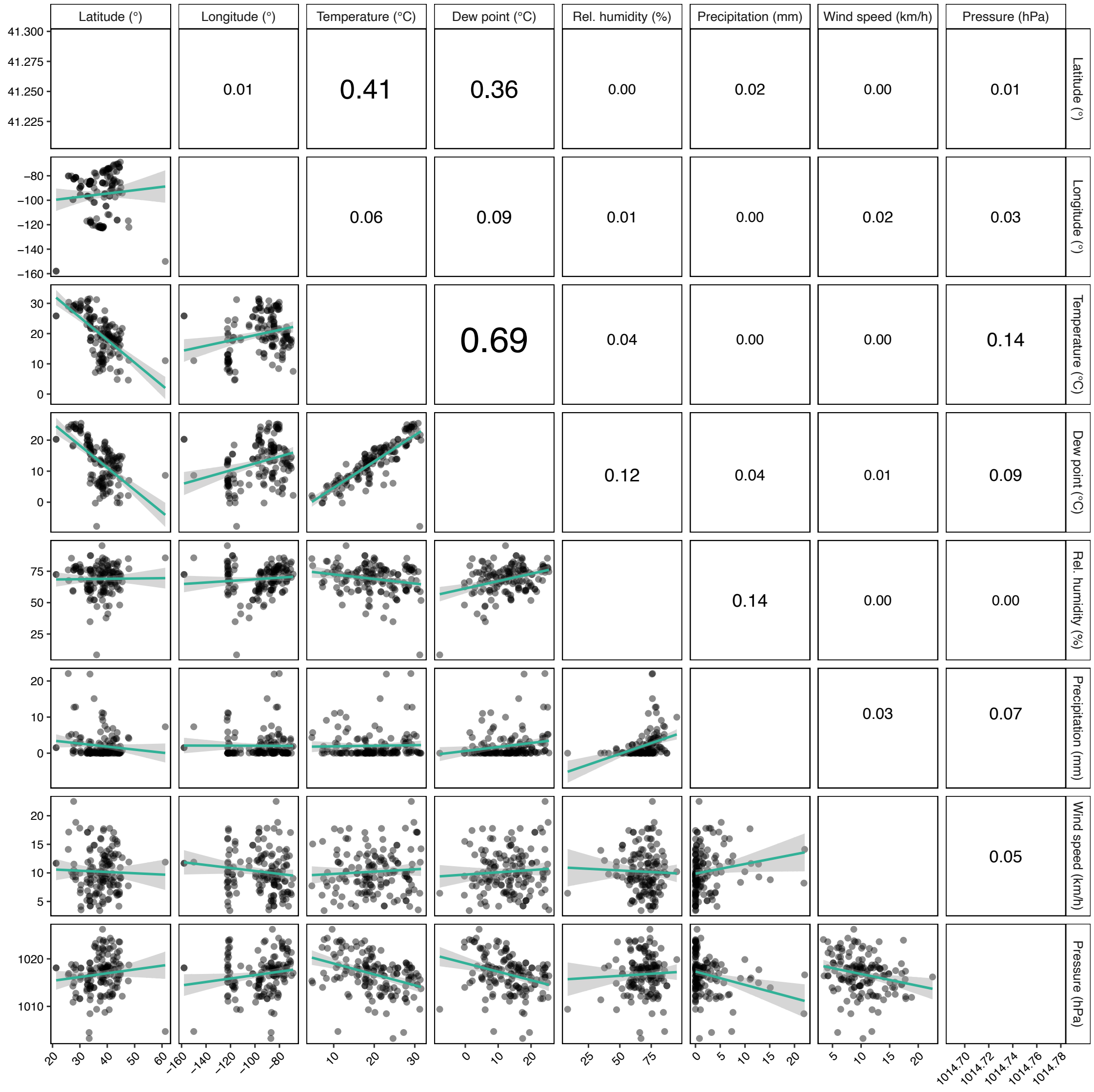


**Fig. S2.** Pairwise collinearity among wastewater treatment plant (WWTP) coordinates and weekly climate at the seasonal center. Each point is a WWTP (n=147). Lower-triangle panels show scatter plots with ordinary least-squares fits (95% CI shading); upper-triangle panels display the corresponding coefficient of determination ($R^2$) for the model *y (row variable) ~ x (column variable)*, with label size proportional to $R^2$. Variables are latitude, longitude, weekly mean temperature, dew point, relative humidity, precipitation, wind speed, and surface pressure. Climate values are the plant-level weekly means for the week containing corresponding to the EV-D68 center of season of the corresponding plant.

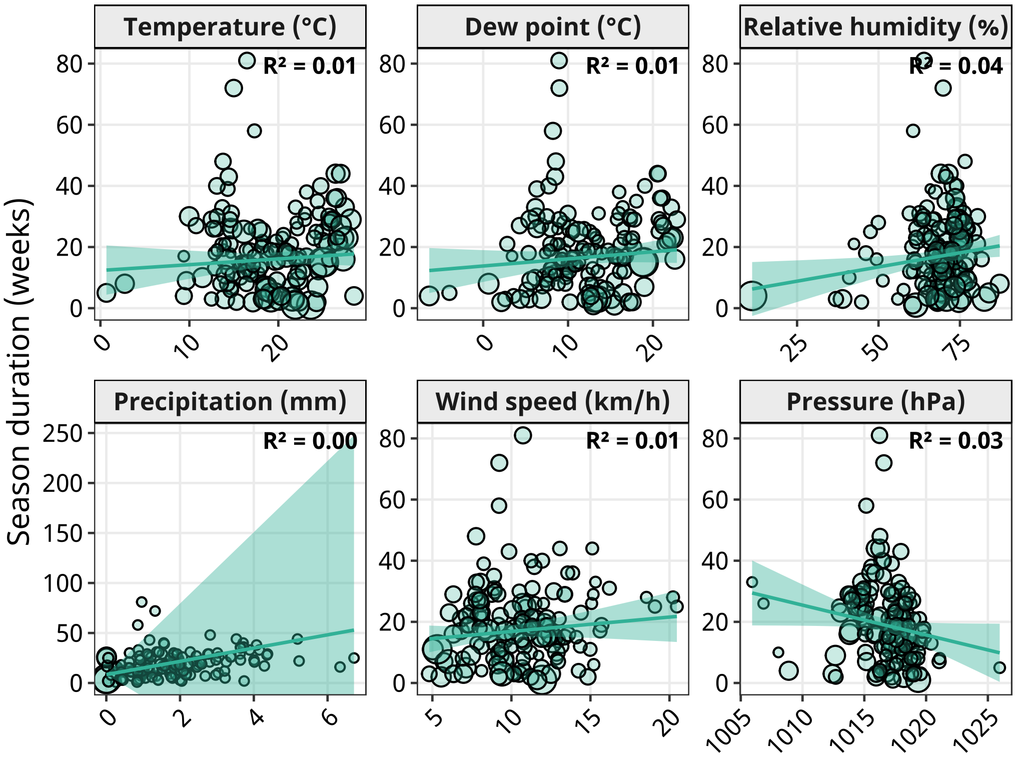


**Fig. S3.** EV-D68 season duration versus mean environmental variables across wastewater treatment plants (WWTPs). For all variables except precipitation, n = 146 WWTPs; for precipitation, n = 144 (two plants had zero within-window standard deviation (SD) and were excluded because inverse-variance weights were undefined). For each plant, the x-axis is the mean of the indicated variable computed from daily values between plant-specific start and end dates; the y-axis is EV-D68 season duration (weeks). Lines show univariate weighted least-squares fits with 95% confidence intervals; weights are the inverse variance of within-window variability ($w = 1/SD^2$). Point sizes are scaled within panels according to weights. The weighted $R^2$ is reported in each panel.

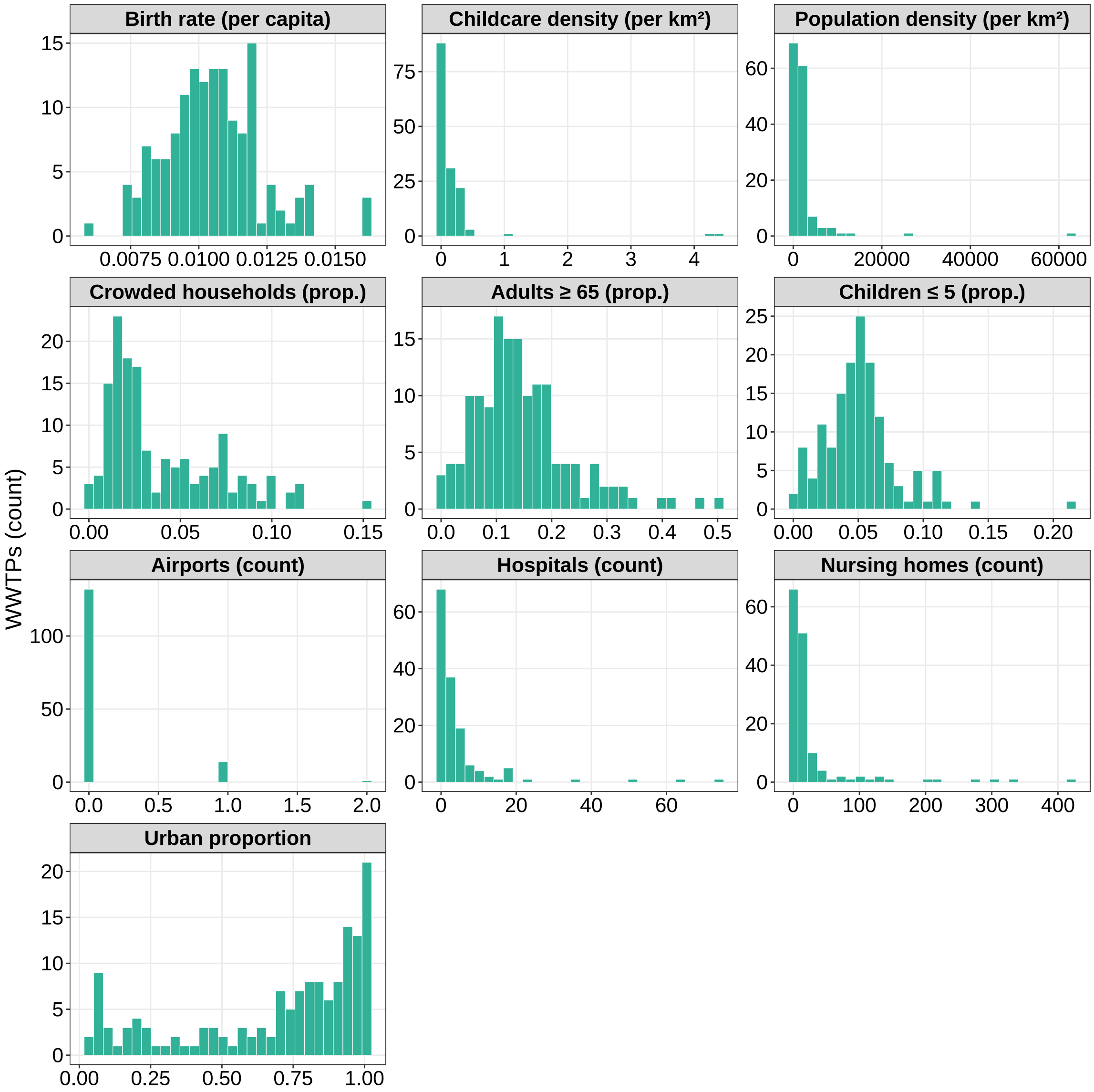


**Fig. S4.** Distribution of sociodemographic characteristics across wastewater treatment plants (WWTPs; n=147). Histograms show the distribution of structural and demographic variables measured at the level of each WWTP catchment. Variables include birth rate (per capita), childcare establishment density (per km²), population density (per km²), proportion of crowded households (defined as number of households with more than one person per room), proportions of adults aged ≥65 years and children aged ≤5 years, counts of airports, hospitals, and nursing homes, and the proportion of the catchment classified as urban.

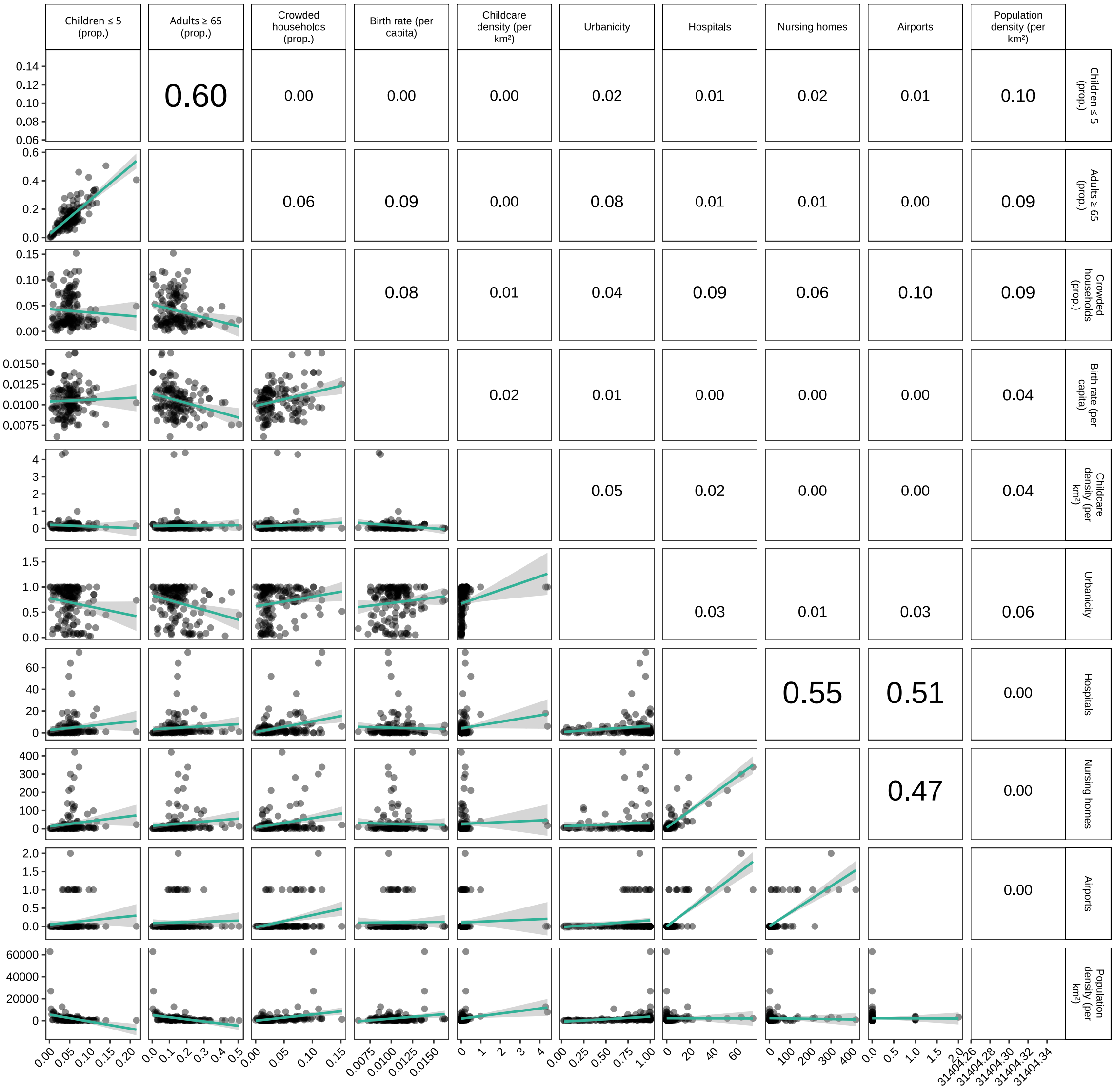


**Fig. S5.** Pairwise correlation between structural and demographic determinants. Scatterplots (lower panels) and coefficients of determination ($R^2$; upper panels) show pairwise associations among sociodemographic variables. $R^2$ values are derived from ordinary least squares regression. Font size of $R^2$ labels corresponds to the strength of association. Diagonal panels show variable names and units. Variables include birth rate (per capita), childcare establishment density (per km²), population density (per km²), proportion of crowded households (defined as number of households with more than one person per room), proportions of adults aged ≥65 years and children aged ≤5 years, counts of airports, hospitals, and nursing homes, and the proportion of the catchment classified as urban.

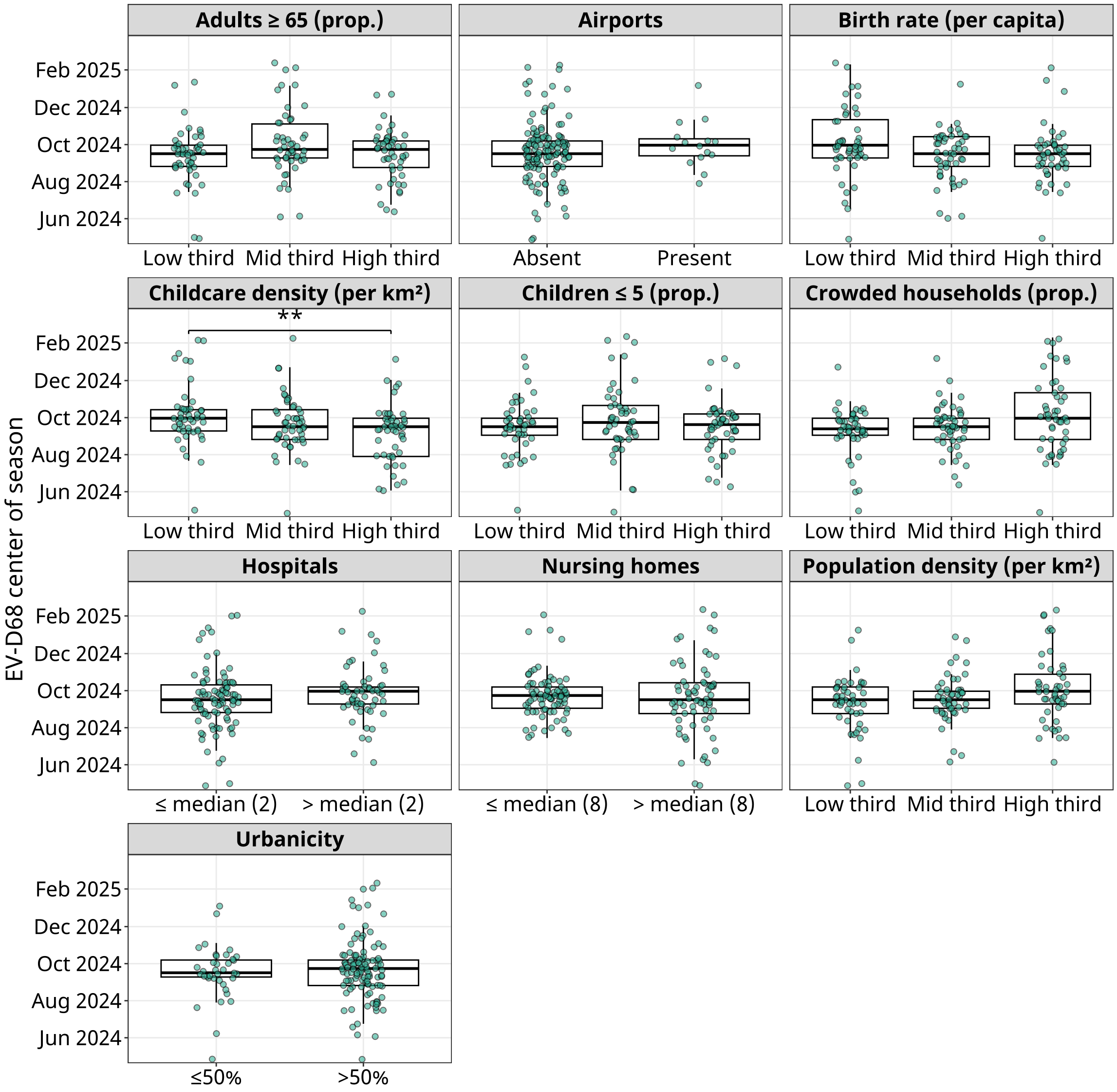


**Fig. S6.** EV-D68 center of season by wastewater treatment plant (WWTP)-level characteristics. Each panel shows the distribution of EV-D68 center of season across categories of a single WWTP–level determinant. Points represent individual WWTPs (n=146) and boxplots show the median, interquartile range (IQR), and whiskers at 1.5×IQR. Airport presence is dichotomised as absent and present. Hospital and nursing home coverage are split using the dataset medians: 2 hospitals and 8 nursing homes within each WWTP catchment area, respectively. Urbanicity is defined by the proportion of the WWTP catchment area classified as urban (≤50% vs >50%). The proportions of children aged ≤5 years, adults aged ≥65 years, crowded households (defined as households with more than one person per room), birth rate (per capita), childcare density (per km²), and population density (per km²) are grouped into within-study tertiles ("low", "middle", "high"). Kruskal–Wallis test was used to assess overall differences, and pairwise comparisons used Wilcoxon rank-sum tests with Bonferroni adjustment. Horizontal connector bars indicate statistically significant differences; asterisks denote adjusted p-values (*p<0.05, **p<0.01, ***p<0.001).

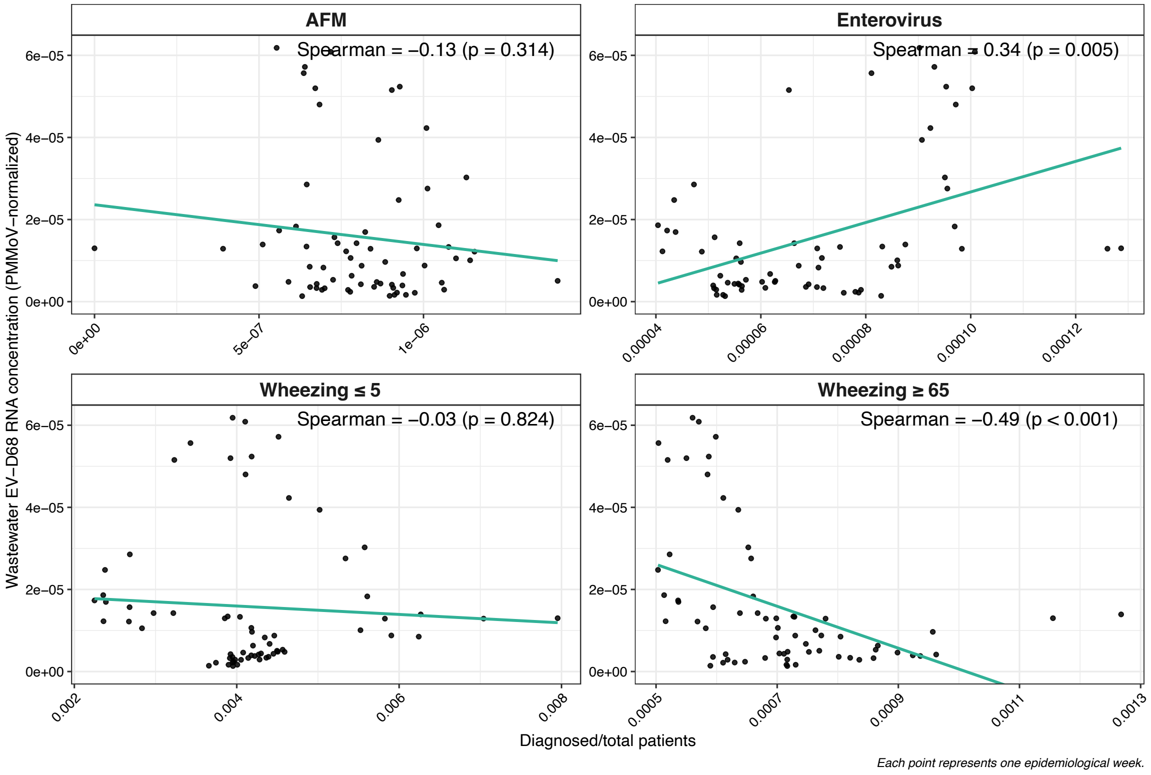


**Fig. S7.** National correlations between wastewater EV-D68 RNA concentration and clinical encounter proportions for selected diagnoses during the EV-D68 season (12 February 2024 to 18 May 2025). Each point represents one epidemiological week. Wastewater concentrations are PMMoV-normalized, aggregated as wastewater plant-week means and population-weighted to the national level. Panels report Spearman rank correlation with p-values. The teal curve shows a linear model fit. Season boundaries were defined from wastewater trends.

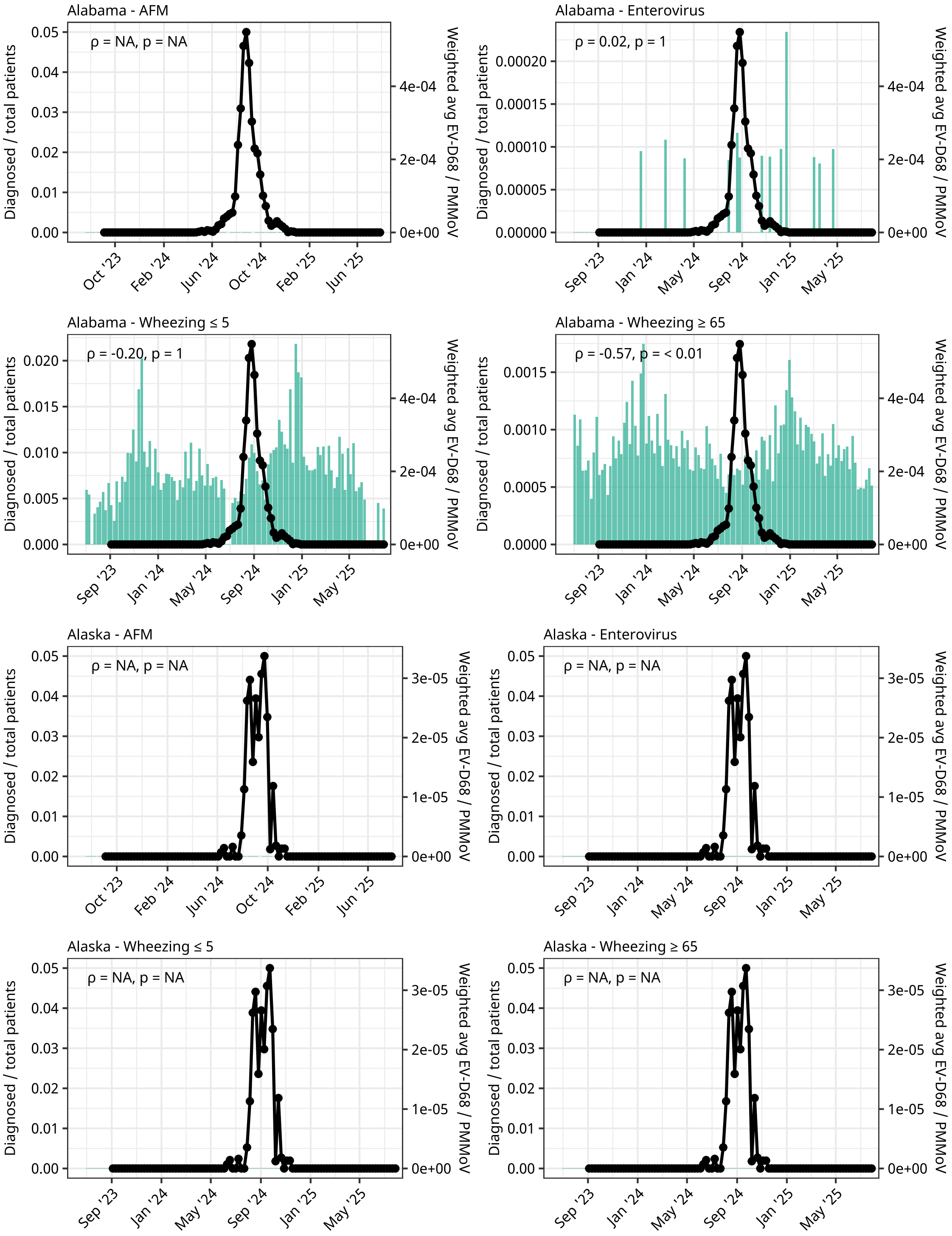
Alabama - AFM
ρ = NA, p = NA
Alabama - Enterovirus
ρ = 0.02, p = 1
Alabama - Wheezing ≤ 5
ρ = -0.20, p = 1
Alabama - Wheezing ≥ 65
ρ = -0.57, p = < 0.01
Alaska - AFM
ρ = NA, p = NA
Alaska - Enterovirus
ρ = NA, p = NA
Alaska - Wheezing ≤ 5
ρ = NA, p = NA
Alaska - Wheezing ≥ 65
ρ = NA, p = NA
Diagnosed / total patients
Weighted avg EV-D68 / PMMoV
Oct '23
Feb '24
Jun '24
Oct '24
Feb '25
Jun '25
Sep '23
Jan '24
May '24
Sep '24
Jan '25
May '25

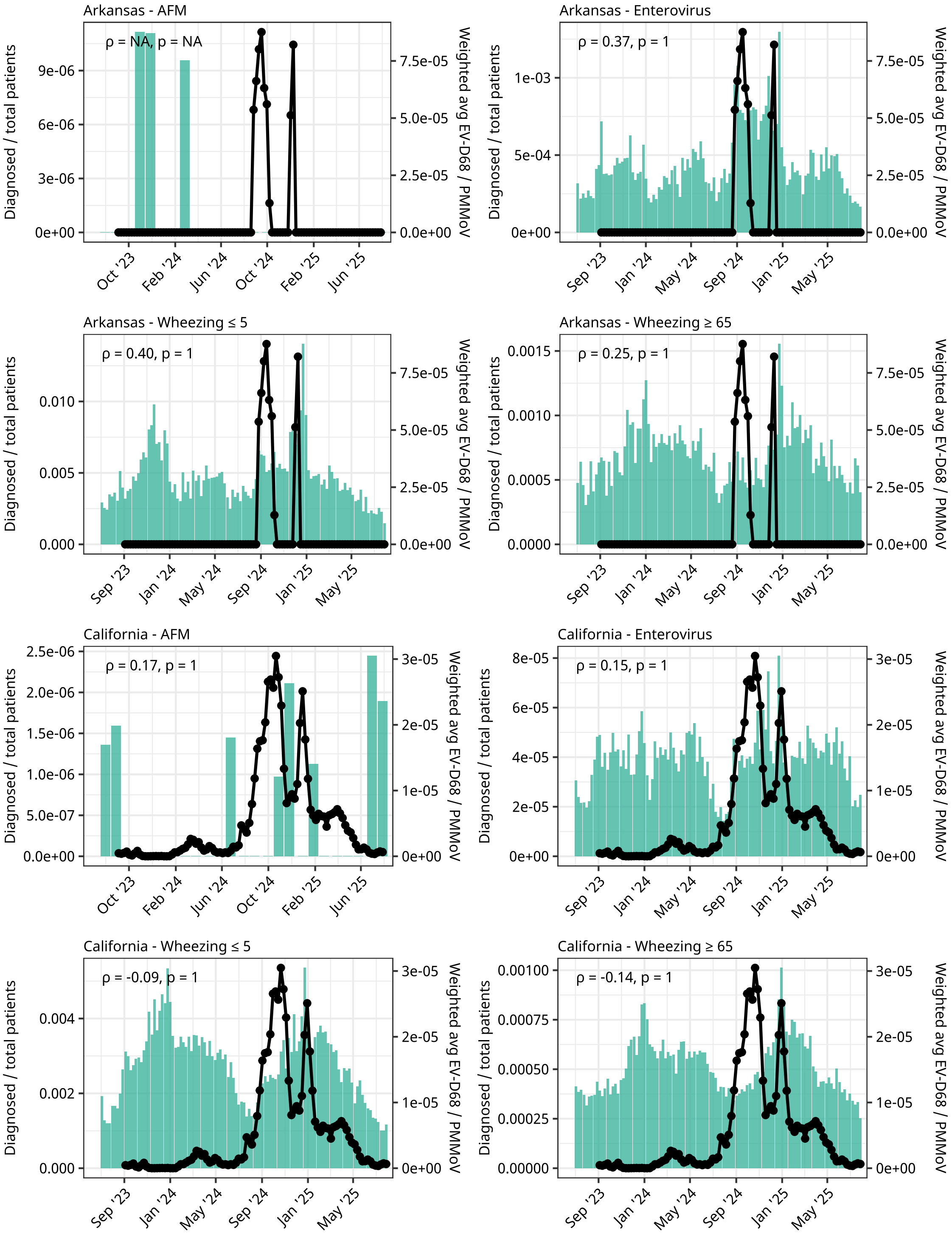
Arkansas - AFM
ρ = NA, p = NA
Arkansas - Enterovirus
ρ = 0.37, p = 1
Arkansas - Wheezing ≤ 5
ρ = 0.40, p = 1
Arkansas - Wheezing ≥ 65
ρ = 0.25, p = 1
California - AFM
ρ = 0.17, p = 1
California - Enterovirus
ρ = 0.15, p = 1
California - Wheezing ≤ 5
ρ = -0.09, p = 1
California - Wheezing ≥ 65
ρ = -0.14, p = 1
Diagnosed / total patients
Weighted avg EV-D68 / PMMoV
Oct '23
Feb '24
Jun '24
Oct '24
Feb '25
Jun '25
Sep '23
Jan '24
May '24
Sep '24
Jan '25
May '25

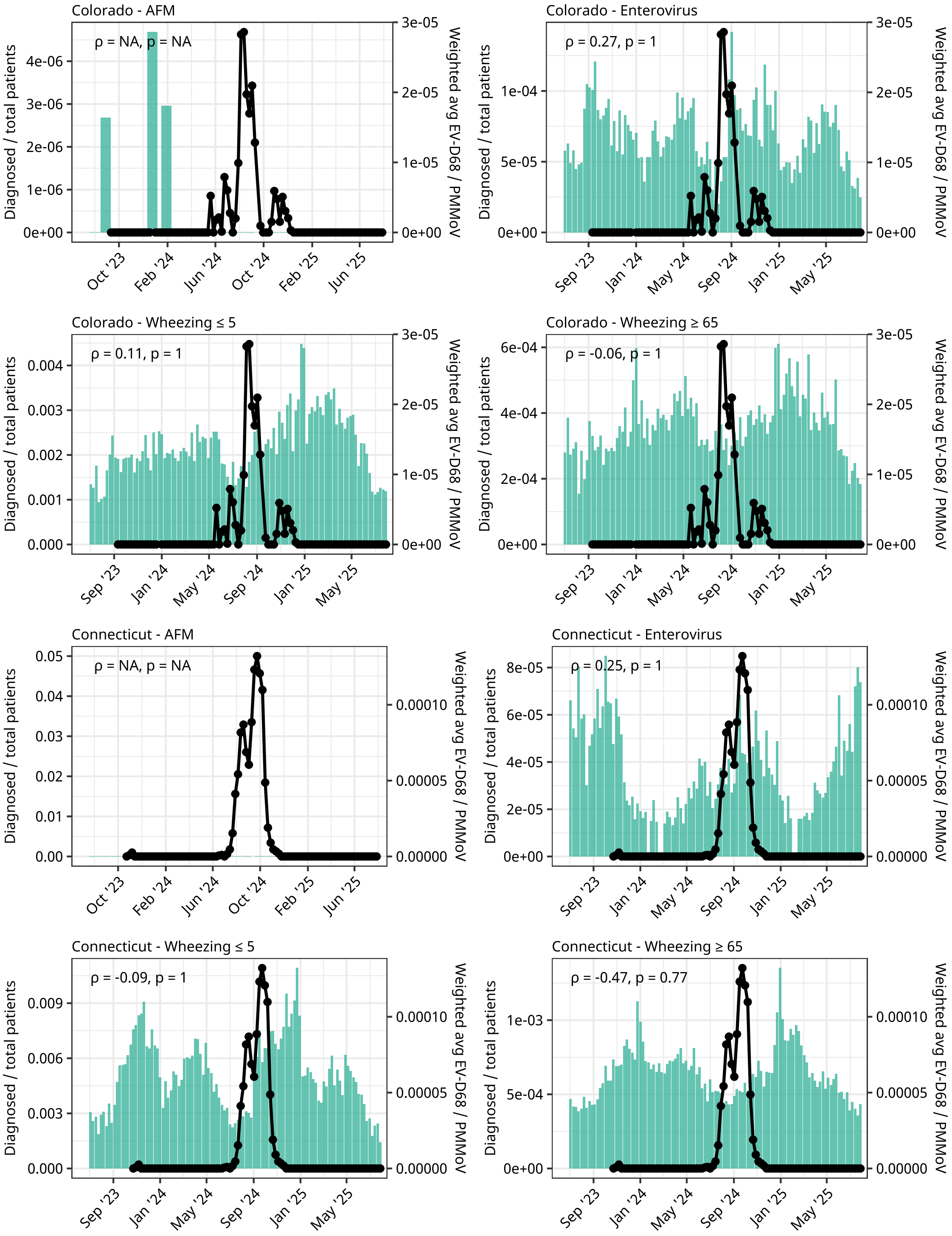
Colorado - AFM
ρ = NA, p = NA
Colorado - Enterovirus
ρ = 0.27, p = 1
Colorado - Wheezing ≤ 5
ρ = 0.11, p = 1
Colorado - Wheezing ≥ 65
ρ = -0.06, p = 1
Connecticut - AFM
ρ = NA, p = NA
Connecticut - Enterovirus
ρ = 0.25, p = 1
Connecticut - Wheezing ≤ 5
ρ = -0.09, p = 1
Connecticut - Wheezing ≥ 65
ρ = -0.47, p = 0.77
Diagnosed / total patients
Weighted avg EV-D68 / PMMoV

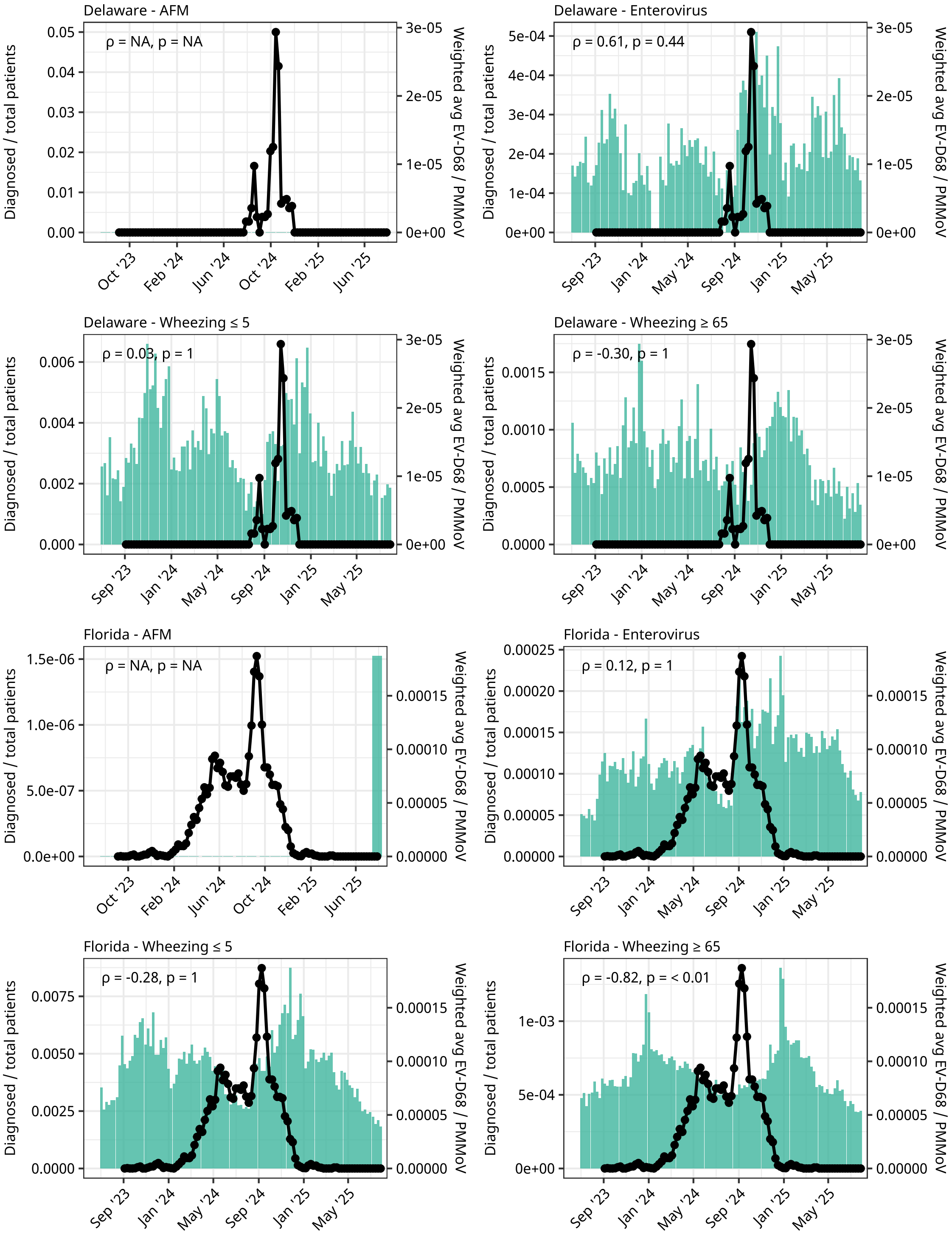

Delaware - AFM
ρ = NA, p = NA
Delaware - Enterovirus
ρ = 0.61, p = 0.44
Delaware - Wheezing ≤ 5
ρ = 0.03, p = 1
Delaware - Wheezing ≥ 65
ρ = -0.30, p = 1
Florida - AFM
ρ = NA, p = NA
Florida - Enterovirus
ρ = 0.12, p = 1
Florida - Wheezing ≤ 5
ρ = -0.28, p = 1
Florida - Wheezing ≥ 65
ρ = -0.82, p = < 0.01
Diagnosed / total patients
Weighted avg EV-D68 / PMMoV

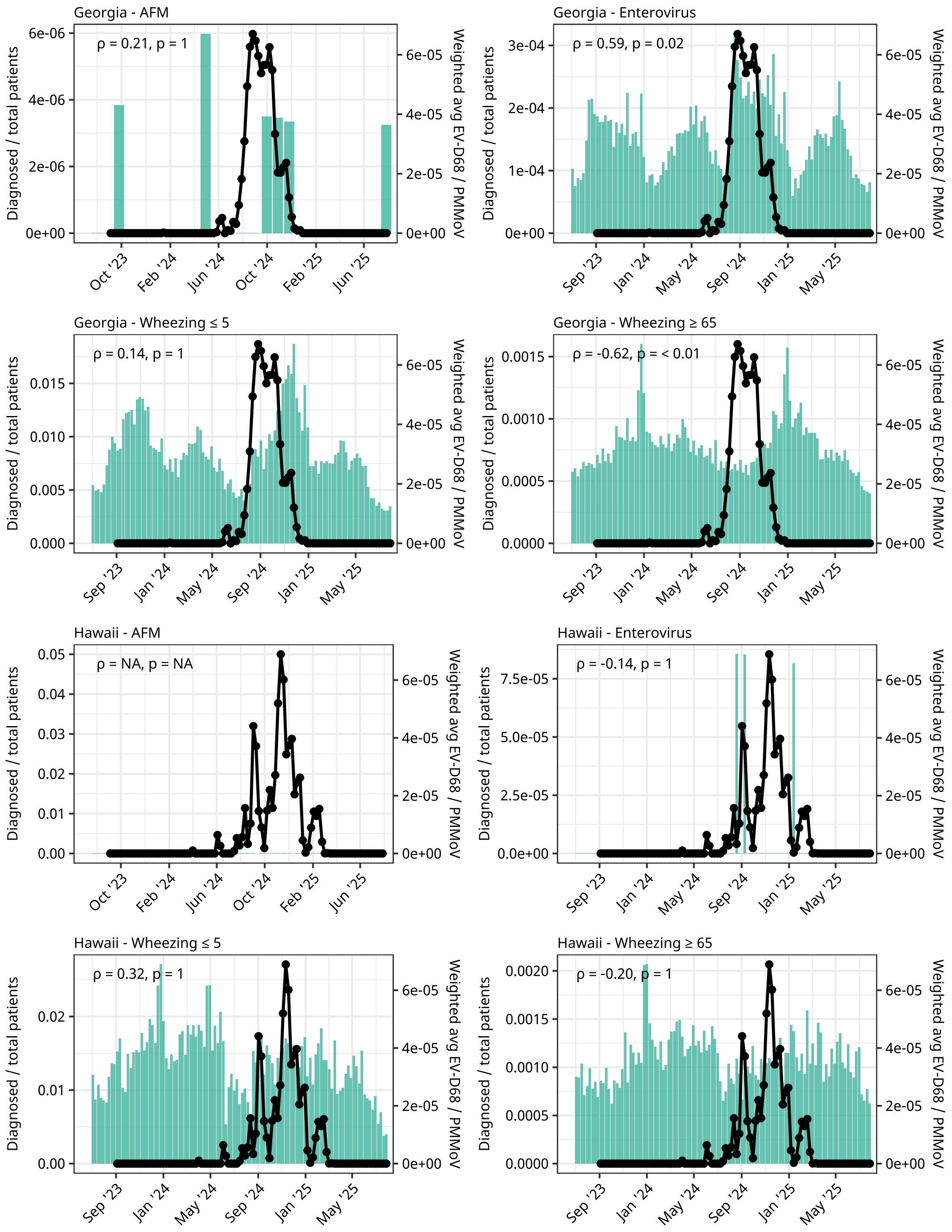
Georgia - AFM
ρ = 0.21, p = 1
Georgia - Enterovirus
ρ = 0.59, p = 0.02
Georgia - Wheezing ≤ 5
ρ = 0.14, p = 1
Georgia - Wheezing ≥ 65
ρ = -0.62, p = < 0.01
Hawaii - AFM
ρ = NA, p = NA
Hawaii - Enterovirus
ρ = -0.14, p = 1
Hawaii - Wheezing ≤ 5
ρ = 0.32, p = 1
Hawaii - Wheezing ≥ 65
ρ = -0.20, p = 1
Diagnosed / total patients
Weighted avg EV-D68 / PMMoV
Oct '23
Feb '24
Jun '24
Oct '24
Feb '25
Jun '25
Sep '23
Jan '24
May '24
Sep '24
Jan '25
May '25

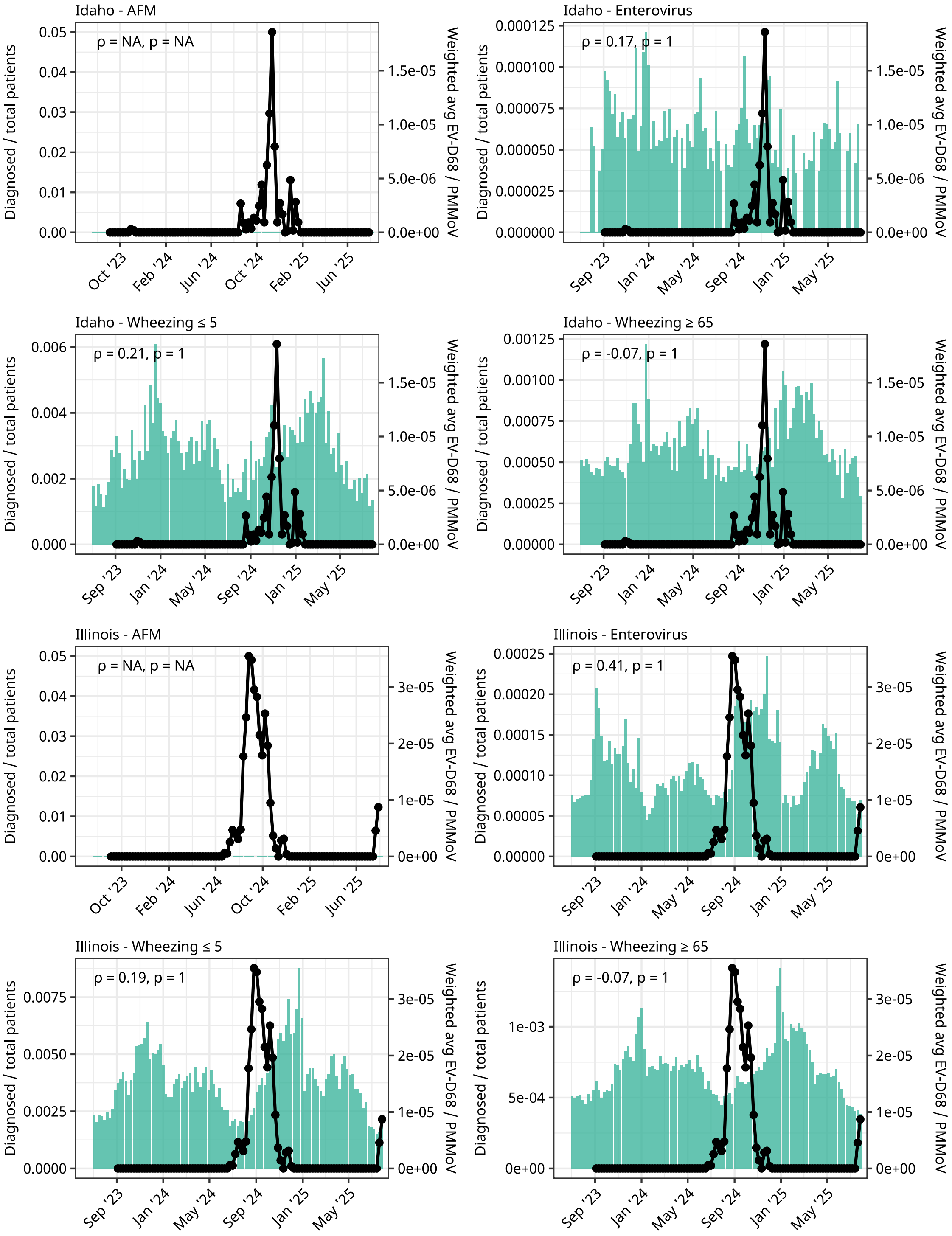
Idaho - AFM
ρ = NA, p = NA
Idaho - Enterovirus
ρ = 0.17, p = 1
Idaho - Wheezing ≤ 5
ρ = 0.21, p = 1
Idaho - Wheezing ≥ 65
ρ = -0.07, p = 1
Illinois - AFM
ρ = NA, p = NA
Illinois - Enterovirus
ρ = 0.41, p = 1
Illinois - Wheezing ≤ 5
ρ = 0.19, p = 1
Illinois - Wheezing ≥ 65
ρ = -0.07, p = 1
Diagnosed / total patients
Weighted avg EV-D68 / PMMoV
Sep '23
Oct '23
Jan '24
Feb '24
May '24
Jun '24
Sep '24
Oct '24
Jan '25
Feb '25
May '25
Jun '25

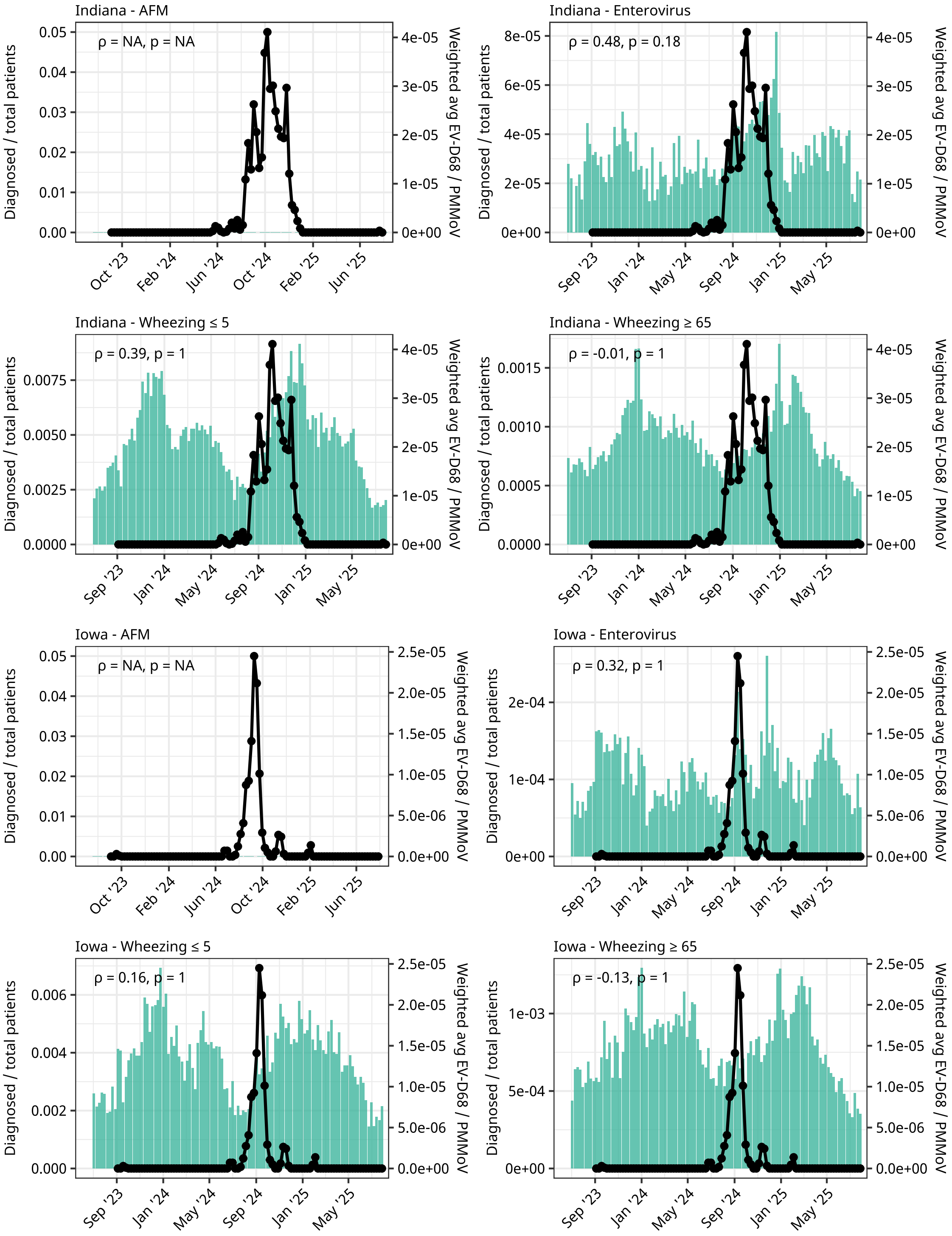
Indiana - AFM
ρ = NA, p = NA
Indiana - Enterovirus
ρ = 0.48, p = 0.18
Indiana - Wheezing ≤ 5
ρ = 0.39, p = 1
Indiana - Wheezing ≥ 65
ρ = -0.01, p = 1
Iowa - AFM
ρ = NA, p = NA
Iowa - Enterovirus
ρ = 0.32, p = 1
Iowa - Wheezing ≤ 5
ρ = 0.16, p = 1
Iowa - Wheezing ≥ 65
ρ = -0.13, p = 1
Diagnosed / total patients
Weighted avg EV-D68 / PMMoV

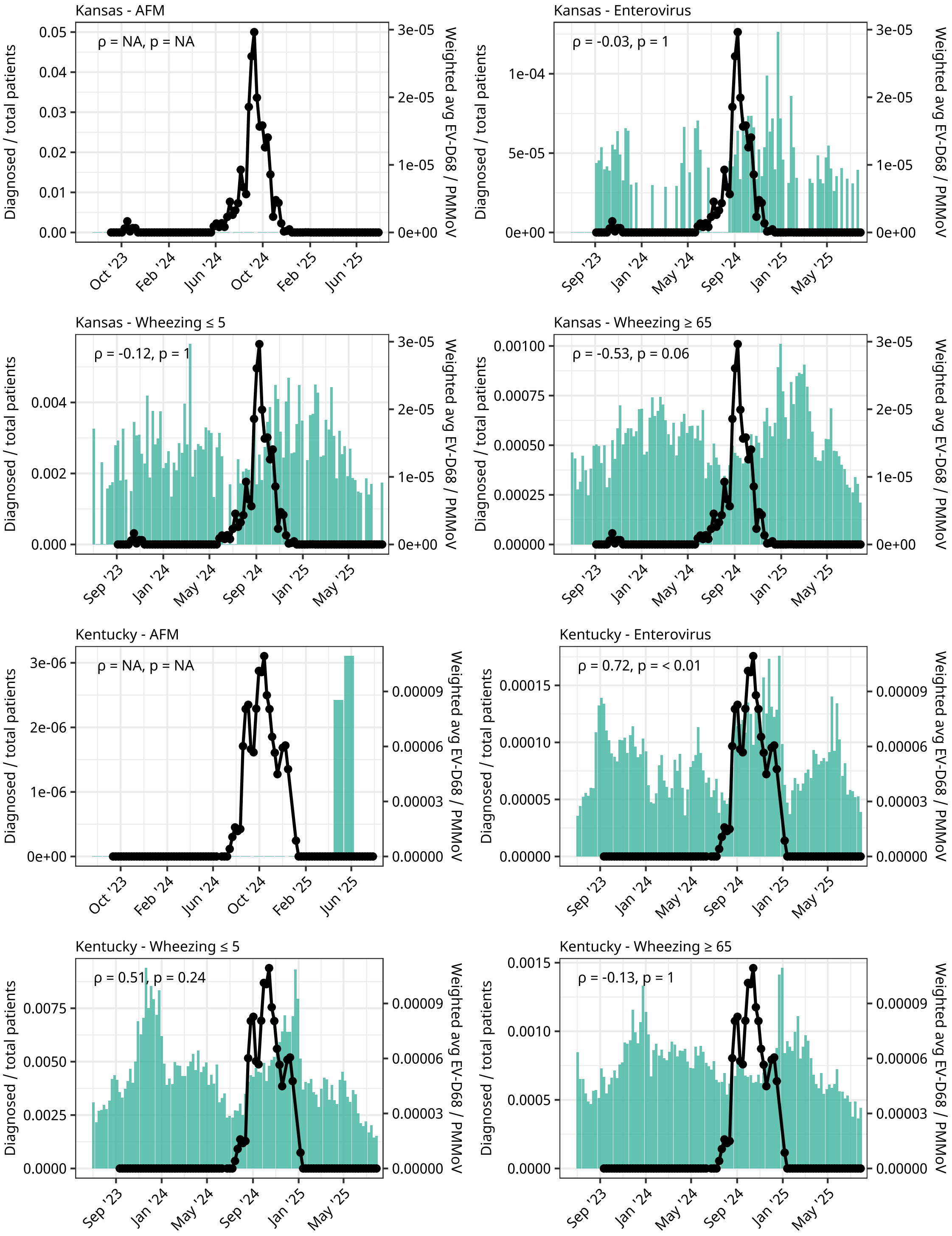
Kansas - AFM
ρ = NA, p = NA
Kansas - Enterovirus
ρ = -0.03, p = 1
Kansas - Wheezing ≤ 5
ρ = -0.12, p = 1
Kansas - Wheezing ≥ 65
ρ = -0.53, p = 0.06
Kentucky - AFM
ρ = NA, p = NA
Kentucky - Enterovirus
ρ = 0.72, p = < 0.01
Kentucky - Wheezing ≤ 5
ρ = 0.51, p = 0.24
Kentucky - Wheezing ≥ 65
ρ = -0.13, p = 1
Diagnosed / total patients
Weighted avg EV-D68 / PMMoV
Sep '23
Oct '23
Jan '24
Feb '24
May '24
Jun '24
Sep '24
Oct '24
Jan '25
Feb '25
May '25
Jun '25

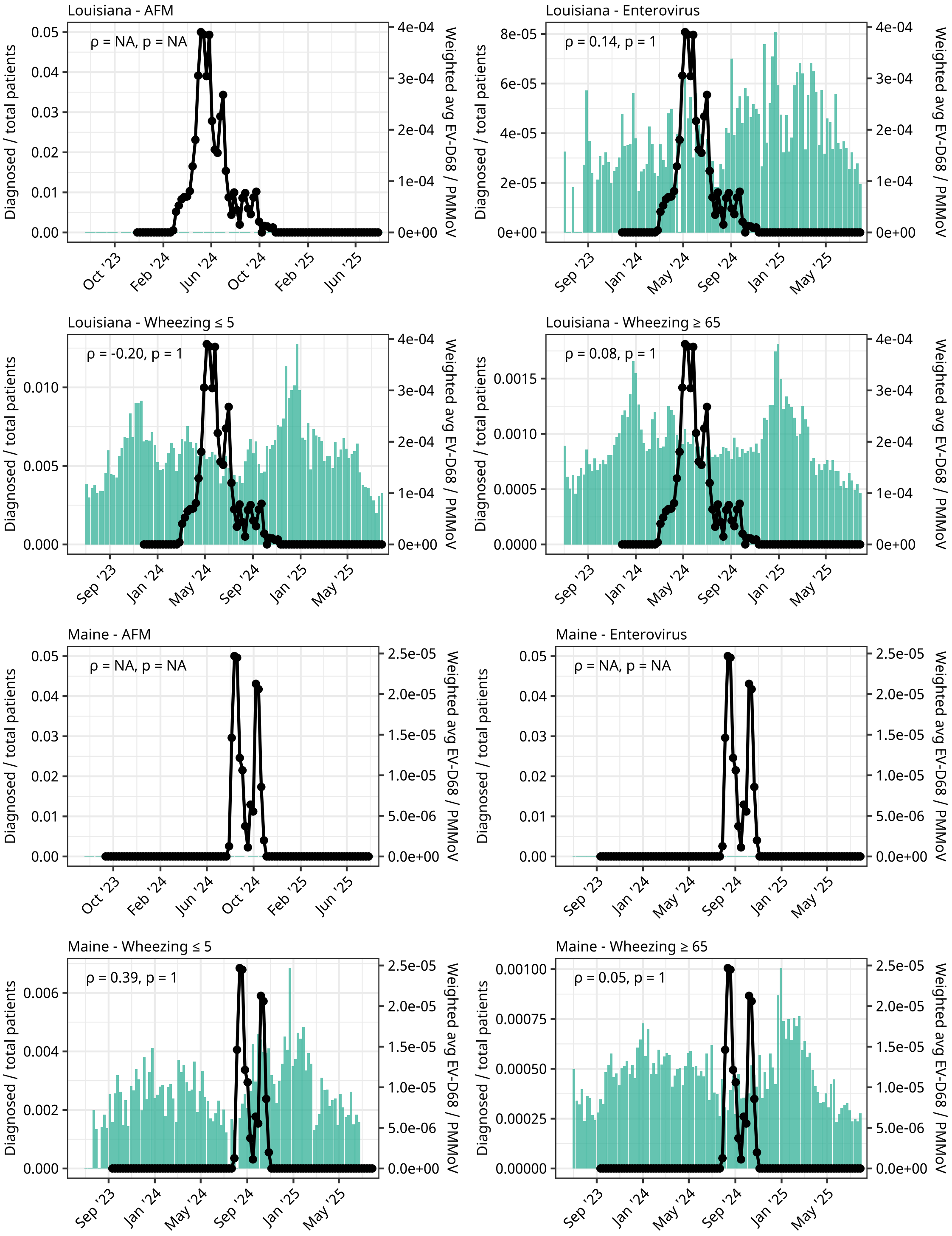
Louisiana - AFM
ρ = NA, p = NA
Louisiana - Enterovirus
ρ = 0.14, p = 1
Louisiana - Wheezing ≤ 5
ρ = -0.20, p = 1
Louisiana - Wheezing ≥ 65
ρ = 0.08, p = 1
Maine - AFM
ρ = NA, p = NA
Maine - Enterovirus
ρ = NA, p = NA
Maine - Wheezing ≤ 5
ρ = 0.39, p = 1
Maine - Wheezing ≥ 65
ρ = 0.05, p = 1
Diagnosed / total patients
Weighted avg EV-D68 / PMMoV

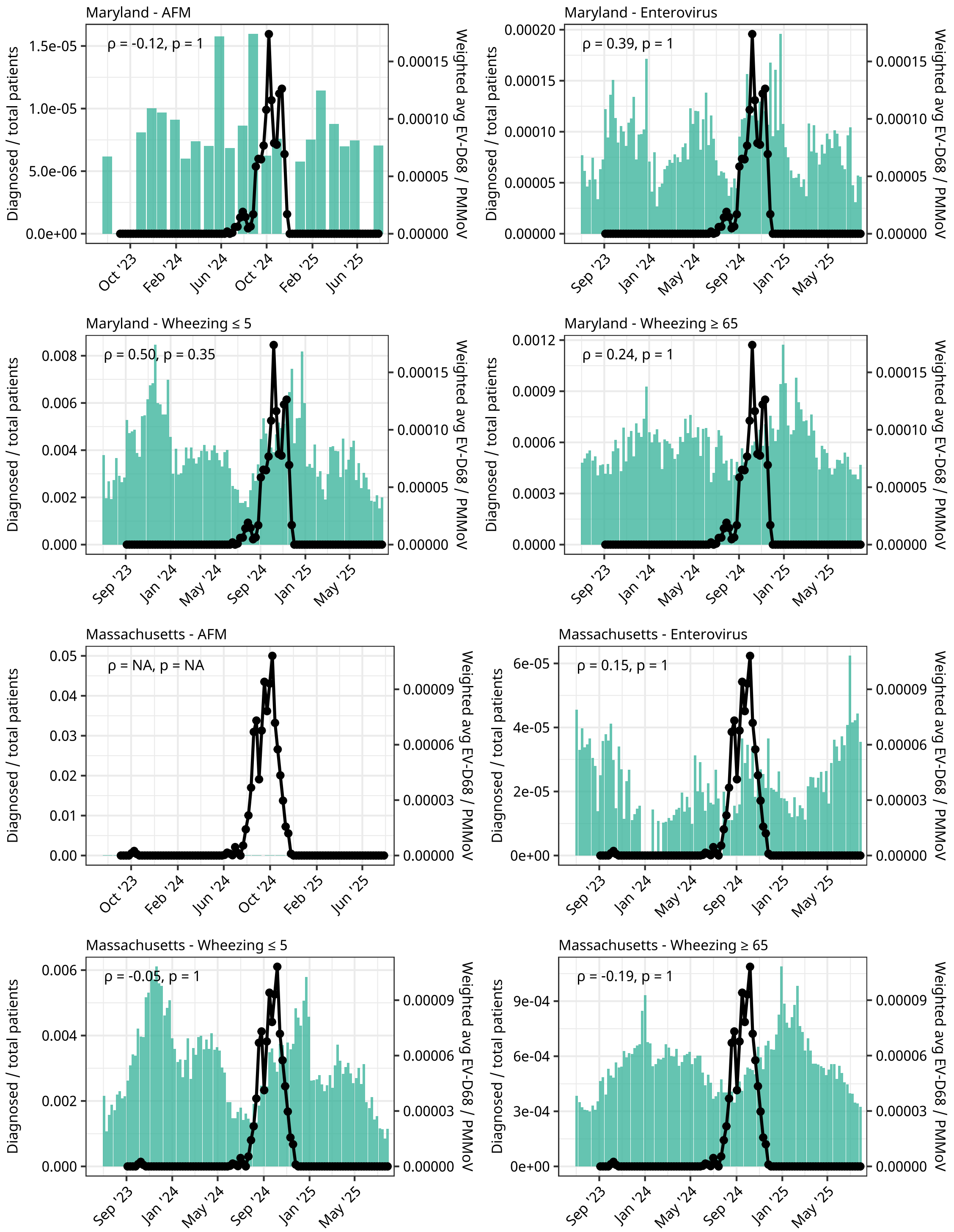
Maryland - AFM
ρ = -0.12, p = 1
Maryland - Enterovirus
ρ = 0.39, p = 1
Maryland - Wheezing ≤ 5
ρ = 0.50, p = 0.35
Maryland - Wheezing ≥ 65
ρ = 0.24, p = 1
Massachusetts - AFM
ρ = NA, p = NA
Massachusetts - Enterovirus
ρ = 0.15, p = 1
Massachusetts - Wheezing ≤ 5
ρ = -0.05, p = 1
Massachusetts - Wheezing ≥ 65
ρ = -0.19, p = 1
Diagnosed / total patients
Weighted avg EV-D68 / PMMoV

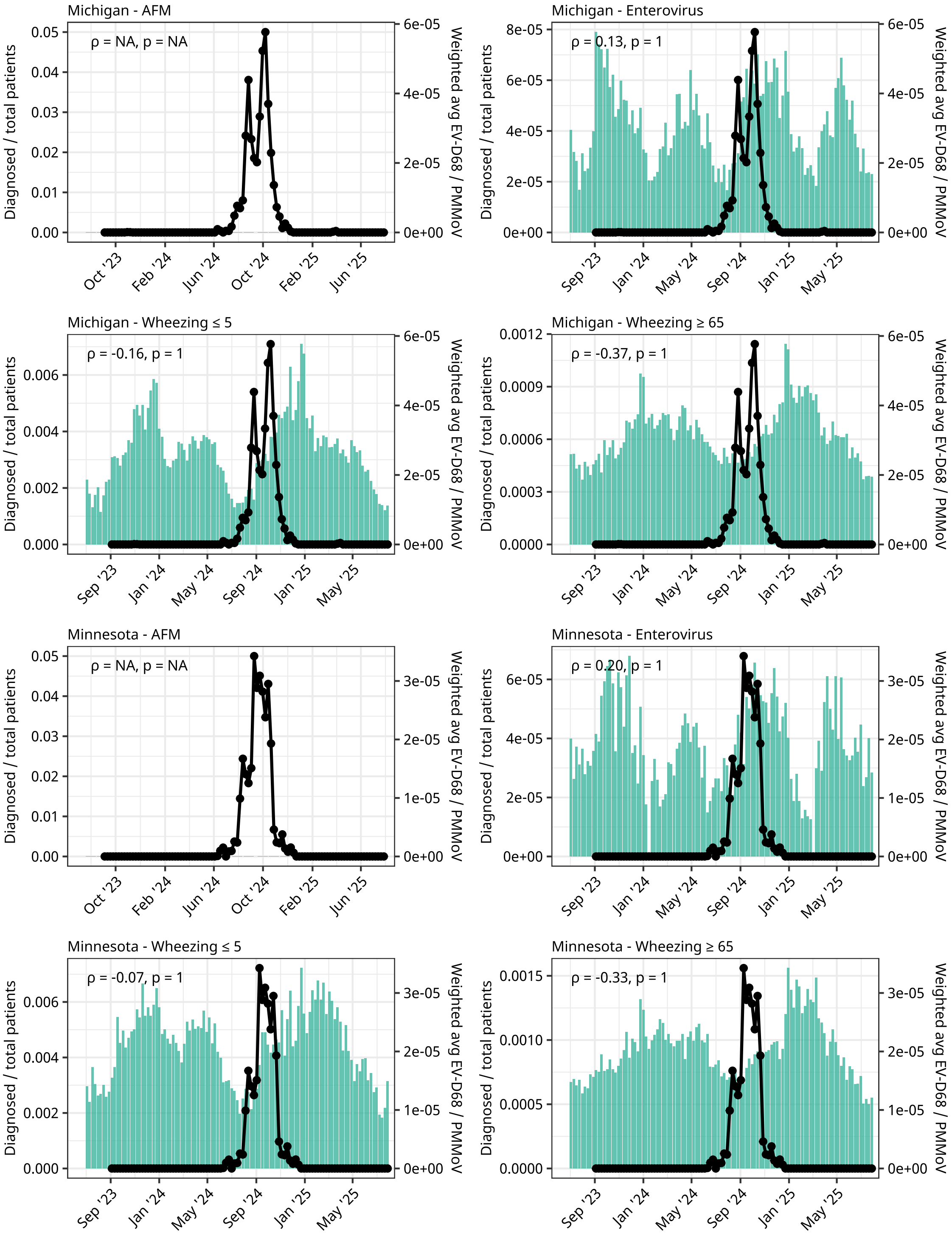
Michigan - AFM
ρ = NA, p = NA
Michigan - Enterovirus
ρ = 0.13, p = 1
Michigan - Wheezing ≤ 5
ρ = -0.16, p = 1
Michigan - Wheezing ≥ 65
ρ = -0.37, p = 1
Minnesota - AFM
ρ = NA, p = NA
Minnesota - Enterovirus
ρ = 0.20, p = 1
Minnesota - Wheezing ≤ 5
ρ = -0.07, p = 1
Minnesota - Wheezing ≥ 65
ρ = -0.33, p = 1
Diagnosed / total patients
Weighted avg EV-D68 / PMMoV
Oct '23
Feb '24
Jun '24
Oct '24
Feb '25
Jun '25
Sep '23
Jan '24
May '24
Sep '24
Jan '25
May '25

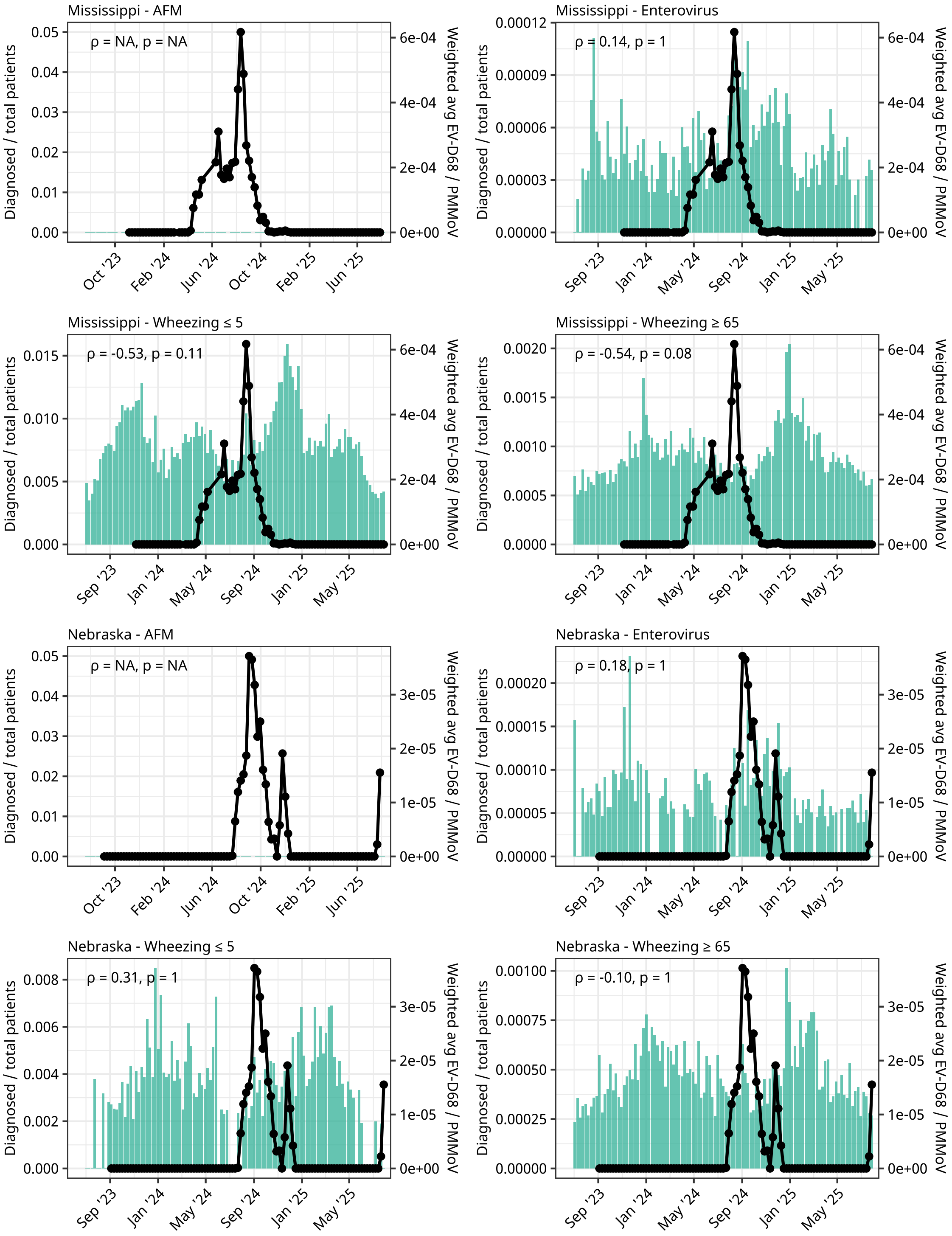
Mississippi - AFM
ρ = NA, p = NA
Mississippi - Enterovirus
ρ = 0.14, p = 1
Mississippi - Wheezing ≤ 5
ρ = -0.53, p = 0.11
Mississippi - Wheezing ≥ 65
ρ = -0.54, p = 0.08
Nebraska - AFM
ρ = NA, p = NA
Nebraska - Enterovirus
ρ = 0.18, p = 1
Nebraska - Wheezing ≤ 5
ρ = 0.31, p = 1
Nebraska - Wheezing ≥ 65
ρ = -0.10, p = 1
Diagnosed / total patients
Weighted avg EV-D68 / PMMoV
Sep '23
Oct '23
Jan '24
Feb '24
May '24
Jun '24
Sep '24
Oct '24
Jan '25
Feb '25
May '25
Jun '25

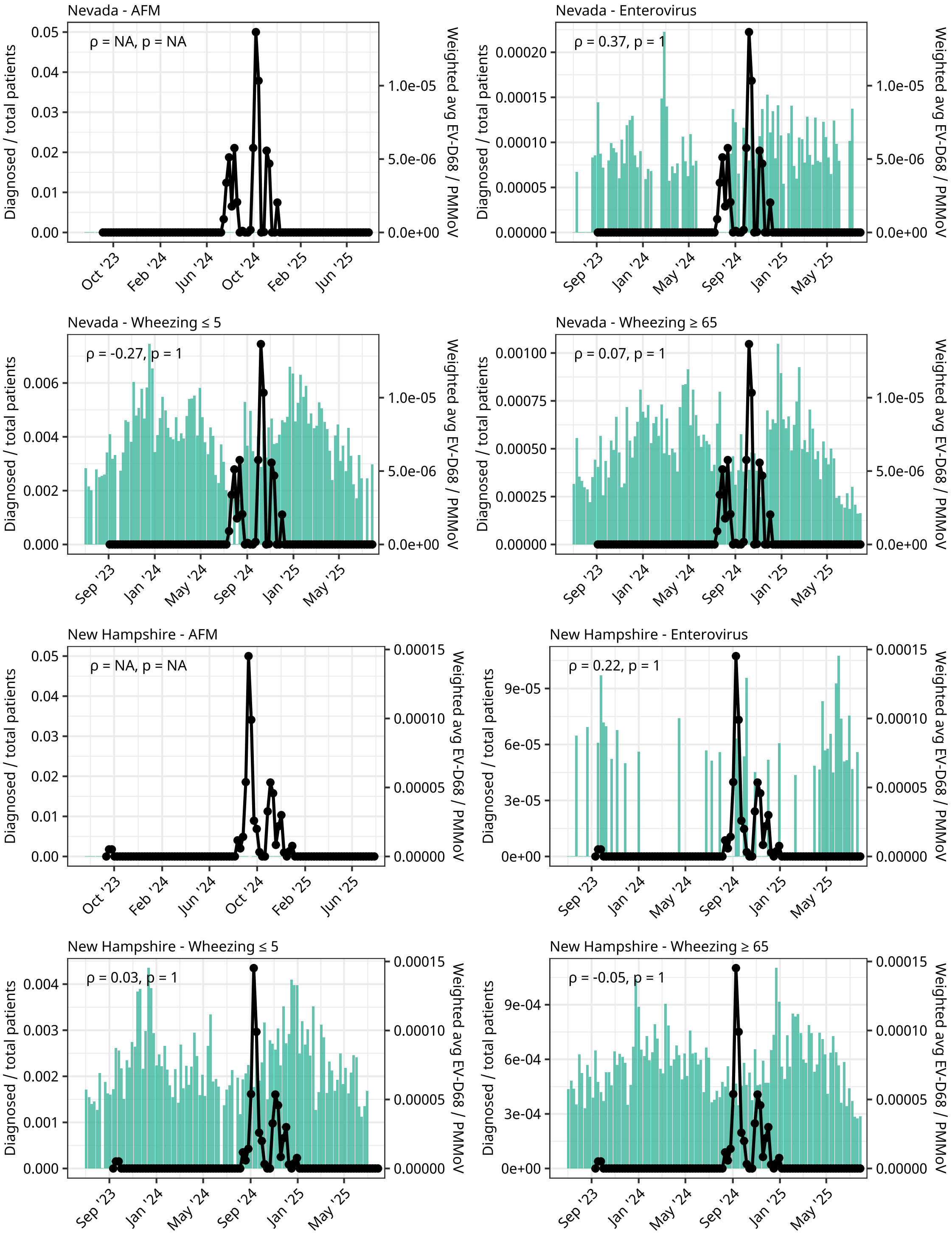

Nevada - AFM
ρ = NA, p = NA
Nevada - Enterovirus
ρ = 0.37, p = 1
Nevada - Wheezing ≤ 5
ρ = -0.27, p = 1
Nevada - Wheezing ≥ 65
ρ = 0.07, p = 1
New Hampshire - AFM
ρ = NA, p = NA
New Hampshire - Enterovirus
ρ = 0.22, p = 1
New Hampshire - Wheezing ≤ 5
ρ = 0.03, p = 1
New Hampshire - Wheezing ≥ 65
ρ = -0.05, p = 1
Diagnosed / total patients
Weighted avg EV-D68 / PMMoV
Oct '23
Feb '24
Jun '24
Oct '24
Feb '25
Jun '25
Sep '23
Jan '24
May '24
Sep '24
Jan '25
May '25

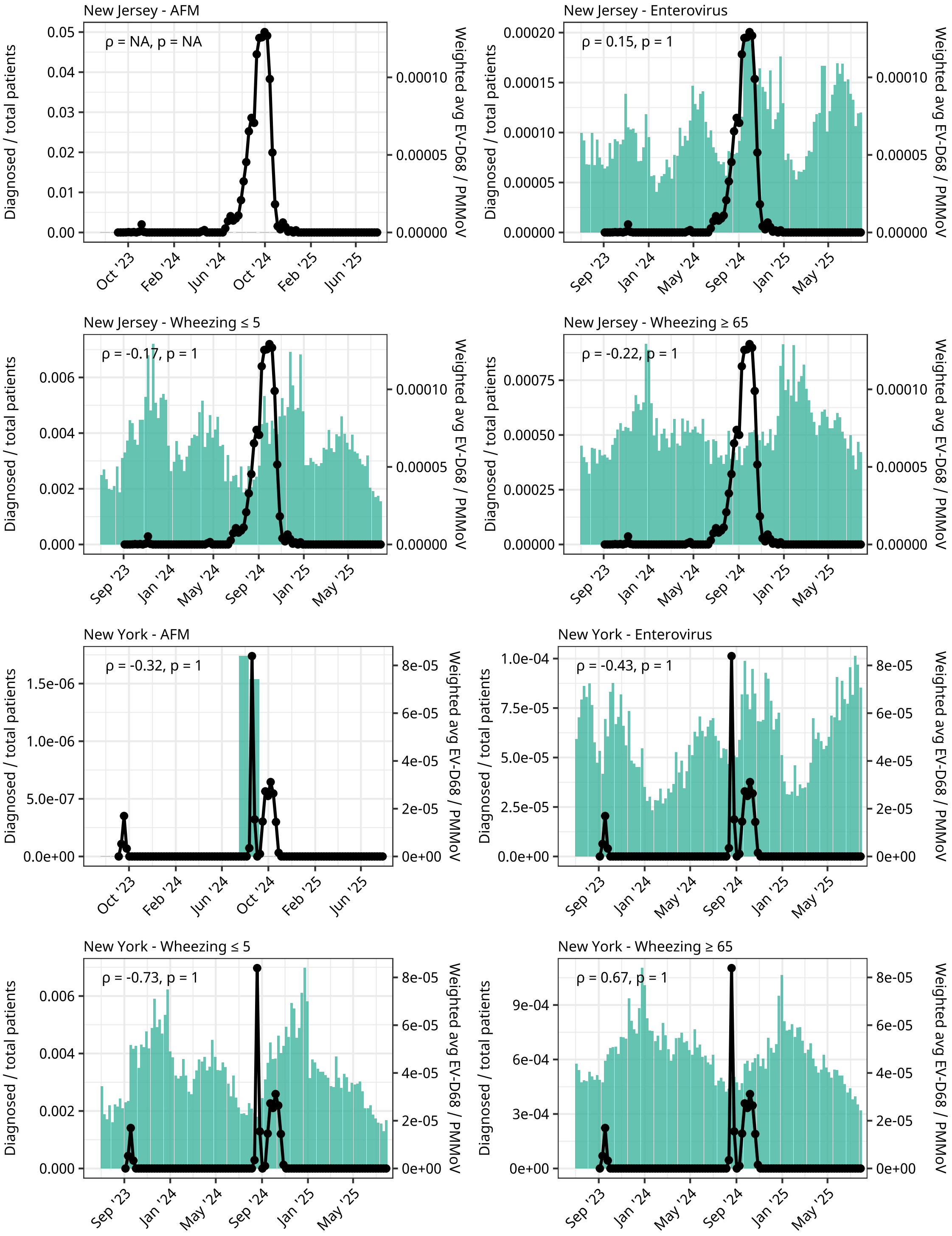
New Jersey - AFM
ρ = NA, p = NA
New Jersey - Enterovirus
ρ = 0.15, p = 1
New Jersey - Wheezing ≤ 5
ρ = -0.17, p = 1
New Jersey - Wheezing ≥ 65
ρ = -0.22, p = 1
New York - AFM
ρ = -0.32, p = 1
New York - Enterovirus
ρ = -0.43, p = 1
New York - Wheezing ≤ 5
ρ = -0.73, p = 1
New York - Wheezing ≥ 65
ρ = 0.67, p = 1
Diagnosed / total patients
Weighted avg EV-D68 / PMMoV
Oct '23
Feb '24
Jun '24
Oct '24
Feb '25
Jun '25
Sep '23
Jan '24
May '24
Sep '24
Jan '25
May '25

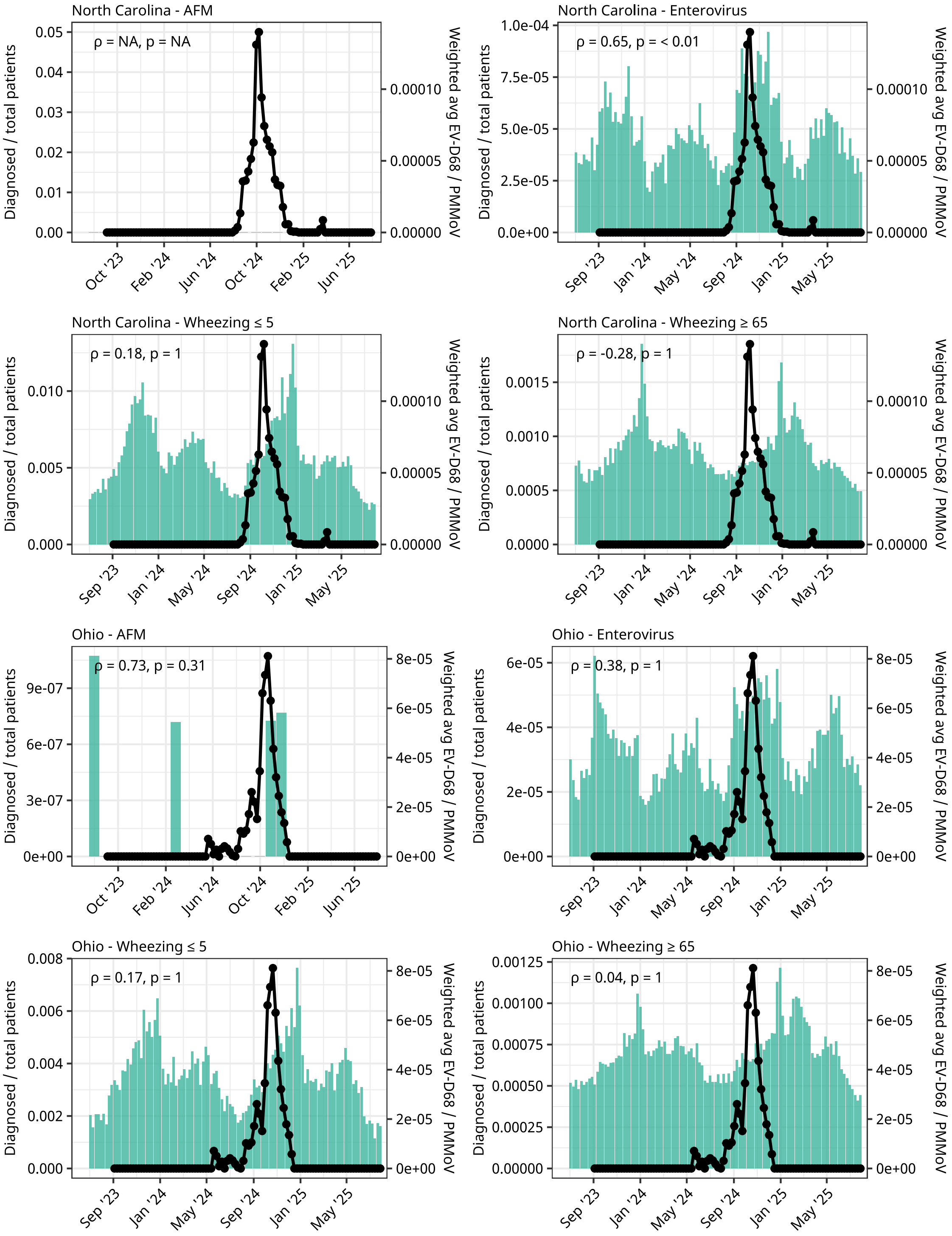
North Carolina - AFM
ρ = NA, p = NA
North Carolina - Enterovirus
ρ = 0.65, p = < 0.01
North Carolina - Wheezing ≤ 5
ρ = 0.18, p = 1
North Carolina - Wheezing ≥ 65
ρ = -0.28, p = 1
Ohio - AFM
ρ = 0.73, p = 0.31
Ohio - Enterovirus
ρ = 0.38, p = 1
Ohio - Wheezing ≤ 5
ρ = 0.17, p = 1
Ohio - Wheezing ≥ 65
ρ = 0.04, p = 1
Diagnosed / total patients
Weighted avg EV-D68 / PMMoV
Oct '23
Feb '24
Jun '24
Oct '24
Feb '25
Jun '25
Sep '23
Jan '24
May '24
Sep '24
Jan '25
May '25

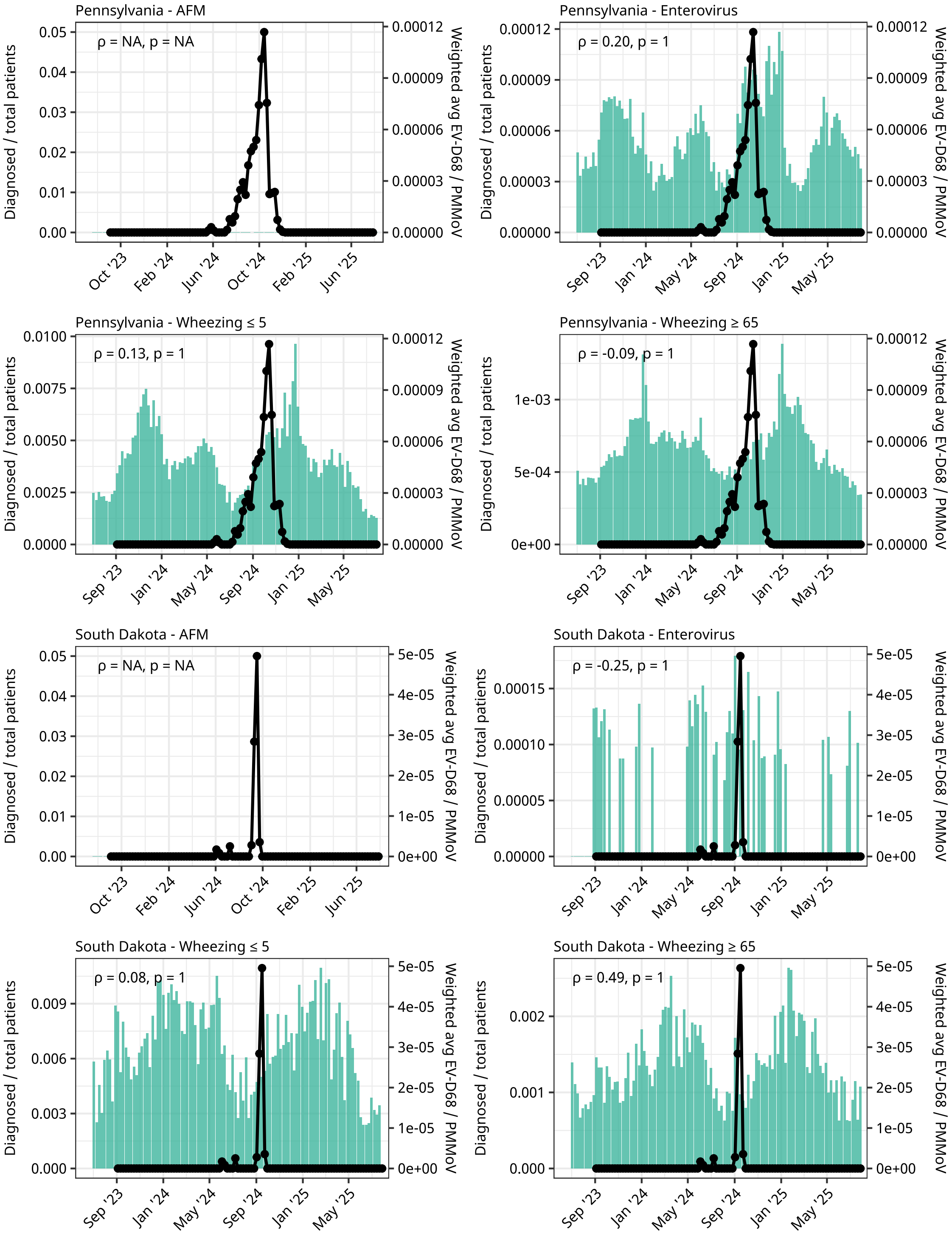
Pennsylvania - AFM
ρ = NA, p = NA
Pennsylvania - Enterovirus
ρ = 0.20, p = 1
Pennsylvania - Wheezing ≤ 5
ρ = 0.13, p = 1
Pennsylvania - Wheezing ≥ 65
ρ = -0.09, p = 1
South Dakota - AFM
ρ = NA, p = NA
South Dakota - Enterovirus
ρ = -0.25, p = 1
South Dakota - Wheezing ≤ 5
ρ = 0.08, p = 1
South Dakota - Wheezing ≥ 65
ρ = 0.49, p = 1
Diagnosed / total patients
Weighted avg EV-D68 / PMMoV
Oct '23
Feb '24
Jun '24
Oct '24
Feb '25
Jun '25
Sep '23
Jan '24
May '24
Sep '24
Jan '25
May '25

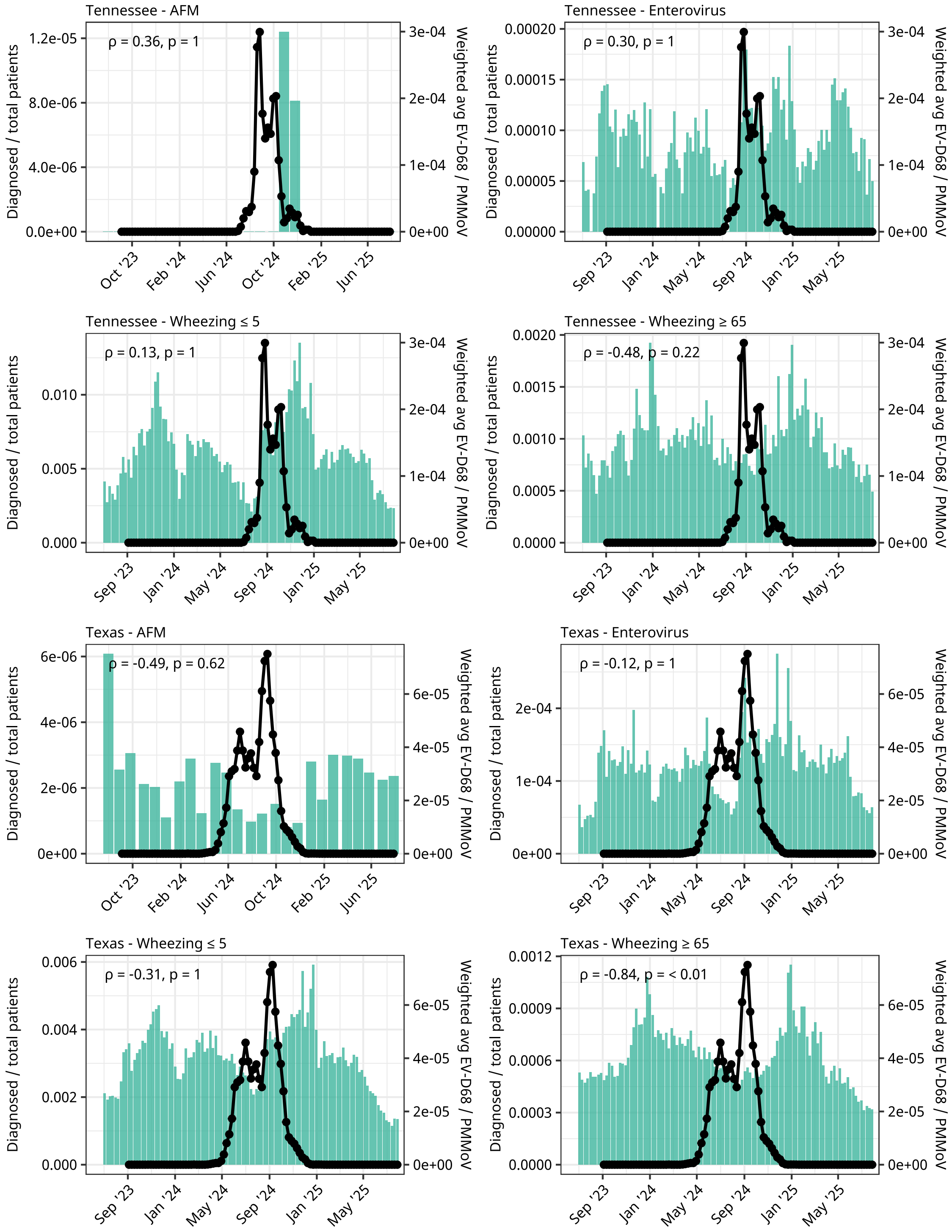

Tennessee - AFM
ρ = 0.36, p = 1
Tennessee - Enterovirus
ρ = 0.30, p = 1
Tennessee - Wheezing ≤ 5
ρ = 0.13, p = 1
Tennessee - Wheezing ≥ 65
ρ = -0.48, p = 0.22
Texas - AFM
ρ = -0.49, p = 0.62
Texas - Enterovirus
ρ = -0.12, p = 1
Texas - Wheezing ≤ 5
ρ = -0.31, p = 1
Texas - Wheezing ≥ 65
ρ = -0.84, p = < 0.01
Diagnosed / total patients
Weighted avg EV-D68 / PMMoV

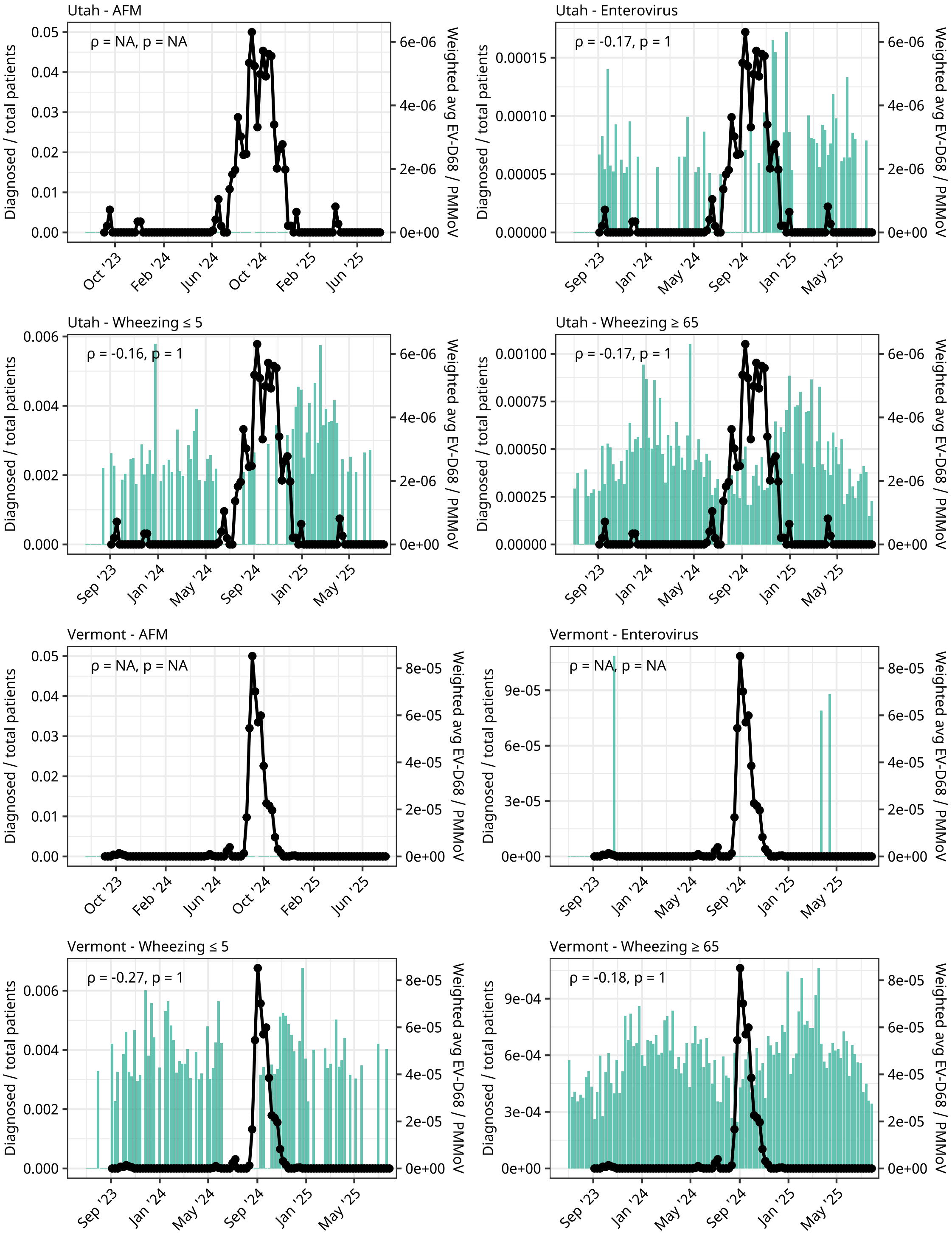
Utah - AFM
ρ = NA, p = NA
Utah - Enterovirus
ρ = -0.17, p = 1
Utah - Wheezing ≤ 5
ρ = -0.16, p = 1
Utah - Wheezing ≥ 65
ρ = -0.17, p = 1
Vermont - AFM
ρ = NA, p = NA
Vermont - Enterovirus
ρ = NA, p = NA
Vermont - Wheezing ≤ 5
ρ = -0.27, p = 1
Vermont - Wheezing ≥ 65
ρ = -0.18, p = 1
Diagnosed / total patients
Weighted avg EV-D68 / PMMoV

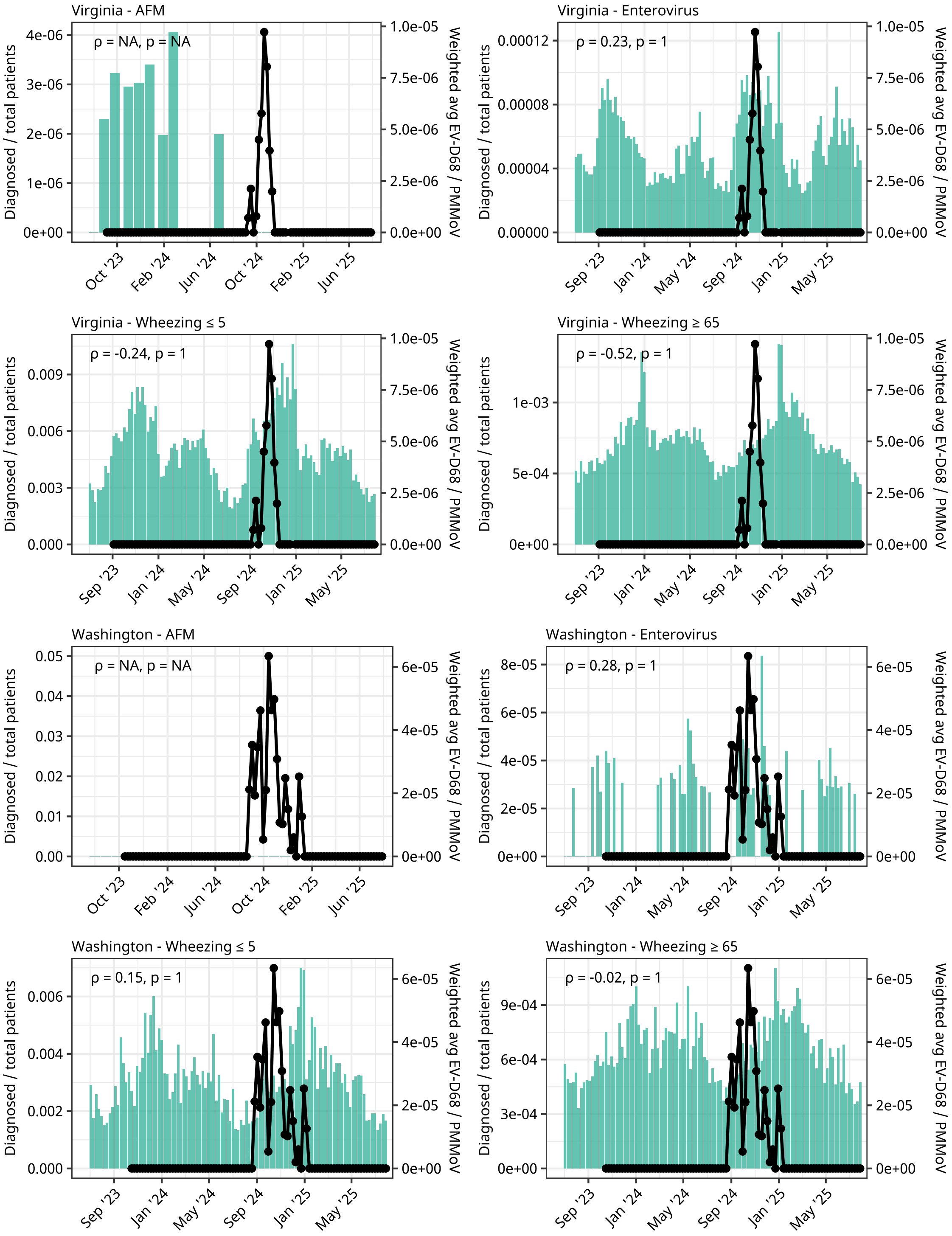
Virginia - AFM
ρ = NA, p = NA
Virginia - Enterovirus
ρ = 0.23, p = 1
Virginia - Wheezing ≤ 5
ρ = -0.24, p = 1
Virginia - Wheezing ≥ 65
ρ = -0.52, p = 1
Washington - AFM
ρ = NA, p = NA
Washington - Enterovirus
ρ = 0.28, p = 1
Washington - Wheezing ≤ 5
ρ = 0.15, p = 1
Washington - Wheezing ≥ 65
ρ = -0.02, p = 1
Diagnosed / total patients
Weighted avg EV-D68 / PMMoV

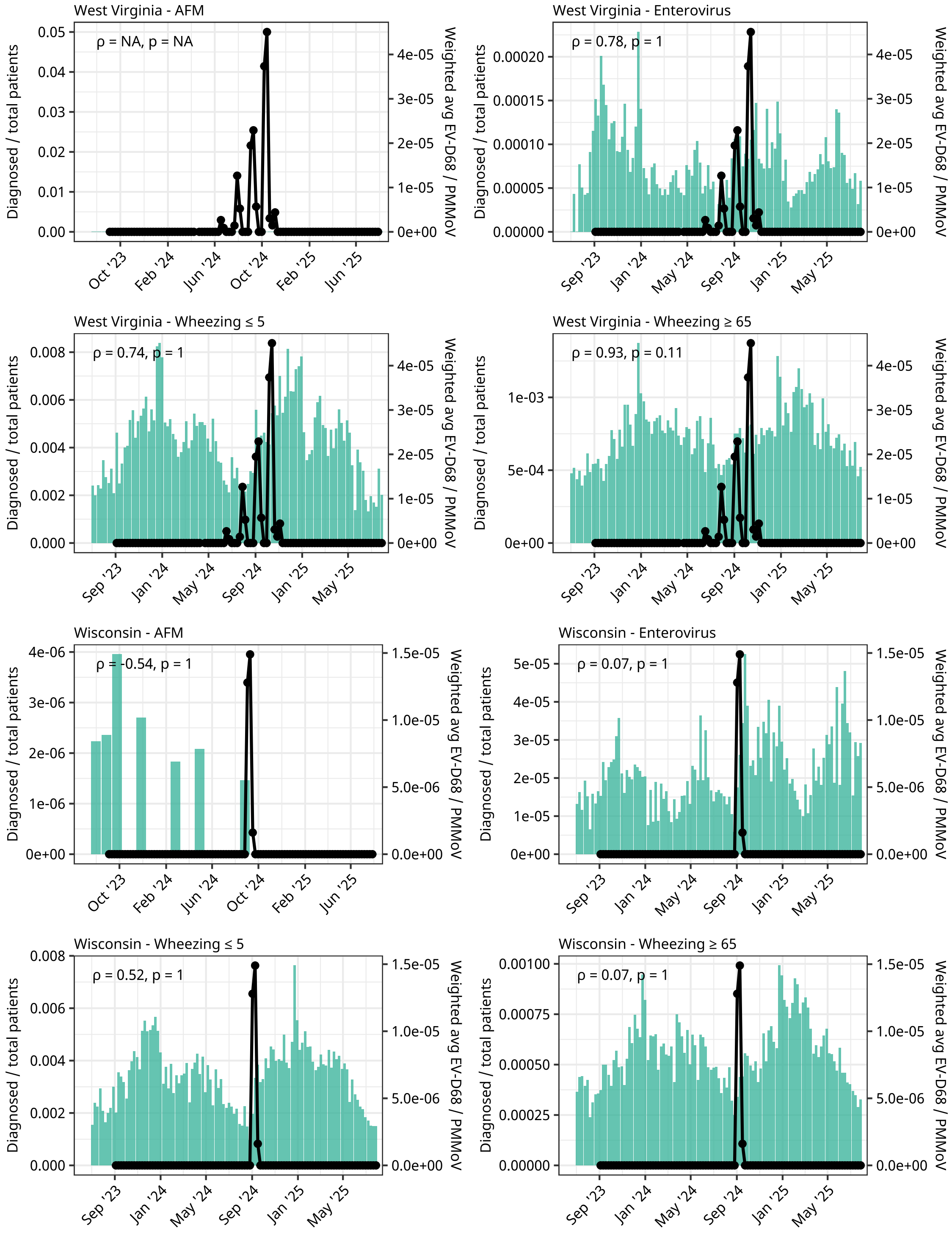


**Fig. S8.** State-level trends in wastewater EV-D68 RNA concentrations and clinical diagnoses from Epic Cosmos. For each state, bars show the weekly proportion of diagnosed encounters for the indicated condition, and the black line with points shows the population-weighted mean wastewater PMMoV normalized EV-D68 RNA concentrations. Plot titles indicate the state and the diagnosis. The label in the top left reports the Spearman correlation (ρ) and the Bonferroni-adjusted p-value between wastewater and the diagnosis within that state. Correlations were calculated over the season duration of EV-D68 defined from wastewater for that state, extended by ±2 weeks for wheezing ≤ 5 years, wheezing ≥ 65 years, and enterovirus-specific encounters, and by ±2 months for acute flaccid myelitis (AFM). AFM uses monthly aggregation; all other diagnoses use weekly data. Panels with no correlation result are labeled “ρ = NA, p = NA.”

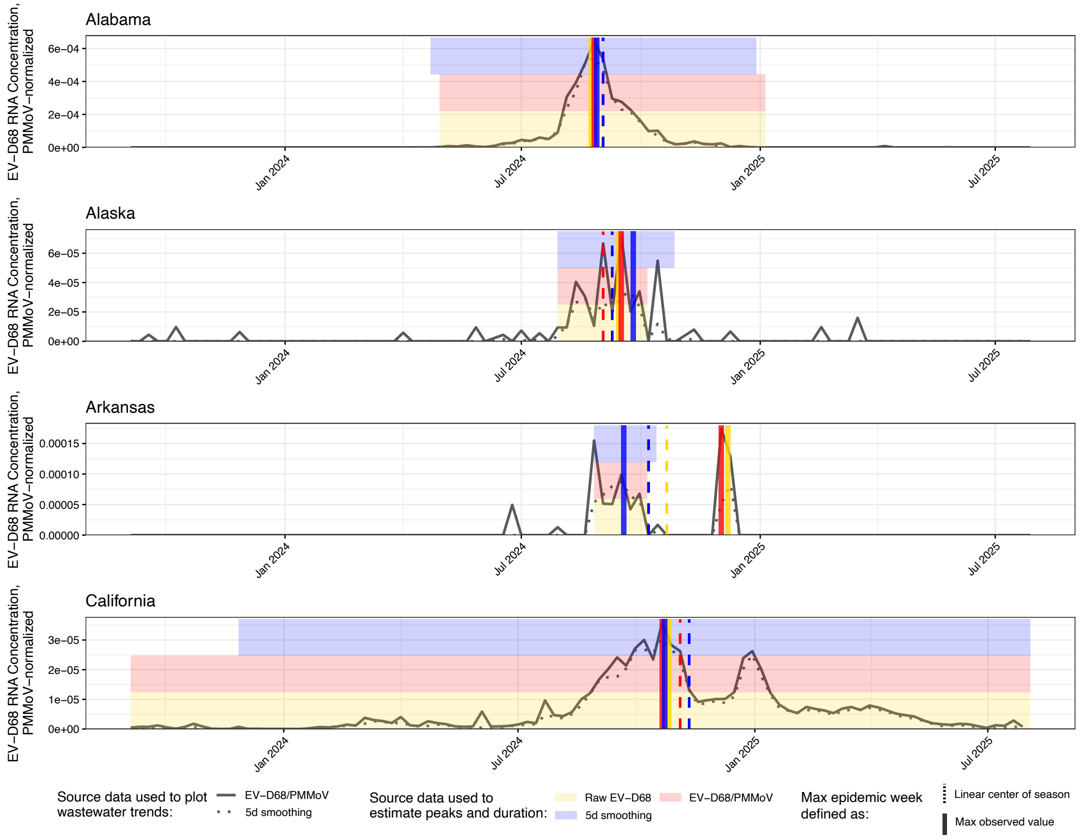
Alabama
Alaska
Arkansas
California
EV−D68 RNA Concentration, PMMoV−normalized
Jan 2024
Jul 2024
Jan 2025
Jul 2025
Source data used to plot wastewater trends:
EV−D68/PMMoV
5d smoothing
Source data used to estimate peaks and duration:
Raw EV−D68
EV−D68/PMMoV
5d smoothing
Max epidemic week defined as:
Linear center of season
Max observed value

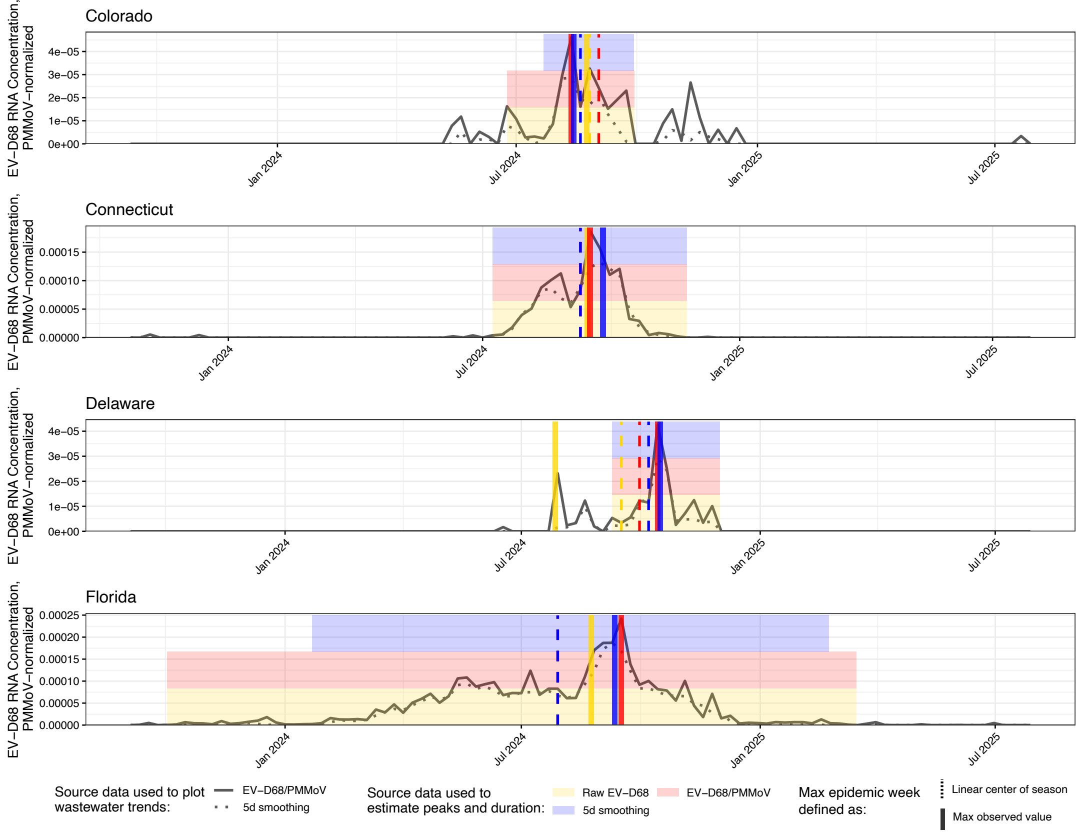
Colorado
Connecticut
Delaware
Florida
EV−D68 RNA Concentration, PMMoV−normalized
Jan 2024
Jul 2024
Jan 2025
Jul 2025
Source data used to plot wastewater trends:
EV−D68/PMMoV
5d smoothing
Source data used to estimate peaks and duration:
Raw EV−D68
EV−D68/PMMoV
5d smoothing
Max epidemic week defined as:
Linear center of season
Max observed value

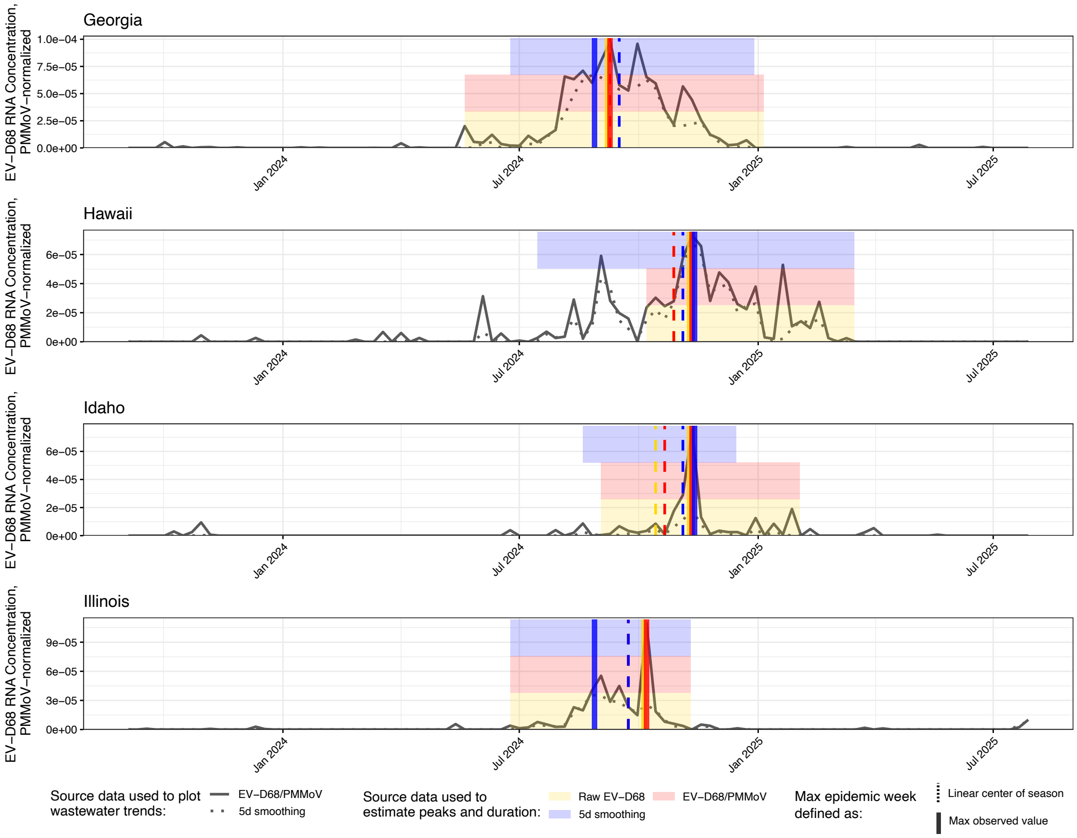
Georgia
Hawaii
Idaho
Illinois
EV−D68 RNA Concentration, PMMoV−normalized
1.0e−04
7.5e−05
5.0e−05
2.5e−05
0.0e+00
6e−05
4e−05
2e−05
0e+00
9e−05
3e−05
Jan 2024
Jul 2024
Jan 2025
Jul 2025
Source data used to plot wastewater trends:
EV−D68/PMMoV
5d smoothing
Source data used to estimate peaks and duration:
Raw EV−D68
EV−D68/PMMoV
5d smoothing
Max epidemic week defined as:
Linear center of season
Max observed value

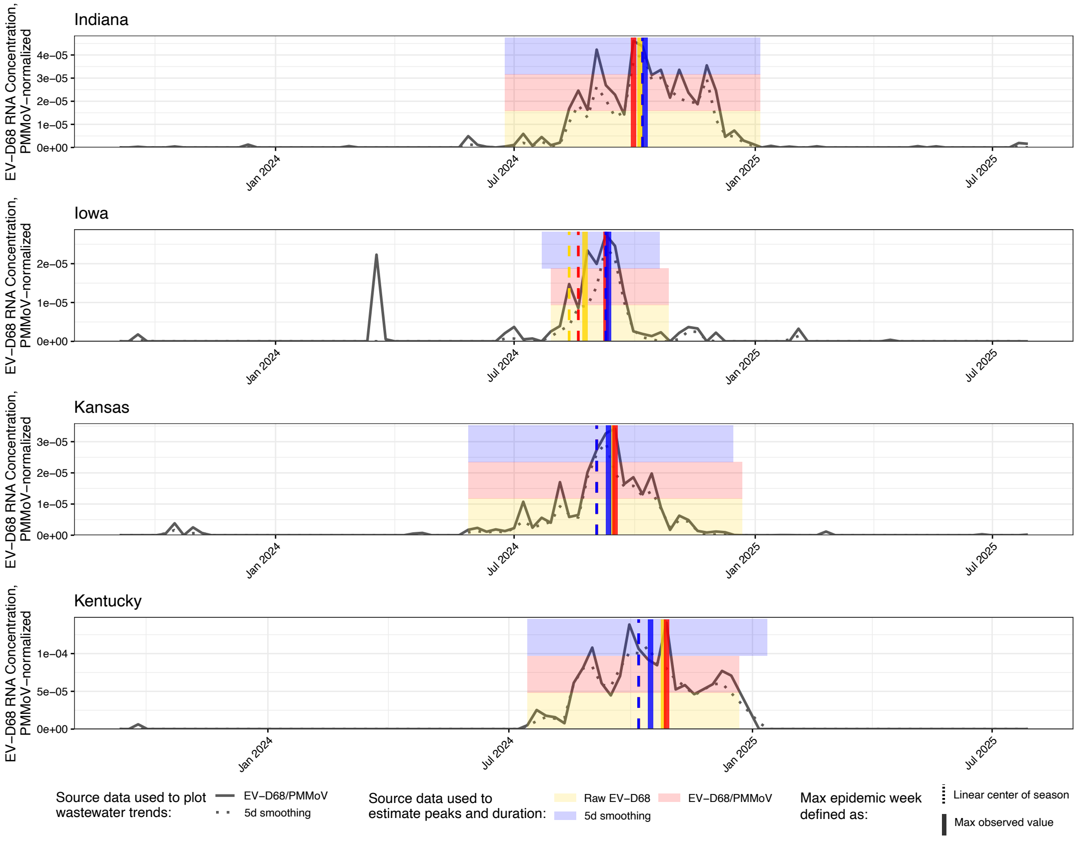
Indiana
Iowa
Kansas
Kentucky
EV−D68 RNA Concentration, PMMoV−normalized
4e−05
3e−05
2e−05
1e−05
0e+00
1e−04
5e−05
Jan 2024
Jul 2024
Jan 2025
Jul 2025
Source data used to plot wastewater trends:
EV−D68/PMMoV
5d smoothing
Source data used to estimate peaks and duration:
Raw EV−D68
EV−D68/PMMoV
5d smoothing
Max epidemic week defined as:
Linear center of season
Max observed value

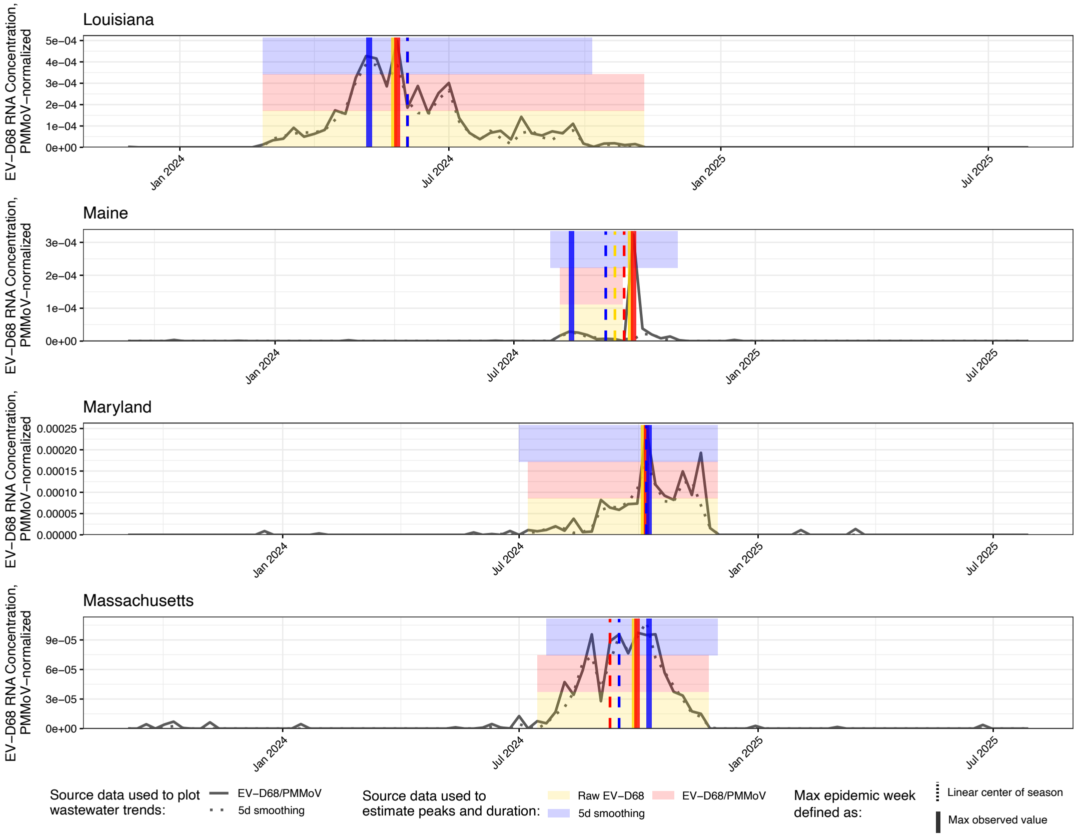
Louisiana
Maine
Maryland
Massachusetts
EV−D68 RNA Concentration, PMMoV−normalized
5e−04
4e−04
3e−04
2e−04
1e−04
0e+00
3e−04
2e−04
1e−04
0.00025
0.00020
0.00015
0.00010
0.00005
0.00000
9e−05
6e−05
3e−05
Jan 2024
Jul 2024
Jan 2025
Jul 2025
Source data used to plot wastewater trends:
EV−D68/PMMoV
5d smoothing
Source data used to estimate peaks and duration:
Raw EV−D68
EV−D68/PMMoV
5d smoothing
Max epidemic week defined as:
Linear center of season
Max observed value

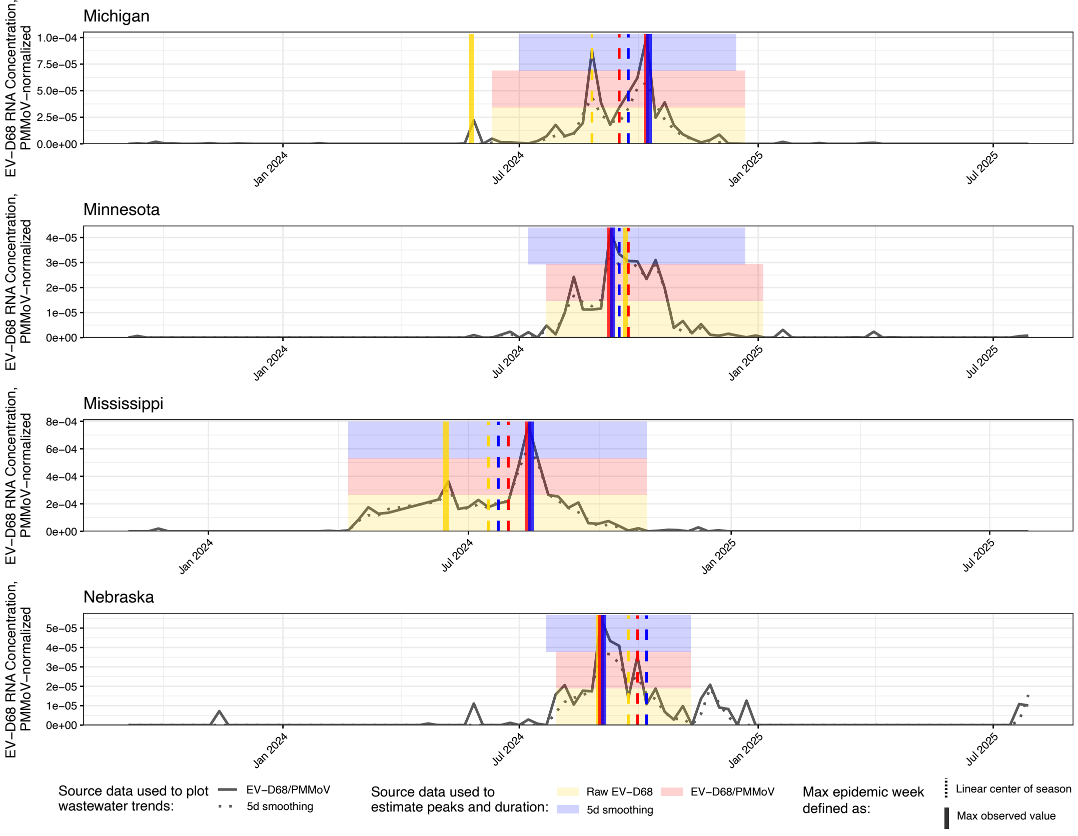
Michigan
Minnesota
Mississippi
Nebraska
EV−D68 RNA Concentration, PMMoV−normalized
1.0e−04
7.5e−05
5.0e−05
2.5e−05
0.0e+00
4e−05
3e−05
2e−05
1e−05
0e+00
8e−04
6e−04
4e−04
2e−04
5e−05
Jan 2024
Jul 2024
Jan 2025
Jul 2025
Source data used to plot wastewater trends:
EV−D68/PMMoV
5d smoothing
Source data used to estimate peaks and duration:
Raw EV−D68
EV−D68/PMMoV
5d smoothing
Max epidemic week defined as:
Linear center of season
Max observed value

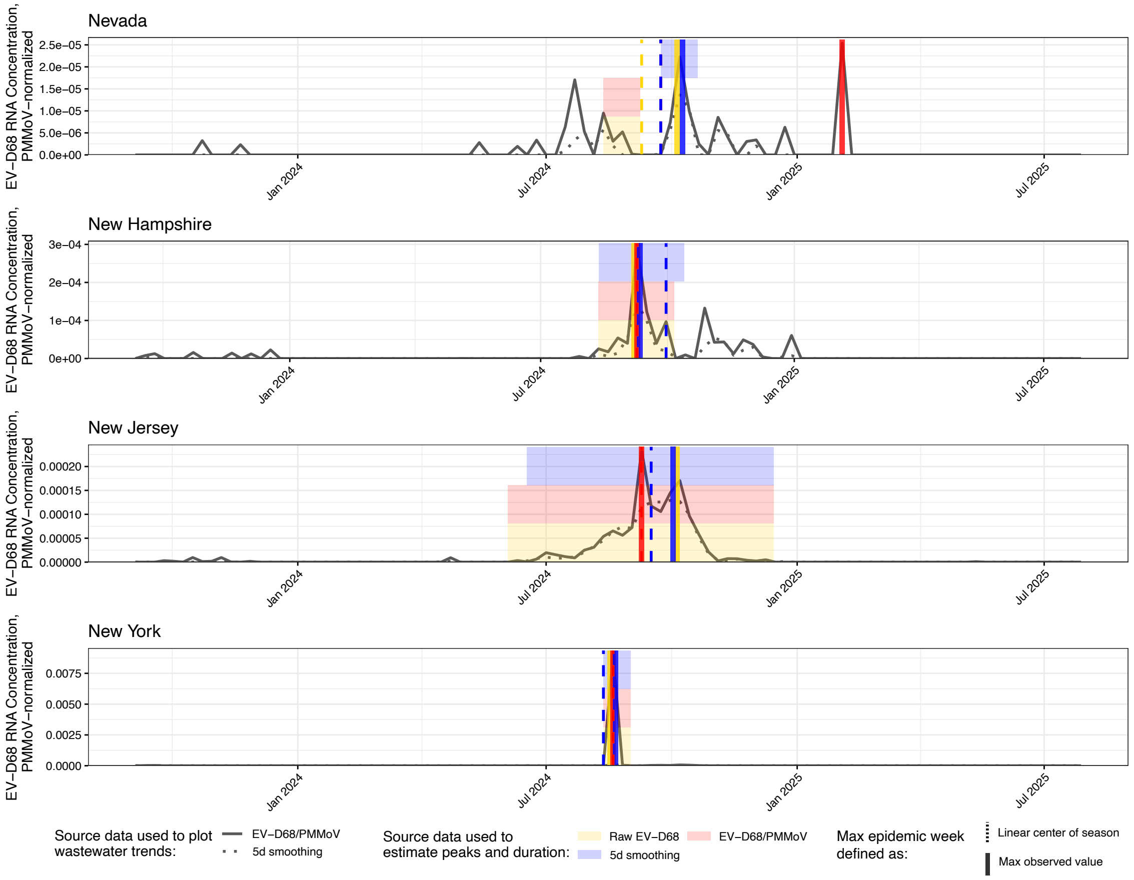
Nevada
New Hampshire
New Jersey
New York
EV−D68 RNA Concentration, PMMoV−normalized
Jan 2024
Jul 2024
Jan 2025
Jul 2025
Source data used to plot wastewater trends:
EV−D68/PMMoV
5d smoothing
Source data used to estimate peaks and duration:
Raw EV−D68
EV−D68/PMMoV
5d smoothing
Max epidemic week defined as:
Linear center of season
Max observed value

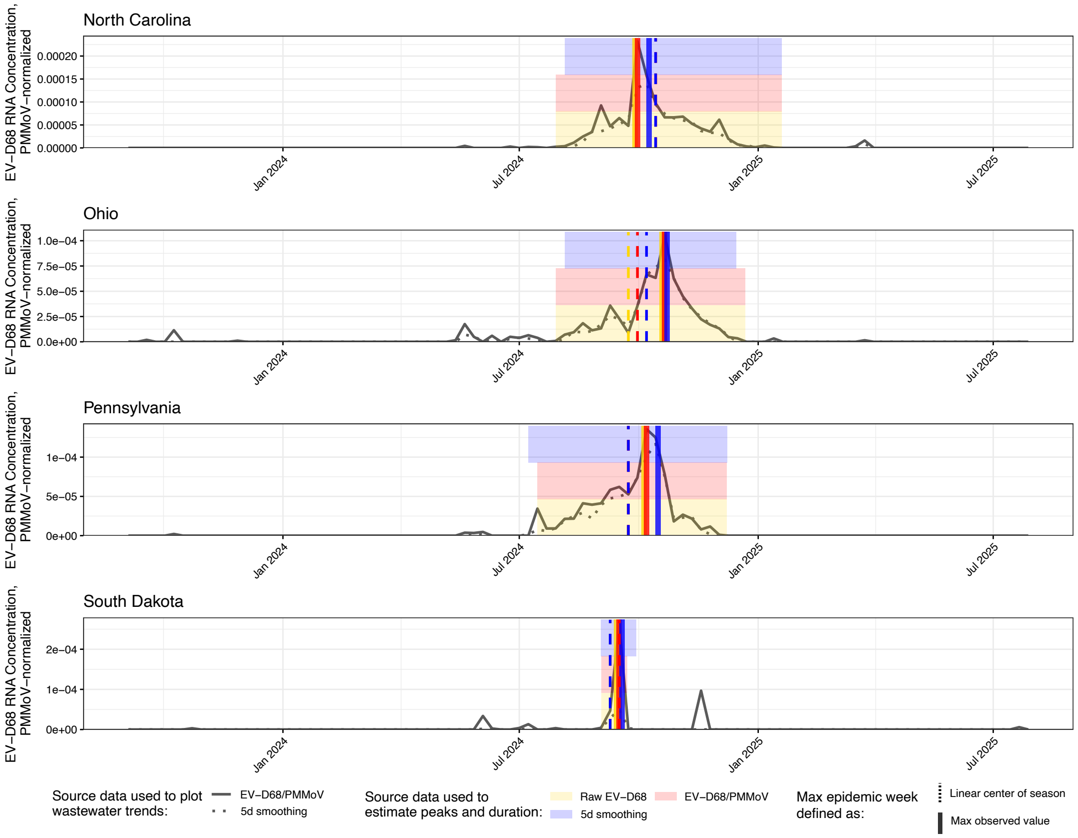
North Carolina
Ohio
Pennsylvania
South Dakota
EV−D68 RNA Concentration, PMMoV−normalized
Jan 2024
Jul 2024
Jan 2025
Jul 2025
Source data used to plot wastewater trends:
EV−D68/PMMoV
5d smoothing
Source data used to estimate peaks and duration:
Raw EV−D68
EV−D68/PMMoV
5d smoothing
Max epidemic week defined as:
Linear center of season
Max observed value

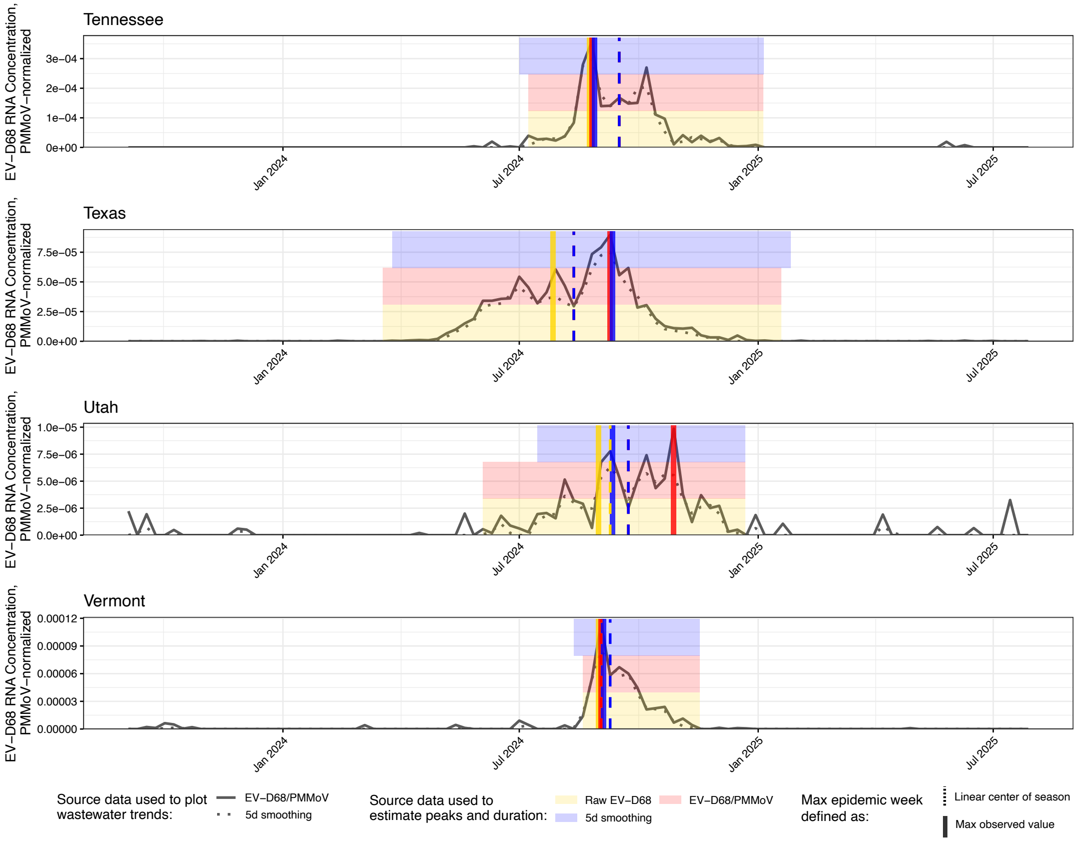
Tennessee
Texas
Utah
Vermont
EV−D68 RNA Concentration, PMMoV−normalized
Jan 2024
Jul 2024
Jan 2025
Jul 2025
Source data used to plot wastewater trends:
EV−D68/PMMoV
5d smoothing
Source data used to estimate peaks and duration:
Raw EV−D68
EV−D68/PMMoV
5d smoothing
Max epidemic week defined as:
Linear center of season
Max observed value

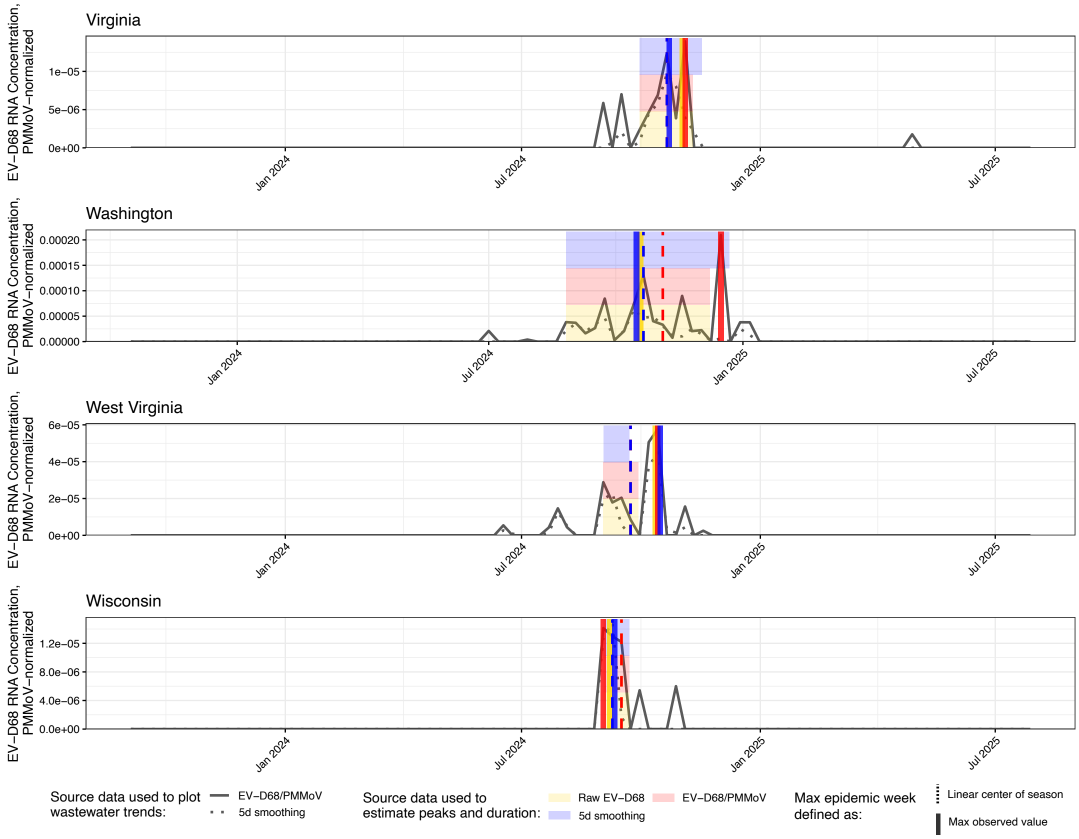


**Fig. S9.** Sensitivity analysis displaying wastewater-derived seasonal variables of EV-D68 RNA concentration in wastewater. For each state, plant-level measurements were aggregated to population-weighted weekly state series, and three data sources were compared: raw EV-D68 RNA concentration (yellow), PMMoV-normalized EV-D68 RNA concentrations (red), and smoothed, PMMoV-normalized EV-D68 RNA concentrations (blue). Black lines display the PMMoV-normalized EV-D68 RNA concentration (solid) and the smoothed, PMMoV-normalized EV-D68 RNA concentration (dotted) to illustrate trends. From each initial data source (namely, raw, PMMoV-normalized, and smoothed, PMMoV-normalized EV-D68 RNA concentrations), three wastewater-derived variables were estimated: the peak week (defined as the week with the highest population-weighted state mean concentration, computed by first taking the arithmetic mean across all measurements within each plant-week and then aggregating across plants using service population as weights), the center of season (defined as the average week of EV-D68 detection, indicating the midpoint of the seasonal activity period based on the timing of detections) and the season duration (defined as the number of epidemiological weeks during which EV-D68 RNA concentrations in wastewater solids remained elevated). For each source, the “max observed value” (defined as the peak week) is shown as a solid, colored vertical bar. The center of season is shown as a colored dashed vertical line. The season duration is depicted as a colored horizontal band spanning the start and end weeks of the detectable period containing the center of season. Duration bands are stacked across the lower, middle, and upper thirds of the y-axis. States without valid data are omitted. Based on this comparison, emphasizing temporal coherence, alignment of peaks and centers, and noise reduction, the smoothed, PMMoV-normalized EV-D68 RNA concentration was selected for retrieval of the wastewater-derived variables center of season and season duration.

**Tables**

**Table S1.** Weekly sampling frequency summary for 147 wastewater treatment plants across the United States, including sampling timeline and sample count range per week..

**Table S2.** Proportion of samples in which EV-D68 was above the detection limit of the molecular assay across 147 wastewater treatment plants.

**Table S3.** Associations between mean environmental conditions and EV-D68 season duration across wastewater treatment plants (WWTPs). Entries are slopes (change in weeks) for fixed increments (temperature and dew point: 5 °C; relative humidity: 10 %RH; precipitation: 5 mm; wind speed: 1 km/h; pressure: 5 hPa). Models are univariate weighted least squares with inverse-variance weights based on within-window variability ($w = 1/SD^2$). *N* indicates the number of WWTPs contributing; for precipitation *n* = 144 because two plants had zero within-window standard deviation and were excluded (weights undefined). Brackets denote 95% confidence intervals; *p*-values are from the weighted models. $R_w^2$ is the weighted $R^2$ of each univariate fit. No geographic covariates (latitude/longitude) were included for duration, as no meaningful associations was observed.

**Table S4.** EV-D68 season duration by sewershed-level determinants. Median (interquartile range, IQR) and mean (± standard deviation, SD) season duration (weeks) are shown for wastewater treatment plants (WWTPs, n=146) grouped by each sociodemographic or structural determinant. The following variables were classified into tertiles: proportion of adults aged ≥65 years, birth rate (per capita), childcare establishment density (per $km^2$), proportion of children ≤5 years, proportion of crowded households (defined as households with more than one person per room), and population density (per $km^2$). Hospitals and nursing homes were divided at the dataset median (2 and 8 per sewershed, respectively). Urbanicity was defined as the proportion of sewershed area designated as urban and classified as ≤50% versus >50%. Airport presence was assessed as a binary variable (absent vs present). P-values indicate overall differences tested using the Kruskal-Wallis test, and pairwise comparisons assessed with Wilcoxon rank-sum tests with Bonferroni correction for multiple testing.

**Table S5.** EV-D68 center of season by sewershed-level determinants. Median interquartile range, IQR) and mean (± standard deviation, SD) center of season are shown for WWTPs grouped by each sociodemographic or structural determinant. Variables in tertiles: proportion of adults aged ≥65 years, birth rate (per capita), childcare establishment density (per $km^2$), proportion of children ≤5 years, proportion of crowded households (defined as households with more than one person per room), and population density (per $km^2$). Hospitals and nursing homes were divided at the dataset median (2 and 8 per sewershed, respectively). Urbanicity was defined as the proportion of sewershed area designated as urban and classified as ≤50% versus >50%. Airport presence was assessed as a binary variable (absent vs present). P-values indicate overall differences tested using the Kruskal-Wallis test, and pairwise comparisons assessed with Wilcoxon rank-sum tests with Bonferroni correction for multiple testing.

**Table S6.** State-level Spearman correlations (ρ) between wastewater PMMoV-normalized EV-D68 RNA concentration and clinical diagnostic indicators. Spearman correlation coefficients (ρ) with Bonferroni-adjusted p-values (p adj.) are reported for associations between wastewater PMMoV-normalized EV-D68 RNA concentrations and the proportion of clinical diagnoses of acute flaccid myelitis (AFM), enterovirus-related encounters, wheezing in children aged ≤5 years, and wheezing in adults aged ≥65 years at the state level. Wastewater measurements were population-weighted by wastewater treatment plant. AFM analyses were conducted using monthly aggregation, whereas other diagnoses used weekly data. "Number of pairs" indicates the number of paired time points (either number of weeks or number of months depending on the diagnosis) included in each correlation.

**Table S7.** Assignment of weather stations to wastewater treatment plants (WWTPs). Each row lists the weather station from which environmental variables were retrieved and the WWTPs associated with that weather station. When multiple WWTPs share the same weather station, all associated WWTP IDs are listed. Weather stations were matched based on geographic proximity and coverage consistency within the respective sewershed regions.

**Table S8.** Description of structural and demographic variables used in the study. Definitions, data sources, computations, spatial resolution, and analytical categorization are shown for each sewershed-level variable. Census-derived proportions were aggregated to the sewershed scale using spatial weighting based on tract overlap. County-level variables were either spatially aggregated to sewersheds or assigned from the predominant county (i.e., the county containing the largest proportion of the sewershed area), as noted. Proportion of adults ≥ 65, proportion of children ≤ 5 and proportion of crowded households were calculated using 2023 5-years American Community Survey data (3). Birth rate was estimated using 2024 US Census Bureau Population Estimates (4). Childcare density was estimated using 2023 County Business Patterns data (5). Urbanicity was calculated as the proportion of each sewershed intersecting urban-designated areas, based on 2020 US Census Bureau Urban-Rural Classification data (6). Population density was defined as the number of residents per square kilometre in the predominant county of each sewershed, using 2022 ACS population estimates (7). Number of hospitals, nursing homes, and airports per sewershed were retrieved from a previous study (8). Categorization schemes reflect how variables were grouped for statistical analysis.
